# Timing of Complementary Feeding for Early Childhood Allergy Prevention: An Overview of Systematic Reviews

**DOI:** 10.1101/2023.06.13.23290959

**Authors:** Paula Kuper, Claudia Hasenpusch, Simone Proebstl, Uwe Matterne, Catherine J. Hornung, Esther Grätsch, Mengtong Li, Antonia A. Sprenger, Dawid Pieper, Jennifer J. Koplin, Michael R. Perkin, Jon Genuneit, Christian Apfelbacher

**Affiliations:** Institute of Social Medicine and Health Systems Research, Medical Faculty, Otto von Guericke University, Magdeburg, Germany; Murdoch Children’s Research Institute, Parkville, Australia; Faculty of Health Sciences Brandenburg, Brandenburg Medical School Theodor Fontane, Institute for Health Services and Health System Research, Rüdersdorf, Germany; Center for Health Services Research, Brandenburg Medical School (Theodor Fontane), Rüdersdorf, Germany; Population Health Research Institute, St. George’s, University London, London, United Kingdom; Pediatric Epidemiology, Department of Pediatrics, Medical Faculty, Leipzig University, Germany; Child Health Research Centre, The University of Queensland, Brisbane; Centre for Food & Allergy Research, Parkville, Australia

**Keywords:** allergic diseases, complementary feeding, overview of reviews, prevention, timing of CF

## Abstract

**Objective:** To summarise and critically appraise systematic review (SR) evidence on the effects of timing of complementary feeding (CF) on the occurrence of allergic sensitisation and disease.

**Design:** Overview of SRs. AMSTAR-2 and ROBIS were used to assess methodological quality and risk of bias (RoB) of SRs. RoB Tool 2.0 was used to assess RoB of primary randomised controlled trials (RCTs) (or extracted). The Certainty of Evidence (CoE) was assessed using GRADE. Findings were synthesised narratively.

**Data sources:** MEDLINE (via PubMed and Ovid), the Cochrane Library and Web of Science Core Collection.

**Eligibility criteria:** SRs investigating the effects of timing of CF on risk of developing food allergy (FA), allergic sensitisation, asthma, allergic rhinitis, atopic eczema, and adverse events in infants or young children (0-3 years), based on RCT evidence.

**Results:** Eleven SRs were included, with predominantly low methodological quality and high RoB. Primary study overlap was very high for specific FA and slight to moderate for FA in general and other primary outcomes. Introducing specific foods (peanut, cooked egg) early probably reduces the risk of specific FA based on evidence across most SRs. The evidence for other allergic outcomes was mostly very uncertain and based on single primary studies. SRs varied regarding the timing of CF, the nature of complementary foods and the population risk, which limited comparability between SRs.

**Conclusions:** The overlap of primary studies within SRs was high to very high for many outcomes, overemphasising single trials. Future research should focus on producing high quality trials and SRs that allow drawing more trustworthy conclusions. For developing guidelines to support decision-making on the timing of CF as a preventive strategy, the early introduction of specific foods (i.e., egg and peanut) seems promising and safe whereas more extensive research is required regarding other allergic outcomes and potential adverse events.

**Registration:** PROSPERO (CRD42021240160); Open Science Forum (https://doi.org/10.17605/OSF.IO/HJKUN)

**Key messages:**

- Evidence supports early introduction of specific complementary foods (peanut, cooked egg) for preventing food-specific allergies
- Evidence regarding prevention of other allergic diseases is sparse and of low certainty
- Future research should focus on producing high-quality trials and reviews for higher certainty of results

## Introduction

Early childhood allergy prevention (ECAP) represents a promising strategy in light of the high burden of allergic diseases among children.^1–3^ Until the early 2000s, avoidance of allergens during pregnancy and lactation, and avoidance or delayed introduction of allergenic foods during infancy was recommended as means to prevent allergy.^4^ However, these recommendations were not sufficiently supported by epidemiological evidence and have since been challenged by new research findings, including observational studies finding a higher risk of developing food allergies when specific allergenic foods were introduced later compared to earlier.^5, 6^

As part of these advances in knowledge, several key randomised controlled trials (RCTs) have been published on the timing of complementary feeding and the subsequent development of atopic diseases, including the LEAP, LEAP-On and EAT studies.^7–9^ Complementary feeding (CF) is defined as the provision of foods and fluids to infants and young children, alongside breastmilk or infant formula when the latter become insufficient to meet the infant’s nutritional needs.^10^ A number of systematic reviews (SRs) on the timing of CF concerning ECAP have already been published to collate and appraise these studies.^11–22^ In a situation like this, overviews of SRs are a powerful tool to provide a single synthesis of the relevant evidence while taking methodological quality and research gaps into consideration.^22^ Overviews can address even broader research questions than single reviews by integrating more outcomes or populations and evaluating inconsistencies across a comprehensive body of evidence. A recently published overview on both RCT and non-RCT (observational) evidence corroborated the results of the aforementioned individual studies showing that the early introduction of peanut and cooked egg probably prevents food-specific allergy. Although the risk of a broad range of long- and short-term outcomes were covered in this overview, the authors did not examine the occurrence of some relevant adverse events such as anaphylaxis or those leading to withdrawal from the study intervention. Systematic investigation of safety outcomes, however, is important for decision-making and informing parents and caregivers by enabling a judgement of the balance between benefits and harms of early CF. This overview aims to provide living systematic evidence by systematically reviewing the RCT evidence synthesised in SRs on the effectiveness and safety of timing of CF for the prevention of allergic sensitisation and diseases.

## Methods

This work is part of living systematic evidence on ECAP, prospectively registered at PROSPERO (CRD42021240160). The protocol for this overview of SRs was prospectively registered at Open Science Framework Registries (https://doi.org/10.17605/OSF.IO/HJKUN).23 Amendments of the protocol are documented in the supplementary material (eAppendix 1). The reporting of this review adheres to the ‘Reporting guideline for overviews of reviews of healthcare intervention’ (PRIOR) (eTable 1).^24^

### Eligibility criteria

Eligibility criteria used to judge SRs regarding their inclusion or exclusion are presented in Table 1.

**Table 1.**
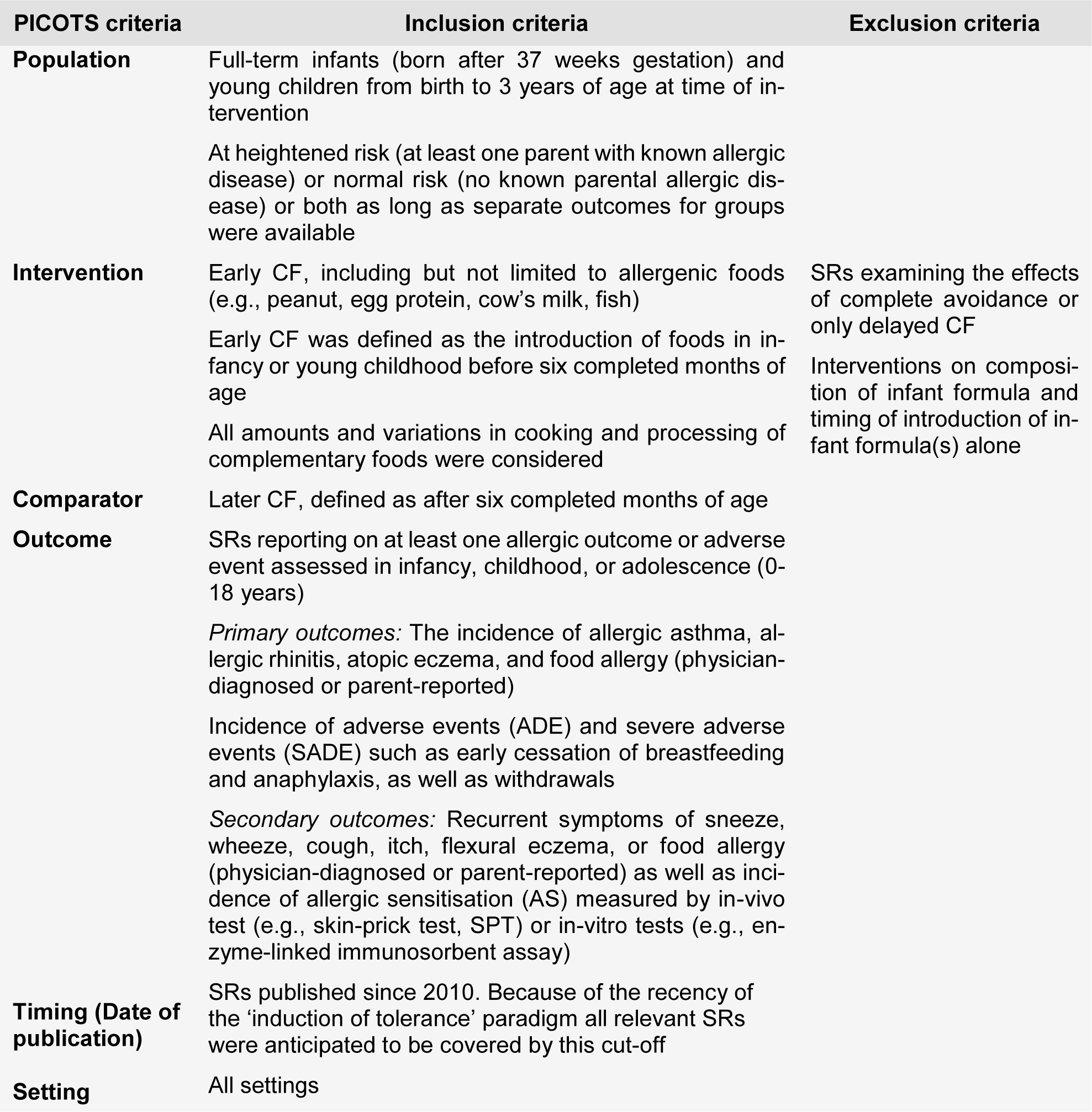
Eligibility criteria according to the PICOTS framework

| PICOTS criteria | Inclusion criteria | Exclusion criteria |
| --- | --- | --- |
| **Population** | Full-term infants (born after 37 weeks gestation) and young children from birth to 3 years of age at time of intervention<br>At heightened risk (at least one parent with known allergic disease) or normal risk (no known parental allergic disease) or both as long as separate outcomes for groups were available |  |
| **Intervention** | Early CF, including but not limited to allergenic foods (e.g., peanut, egg protein, cow's milk, fish)<br>Early CF was defined as the introduction of foods in infancy or young childhood before six completed months of age<br>All amounts and variations in cooking and processing of complementary foods were considered | SRs examining the effects of complete avoidance or only delayed CF<br>Interventions on composition of infant formula and timing of introduction of infant formula(s) alone |
| **Comparator** | Later CF, defined as after six completed months of age |  |
| **Outcome** | SRs reporting on at least one allergic outcome or adverse event assessed in infancy, childhood, or adolescence (0-18 years)<br>*Primary outcomes:* The incidence of allergic asthma, allergic rhinitis, atopic eczema, and food allergy (physician-diagnosed or parent-reported)<br>Incidence of adverse events (ADE) and severe adverse events (SADE) such as early cessation of breastfeeding and anaphylaxis, as well as withdrawals<br>*Secondary outcomes:* Recurrent symptoms of sneeze, wheeze, cough, itch, flexural eczema, or food allergy (physician-diagnosed or parent-reported) as well as incidence of allergic sensitisation (AS) measured by in-vivo test (e.g., skin-prick test, SPT) or in-vitro tests (e.g., enzyme-linked immunosorbent assay) |  |
| **Timing (Date of publication)** | SRs published since 2010. Because of the recency of the 'induction of tolerance' paradigm all relevant SRs were anticipated to be covered by this cut-off |  |
| **Setting** | All settings |  |

We defined SRs using the following criteria:^23^

1. clearly stated objectives with pre-defined eligibility criteria;
2. systematic search that attempts to identify all studies that would meet the eligibility criteria;
3. assessment of the validity of the findings of the included studies (assessment of risk of bias, RoB);
4. systematic presentation and synthesis of the characteristics and findings of included studies;
5. explicit, reproducible methodology including the search strategy, a comprehensive search, and acceptable methods for assessing the validity of included studies.

When updated versions of the same SR were available, only the most recent version was included unless relevant details were only available from earlier versions. No language restrictions were imposed *a priori*.

### Information sources and search strategy

A comprehensive search of MEDLINE (via PubMed and Ovid), the Cochrane Library and Web of Science Core Collection from 2010 was conducted on 13^th^ January 2022. We reran the search on 27^th^ February 2023 to update the initial search using a pre-specified and tested search syntax (eAppendix 2). References of included SRs were hand-searched for potentially relevant SRs. We searched the PROSPERO database for registered SRs and assessed conference abstracts from the European Academy of Allergy and Clinical Immunology congresses. If an SR was commissioned by an agency, we also searched for ‘unpublished’ full reports of the same review for further information.

### Study selection, data extraction and management

Two reviewers screened titles and abstracts independently. Full-texts were obtained and reviewed independently for eligibility in duplicate. SRs were included regardless of their amount of primary study or PICO criteria overlap. Five reviewers extracted data independently, using an adapted form of the summary of findings (SoF) table provided by Cochrane.^25^ Discrepancies and conflicts throughout the process were resolved by discussion or consultation of another reviewer. Data were extracted on study characteristics, outcomes, the certainty of evidence and any information that was required to assess the quality of the SRs. The characteristics and RoB of primary studies were also extracted from the SRs. Discrepant, missing, or unclear data identified during the data extraction process were discussed with reference to the original primary studies, ensuring clear labeling and transparent discussion of data extracted from primary studies rather than SRs. For each outcome and across all outcomes, overlap of RCTs included in the SRs was assessed by calculating the corrected cover area (CCA), as recommended by Pieper et al.^26^ The overlap was calculated and illustrated using the ‘Graphical Representation of Overlap for OVerviews’ (GROOVE) tool.^27^

### Quality and risk of bias assessment of the included reviews

RoB and quality assessment of each included SR were based on the review as a whole, i.e., also considering how non-randomised studies of interventions (NRSI) were incorporated into the respective SR. The revised ‘A MeaSurement Tool to Assess systematic Reviews’ (AMSTAR-2) was used to evaluate the methodological quality of each individual SR.^28^ RoB was evaluated using the ‘Risk Of Bias In Systematic reviews’ tool (ROBIS).^29^ The articles were independently evaluated by two reviewers in duplicate. A comprehensive assessment of the RoB and quality of the included SRs before the updated search is reported elsewhere.^22^ During the updated search, one additional SR was found for which ROBIS and AMSTAR-2 assessments were independently conducted. Disagreements were discussed and resolved by a third reviewer, if necessary.

### Risk of bias assessment of primary studies within included reviews

SRs used a variety of different tools to assess the quality of included primary studies. RoB 2.0 was extracted from the SRs if provided. Otherwise, we re-assessed RoB using the Cochrane RoB 2.0 Excel tool for each relevant result related to our primary outcomes for the effect of assignment to interventions at baseline (‘intention-to-treat’ (ITT) effect).^30, 31^ RoB was assessed for all five domains (randomisation process, deviations from intended interventions, outcome data, measurement of the outcome, selection of the reported result) and overall by assigning a RoB level that was at least severe as the domain which had the highest RoB. If similar RoB judgements for multiple results within one study were deemed highly plausible (e.g., results for multiple assessment times with low drop-out rates), multiple RoB assessments were summarised into one.

### Certainty of evidence (CoE) assessment

We extracted the CoE for each primary outcome within SRs if it was based on the ‘Grading of Recommendations Assessment’ (GRADE) approach, for four out of five domains (imprecision, indirectness, inconsistency, other considerations). As not all SRs reported GRADE assessments, the grading was done fully anew for outcomes within these SRs by using the GRADEPro Tool.^32, 33^ The fifth domain “RoB” was re-assessed for all outcomes where RoB was also reassessed with the RoB 2.0 Tool. The GRADEPro tool was used to integrate up- and/or down-grades of the individual domains into an overall judgement of the quality of the body of evidence resulting in one of four grades: high, moderate, low or very low (for a definition see eTable 8). If an outcome was based on one single primary study, the domain “inconsistency” across studies was not applicable and the overall evaluation was based on the remaining domains. The grading was based on information provided in the SRs and primary studies they referred to. For secondary outcomes, all approaches assessing the CoE were accepted and extracted.

### Data synthesis

The results are presented narratively and are supported by a SoF table showing the extracted numerical results that were provided by the SRs. If the scope of the SR was broader than the scope of the overview, only results for comparisons that were relevant according to the predefined inclusion criteria were extracted, i.e., comparisons based on RCT evidence and related to our predefined outcomes. GRADE was re-assessed if existing assessments referred to a larger evidence base. The results and their certainty are presented grouped by outcome, timing of the intervention and SR to show differences between SRs. We report and discuss clinical heterogeneity across the reviews based on the extracted study characteristics. Statistical heterogeneity was reported as presented in the SRs (e.g. *I*^2^ statistics, between-study variance *τ*^2^) using a rough interpretation of the *I*^2^ statistics (low: 0–25%, moderate: 25–75%, high: 75–100%).^34^ If available, the results of sensitivity analyses regarding missing data, publication bias and stratification by RoB were extracted from the SRs. Results of subgroup analyses are considered in the SoF table and addressed in the reporting of the results.

## Results

Titles and abstracts of 3969 articles were examined for eligibility and 3918 articles excluded. Of 52 examined full-texts, eleven articles were identified as eligible including a total of 48 primary studies. Figure 1 shows the selection process. A list of excluded reviews with reasons for exclusion after full-text screening is given in eTable 2.

**Figure 1.**
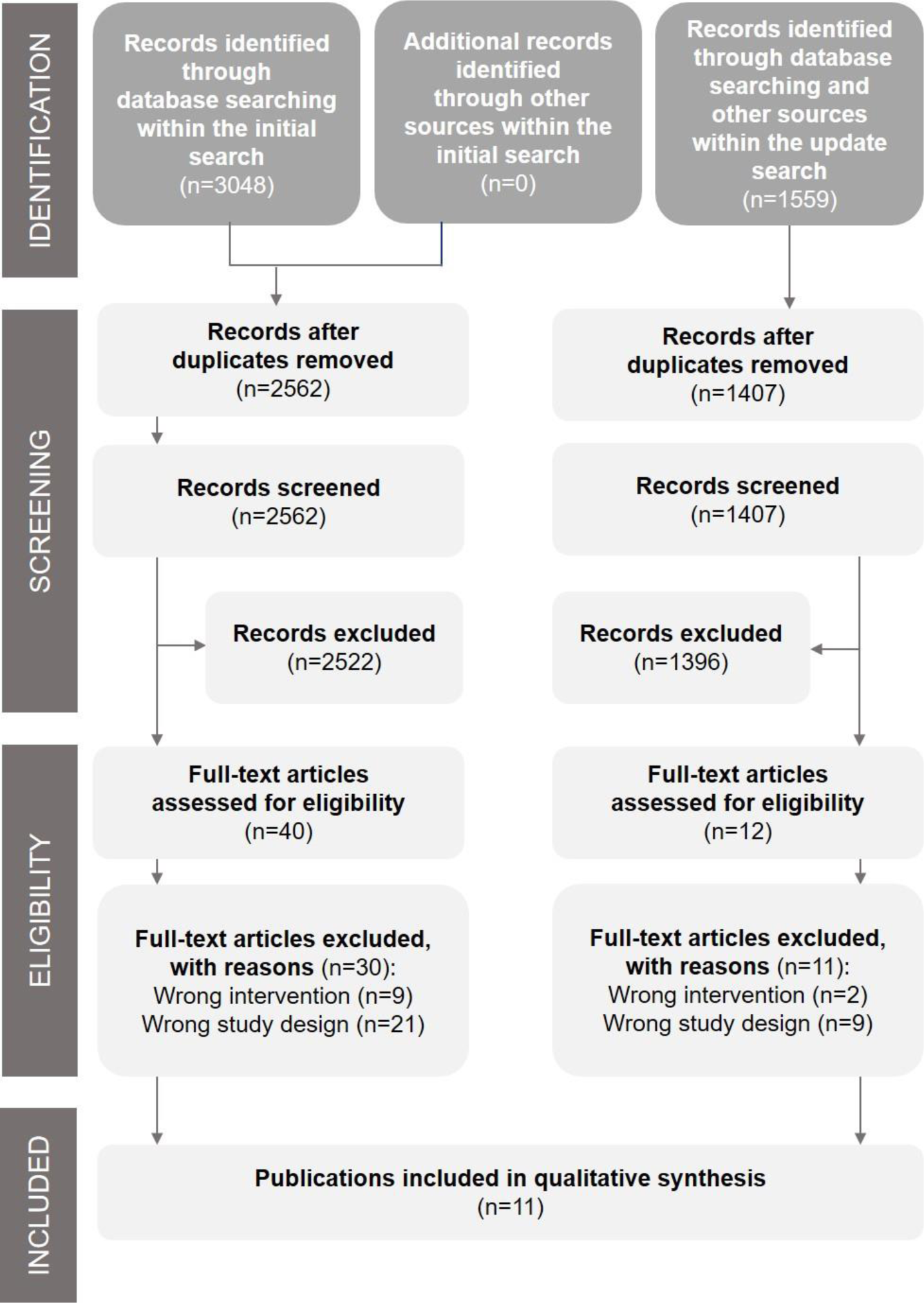
PRISMA Flow Diagram of the overview of systematic reviews

### Characteristics of systematic reviews

Table 2 summarises the characteristics of the SRs, with more detailed information provided in eTable 3. The reviews varied with respect to the definition of timeframes for CF, with an early food introduction defined as from around three months or later^11^ or before 6 months^15, 20^, whereas other SRs investigated the age at CF without further classification.^12, 13, 16, 18, 19, 21^ Two SRs compared late to early introduction before 6 or 12 months, respectively.^14, 17^ The complementary foods that were introduced also varied across and within SRs, encompassing both potentially allergic and non-allergic foods, as well as single or multiple foods as part of either multifaceted or single interventions. One SR focused solely on healthy infants^20^ while the remainder examined both children at population risk and high-risk or did not further restrict the population regarding the baseline allergy risk.^11–19, 21^

**Table 2.**
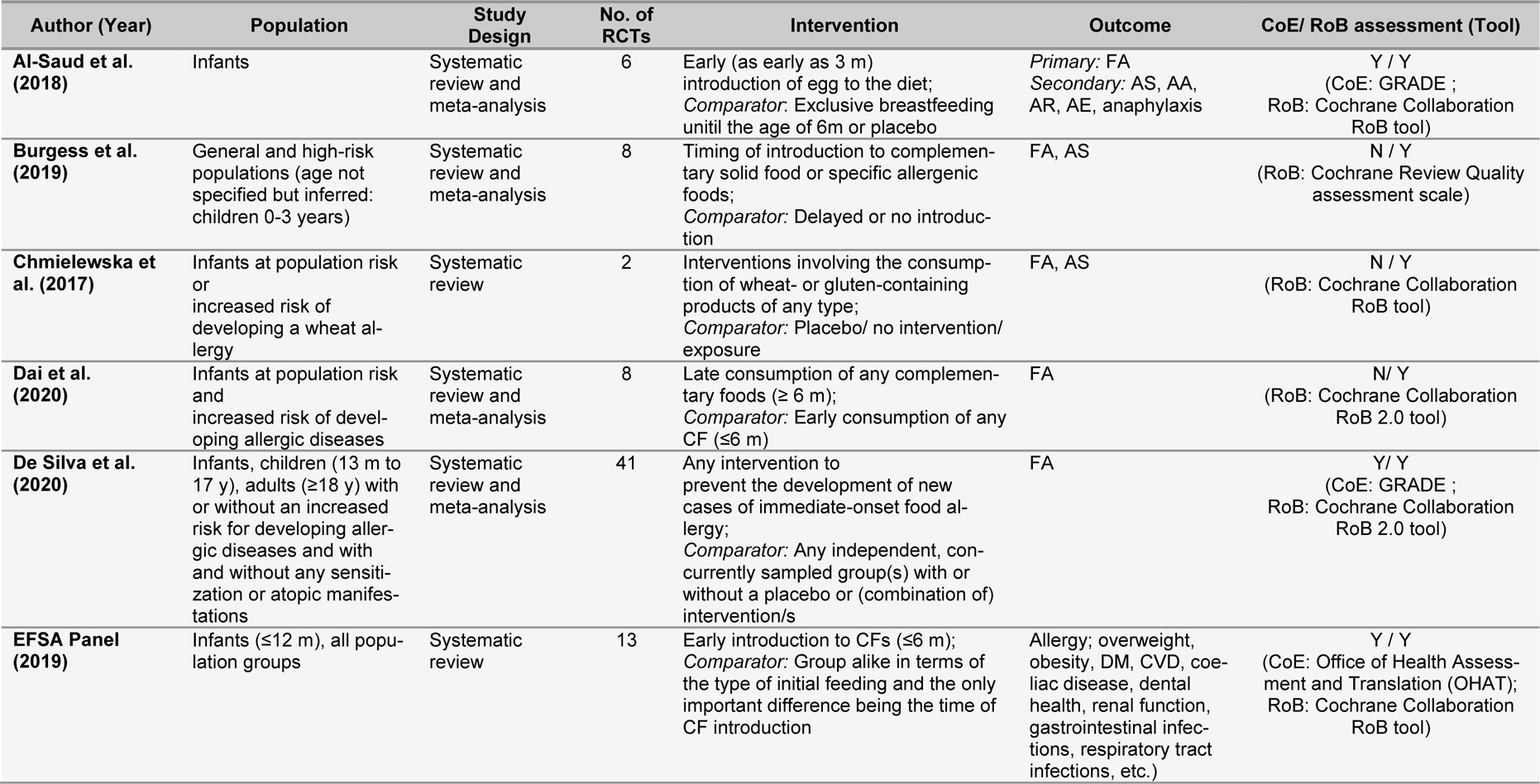
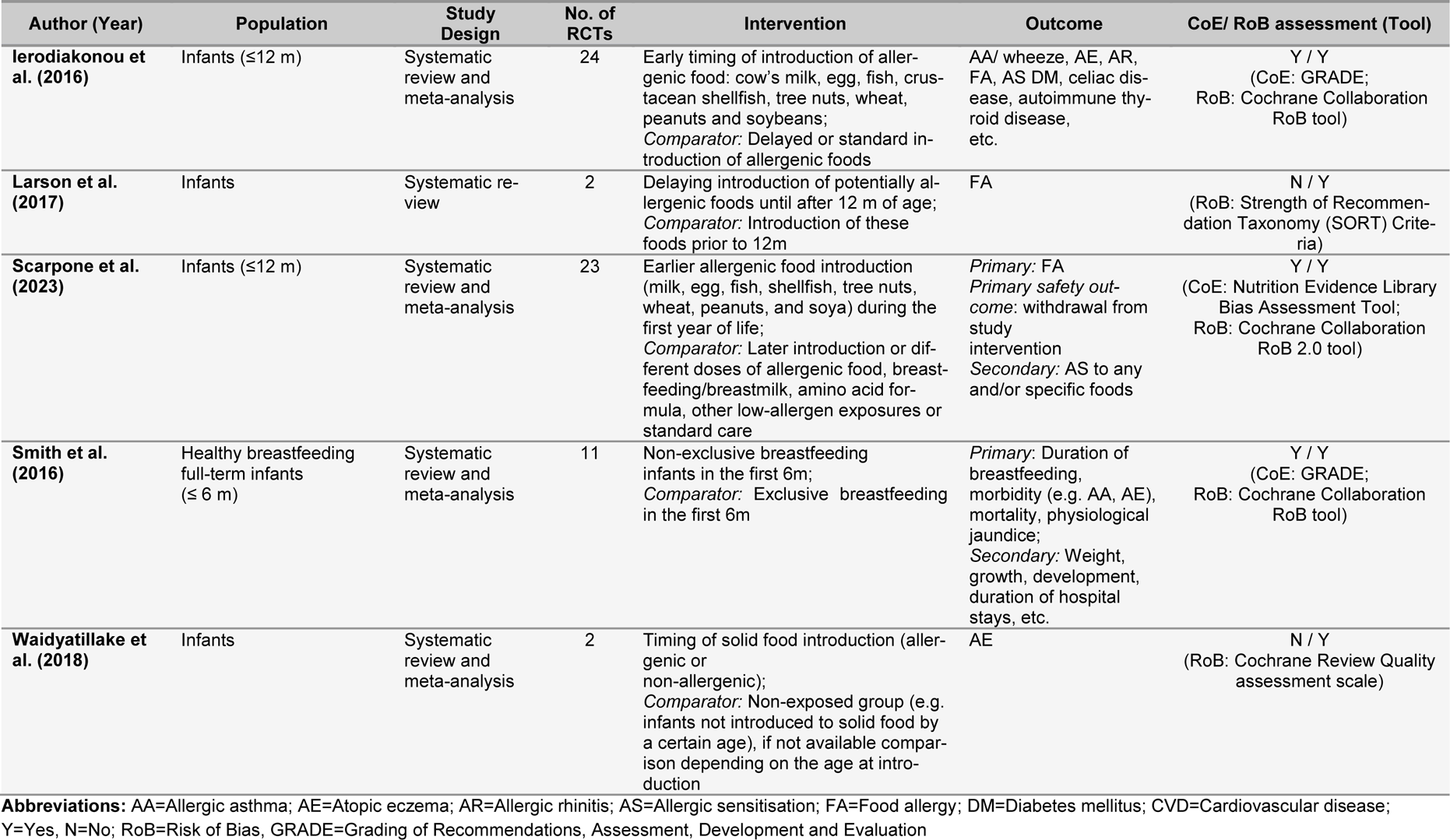
Characteristics of included systematic reviews

### Risk of bias and methodological quality of systematic reviews

Methodological quality and RoB of the initially included reviews are published elsewhere.^22^ Assessments for the SRs identified during the update search can be found in eTables 4-5. All SRs defined the PICO components adequately. Nearly all SRs established their methods prior to conducting the review, assessed RoB of the included studies, reported conflicts of interest, and performed data extraction in duplicate. Common limitations were not elaborating on the funding of the primary studies or the decision for selecting specific study designs, and not providing a list of the excluded studies with a justification. The RoB assessment found that most SRs hat major deficiencies in at least two stages of the review process, from study eligibility to synthesis and findings.^11–15, 17, 19–21^ Only two reviews conducted by a similar team of authors were rated as overall low RoB.^16, 18^

### Risk of bias of primary studies included in systematic reviews

Two SRs reported RoB of primary studies based on the Cochrane RoB 2.0 Tool.^18, 19^ However, one^19^ did not perform assessments at the result-level as recommended by Cochrane.^35^ Thus, RoB was re-assessed for all relevant primary study results reported in all but one SR^18^ and is presented in eTable 6, RoB for all other outcomes is shown in eTable 7. RoB assessment regarding (serious) adverse events was not applicable because quantitative effect estimates and/or detailed information were rarely reported in primary studies. Most assessments of primary studies yielded “some concerns” or “high” RoB for each outcome. Across all outcomes, common limitations of primary studies were lack of blinding of outcome assessment, high loss to follow-up while insufficiently examining potential bias introduced by missing data and lack of preregistration including a sufficiently detailed analysis plan. Studies published longer ago were more affected by these limitations.

### Certainty of evidence

The tools used for assessing the CoE in the SRs are presented in Table 2, showing that five SRs did not grade the CoE.^12–14, 17, 21^ eTable 8 shows the overall CoE for primary outcomes based on the GRADE approach (see eTable 9 for assessments of all domains) and eTable 10 shows the extracted CoE for secondary outcomes and adverse events based on the reported approaches in the SRs. The level of certainty was found to be very low to low for most of the outcomes except for the risk of developing egg and peanut allergy in some SRs^11, 14, 18^ and eczema in two SRs.^11, 20^ The most common reasons for rating down the CoE were high RoB of the body of evidence, imprecision and nongeneralisability of the study populations.

### Summary of results

A summary of key quantitative primary findings is presented in Table 3 along with the respective CoE (more detailled see eTable 8). eTable 10 shows findings for secondary outcomes and adverse events, where available. All reviews adhered to their pre-defined research questions and analyses. In cases where the SR authors addressed the presence of reporting or publication biases, this is described in the comments section of eTables 8 and 10 and considered in the respective GRADE assessments and in the reporting of results. The total primary study overlap across all outcomes was high (CCA=11%; see eFigure 1). eTable 11 shows the overlap for each outcome separately.

**Table 3.** Summarised results of the GRADE assessments

| Author (Year) | No. of RCTs (participants) | Effect estimate (95% Confidence interval) | Certainty of Evidence |
| --- | --- | --- | --- |
| **Outcome: Risk of food allergy** |  |  |  |
| **Exposure: Allergenic food (alone or in combination)** |  |  |  |
| Scarpone et al. (2023) | 4 (3295) | Introduction of multiple foods: RR 0.49 (0.33-0.74) | Moderate |
| Scarpone et al. (2023) | 4 (3250) | Introduction of wheat: RR 0.40 (0.20-0.79) | Low |
| Scarpone et al. (2023) | 4 (3295) | Introduction of egg: RR 0.49 (0.33-0.74) | Very Low |
| Burgess et al. (2019) | 5 (3927) | Introduction of peanut: RR 0.60 (0.38-0.94) |  |
| De Silva et al. (2020) | 1 (1303) | RR 0.80 (0.51-1.25)<sup>a</sup> |  |
| Burgess et al. (2019) | 1 (1303) | RR 0.80 (0.51-1.25)<sup>a</sup> |  |
| EFSA Panel (2019) | 1 (1162) | RR 0.80 (0.51-1.25)<sup>a</sup> |  |
| Smith et al. (2016) | 1 (1162) | RR 0.80 (0.51-1.25)<sup>a</sup> |  |
| **Outcome: Risk of egg allergy** |  |  |  |
| **Exposure: Egg (alone or in combination)** |  |  |  |
| Scarpone et al. (2023) | 9 (4811) | RR 0.60 (0.46-0.77) | High |
| Dai et al. (2019) | 6 (2663) | RR 0.60 (0.46-0.79) |  |
| Al-Saud et al. (2018) | 6 (2663) | RR 0.60 (0.44-0.82) |  |
| Burgess et al. (2019) | 6 (3085) | OR 0.63 (0.44-0.90) | Moderate |
| De Silva et al. (2020) | 1 (147) | Introduction of cooked egg: RR 0.22 (0.09-0.54) | Low |
|  | 3 (1289) | Introduction of raw/ pasteurised egg: RR Range 0.65- 3.30 |  |
| EFSA Panel (2019) | 2 (1569) | General population: RR 3.30 (0.35-31.32) |  |
|  | 3 (1087) | At-risk population: RR 0.69 (0.51-0.93) |  |
| Larson et al. (2017) | 1 (86) | No difference |  |
| **Outcome: Risk of peanut allergy** |  |  |  |
| **Exposure: Peanut (alone or in combination)** |  |  |  |
| Scarpone et al. (2023) | 4 (3796) | RR 0.31 (0.19-0.51) | High |
| Burgess et al. (2019) | 2 (1943) | OR 0.28 (0.14-0.57) | Low |
| De Silva et al. (2020) | 2 (640) | RR Range 0.14-0.35 | Very Low |

| Outcome: Risk of peanut allergy |  |  |  |
| --- | --- | --- | --- |
| Exposure: Peanut (alone or in combination) |  |  |  |
| EFSA Panel (2019) | 1 (1168) | RR 0.49 (0.20-1.19) | Very Low |
| Larson et al. (2017) | 1 (640) | Low risk infants: RD 11.8% |  |
|  |  | High risk infants: RD 24.7% |  |
| Ierodiakonou et al. (2016) | 2 (1793) | RR 0.29 (0.11-0.74) |  |
| Dai et al. (2019) | 2 (1793) | n. a. | N. A. |
| Outcome: Risk of grain allergy |  |  |  |
| Exposure: Cereals (wheat) (alone or in combination) |  |  |  |
| Chmielewska et al. (2017) | 1 (1303) | 1/572 vs. 0/597 | Low |
| EFSA Panel (2019) | 1 (1169) | n. a. |  |
| Scarpone et al. (2023) | 3 (3169) | RR 0.66 (0.10-4.47) | Very Low |
| Outcome: Risk of cow's milk allergy |  |  |  |
| Exposure: Cow's milk (alone or in combination) |  |  |  |
| Dai et al. (2019) | 2 (1550) | n. a. | N. A. |
| Ierodiakonou et al. (2016) | 2 (1550) | RR 0.76 (0.32-1.78) | Ver Low |
| Scarpone et al. (2023) | 6 (3900) | RR 0.84 (0.38-1.87) |  |
| Outcome: Risk of developing eczema |  |  |  |
| Exposure: Allergenic food (alone or in combination) |  |  |  |
| Al-Saud et al. (2018) | 2 (1001) | Introduction of egg powder: RR 0.90 (0.70-1.16) | Moderate |
| Smith et al. (2016) | 1 (284) | RR 0.86 (0.51-1.44) |  |
| Waidyatillake et al. (2019) | 2 (1139) | OR 0.87 (0.68-1.12) | Low |
| EFSA Panel (2019) | 2 (1001) | Introduction of egg: OR 0.82 (0.01-130.12) | Very Low |
| Ierodiakonou et al. (2016) | 8 (2333) | Introduction of cow's milk (outcome at ≤4 years): RR 1.14 (0.87-1.49) |  |
|  | 6 (1665) | Introduction of cow's milk (outcome at 5-14 years: RR 1.05 (0.90-1.23) |  |
| Outcome: Risk of developing asthma |  |  |  |
| Exposure: Allergenic food (alone or in combination) |  |  |  |
| EFSA Panel (2019) | 1 (1127) | OR 0.98 (0.47-2.05) | Low |
|  | 1 (820) | Introduction of egg: RR 1.12 (0.87-1.45) |  |
| Outcome: Risk of developing allergic rhinitis |  |  |  |
| Exposure: Allergenic food (alone or in combination) |  |  |  |
| EFSA Panel (2019) | 1 (1127) | n. a. | N. A. |
| Ierodiakonou et al. (2016) | 6 (1735) | Introduction of cow's milk (outcome at ≤4 years): RR 1.52 (0.97-2.37) | Low |
|  | 5 (1673) | Introduction of cow's milk (outcome at 5-14 years: RR 1.21 (0.85-1.73) |  |

**Abbreviations:** n. a.= not available; N. A.= not applicable

### Risk of developing food allergy (in general)

Five SRs investigated the effects of timing of CF on the risk of developing food allergy in general with a moderate primary study overlap (CCA=9%).^12, 15, 18–20^ Among these, four reviews relied on a single primary RCT study in the general population finding very uncertain evidence about an association of introducing multiple foods early with the risk of FA in the ITT sample, and a lower risk in the per-protocol (PP) sample that adhered to the assigned dietary regimen.^12, 15, 19, 20^ Scarpone et al. included newer trials (14% overlap with the other reviews) with populations at average and at increased allergy risk and examined the effects of early introduction of both single and multiple allergenic foods on the risk of developing FA.^18^ The early introduction of multiple allergenic foods likely decreased the risk of FA, supported by a sensitivity analysis that was restricted to studies with low RoB.^18^ There was very uncertain evidence indicating a protective effect on FA when separately introducing egg, peanut and wheat. No effect was shown on FA in the groups that introduced cow’s milk, soya, fish and crustaceans and nuts early versus late, with high imprecision and very uncertain evidence.^18^

### Risk of developing egg allergy

A total of eight reviews investigated the risk of developing egg allergy with a very high primary study overlap (CCA=52%).^11, 12, 14–19^ Five reviews showed a (probably) reduced risk regarding the introduction of egg in general or cooked egg in mixed populations across studies after 12-36 months, with moderate to high certainty.^11, 12, 14, 18, 19^ Two SRs showed similar findings with low to very low CoE.^15, 16^ Differences in certainty ratings were due to the broader evidence base, with high RoB in Ierodiakonou et al.,^16^ and because SRs graded the dimension “indirectness” differently. Findings of two reviews by Larson et al.^17^ and De Silva et al.^19^ (25% overlap) suggested no group difference in the risk of egg allergy at 12-36 months when introducing raw egg or pasteurized raw egg powder. Subgroup analyses in three trials showed a more pronounced effect in studies using lower doses of egg.^11, 14, 18^

### Risk of developing peanut allergy

The risk of developing peanut allergy up to 72 months has been studied frequently in a very similar set of primary studies across SRs with very high overlap (CCA=30%). Four reviews suggested a reduced risk of peanut allergy due to early introduction of peanut.^12, 16, 17, 19^ Scarpone et al. also showed a risk reduction for early peanut introduction, including more recent evidence (20-50% overlap to the other SRs).^18^ Sensitivity analyses with low RoB studies corroborated this finding.^18^ Evidence regarding the prevention of peanut allergy through early introduction of multiple allergenic foods is very uncertain.^15^

### Risk of developing allergy to other specific allergenic foods (e.g., wheat, cow’s milk)

Two reviews^13, 15^ examined the same single study and found evidence suggesting that introducing multiple foods early may result in little to no difference in the risk of developing wheat allergy at 12-36 months in the general population. This was also shown by the results of a recent high quality review conducted across at risk and general populations (very high overall overlap; CCA=33%).^18^

Ierodiakonou et al. and Scarpone et al.^16, 18^ found very uncertain evidence regarding the effect of timing of CF on the risk of cow’s milk allergy (33% overlap) across the introduction of single and multiple foods in high-risk and average-risk populations. Dai et al.^14^ drew no conclusions due to large clinical heterogeneity (moderate overall overlap; CCA=6%). Results for developing food allergies to other common allergenic foods (soy and fish) were only studied in one SR and are presented in eTable 8.

### Risk of allergic sensitisation to any food

The evidence of two SRs (very high overlap; CCA=25%) suggested no difference in the risk of any food sensitisation between early and late introduction groups at 12, 36 or after 11-36 months across at-risk and general populations.^14, 17^ Another SR suggested similar findings based on studies investigating the effect of introducing single foods such as egg, peanut, cow’s milk, and wheat.^18^

### Risk of allergic sensitisation to egg

The primary study overlap across SRs was high (CCA=36%). Two SRs (100% overlap) found that early introduction of egg across populations with varying baseline allergy risks likely resulted in a reduced risk of egg sensitisation after 12 months.^11, 12^ A more recent review^18^ found a similar effect across 8 RCTs (62,2% overlap to the afore mentioned SRs) as well as sensitivity analyses in Ierodiakonou et al.^16^ that excluded studies at unclear RoB or abstract publications. However, the main analysis did not support this result^16^ and neither did the findings of two other SRs^15, 17^ for an age of 12 or 36 months for the general and high-risk population at 12 months, with unknown CoE.

### Risk of allergic sensitisation to peanut

There was no difference between groups in risk of sensitisation to peanut in the general population in two SRs based on one primary study (100% overlap) with unknown CoE.^15, 16^ A recently published SR suggested the same finding across populations with varying baseline risk (very high overall overlap; CCA= 25%).^18^

### Risk of allergic sensitisation to other common foods

Four SRs examined the risk of developing wheat, cow’s milk and other common food sensitisation.^13, 15, 16, 18^ The overlap was very high for wheat and cow’s milk and the results are presented in eTable 4.

### Risk of developing eczema

There was a slight overlap between SRs (CCA=4%).^11, 15, 16, 20, 21^ The risk of developing eczema after 12 months probably did not differ by the age of introducing egg or a combination of multiple potentially allergenic foods.^11, 20^ The evidence in one SR^21^ suggested similar results, whereas the evidence in another SR was judged to be very uncertain.^15^ CoE reportings deviated due to different evidence bases and different assessments of the generalisbility in two SRs.^11, 15^ Ierodiakonou et al. included an older evidence base showing that multifaceted interventions may have little to no effect on the risk of developing eczema up to four and after 5-14 years in average-risk and high-risk populations.^16^ Subgroup analyses did not show differences of the effect conditioned on the risk status, RoB or the type of intervention (multifaceted vs. not multifaceted).^16^

### Risk of developing asthma

One SR by the EFSA Panel found no evidence suggesting an association between the timing of CF and the development of asthma-like symptoms in the general population up to 36 months or at 12 months.^15^ Other SRs investigated asthma using wheeze as an outcome, for which the results are presented in eTable 10.

### Risk of developing allergic rhinitis

Two SRs^15, 16^ studied the effect of timing of CF on the risk of developing allergic rhinitis (0% overlap). Ierodiakonou et al. synthesised findings across populations with average and high risk, showing that early introduction of cow’s milk resulted in little to no difference in the risk of developing eczema up to four years and 5-14 years.^16^ High heterogeneity could be partially explained by excluding one primary study, which did not change the overall conclusion. The SR by the EFSA Panel showed results in a similar range but relied on a single study with unknown certainty.^15^

### Risk of withdrawal from the study intervention

Scarpone et al. systematically examined the risk of withdrawal as a primary outcome and reported that early introduction of cow’s milk probably may result in no difference between intervention groups.^18^ They also found that the risk of withdrawal probably increases with an early introduction of multiple allergenic foods at 12-60 months and may increase when introducing egg. Evidence was uncertain when introducing peanut.

### Risk of (serious) adverse events

The risk of anaphylaxis was reported in two SRs (0% overlap).^11, 15^ Al-Saud et al. investigated the occurrence of adverse events, reporting no effect of timing of CF on the occurrence of anaphylaxis in an at-risk population (GRADE not available, n.a.).^11^ The SR by the EFSA Panel^15^ reported that one RCT conducted in an average-risk population stopped recruiting early because allergic symptoms were more common in the intervention group, among other reasons (GRADE n.a.).

Two SRs^15, 20^ reported no group differences in growth-related outcomes (body length/ height changes) between the early and late CF (GRADE n.a.). Smith et al. found that early introduction of potentially allergenic foods may not result in a mean difference between the two groups in terms of days the infants had fever.^20^ They also reported no association of early CF with the median duration of breastfeeding in an average-risk population^9^, upper respiratory illness and diarrhea in healthy children and infant mortality at discharge (GRADE n.a. for all outcomes).

## Discussion

We identified eleven SRs of mostly poor methodological quality and high RoB published between 2016-2023, including 48 primary studies investigating the risk of developing allergy in infants without allergy, based on RCT evidence.^22^ There was a very high primary study overlap between SRs investigating the risk of developing specific FA and sensitisation and a moderate overlap regarding the risk of developing FA in general. For other allergic outcomes and adverse events, the overlap was slight or not assessable because only single SRs investigated these research questions.

Evidence was most extensive and certain regarding the prevention of egg and peanut allergy (very high primary study overlap), indicating a probably reduced risk by early introduction of cooked egg and peanut. Inconclusive evidence was found for preventing FA in general (moderate overlap). Based on newer RCT evidence, the introduction of multiple allergenic foods may decrease the risk of developing FA. Results of older SRs based on one primary study, however, provide very uncertain evidence for this outcome. The evidence for the prevention of FA in general and to specific foods by introducing single allergenic foods was also very uncertain. Introducing egg powder or multiple allergenic foods early probably does not reduce the risk of developing eczema in populations at average and heightened risk. Scarce evidence suggested that early introduction of multiple allergenic foods, egg and cow’s milk does not reduce the risk of allergic asthma and rhinitis.^15, 16^ Regarding safety outcomes, one SR examined study withdrawal as a primary outcome, finding that introducing multiple foods likely increases the risk of withdrawal.^18^ The early introduction of egg may also increase the risk of withdrawal from the study. Early CF appeared to be safe, but this conclusion is limited due to a sparse evidence base and unknown CoE. Findings suggest that introducing egg, peanut, cow’s milk or wheat early does not reduce the risk of any allergic sensitisation or sensitisation to these specific foods, except for probably reducing the risk of egg sensitisation.^18, 20^

The body of evidence is dominated by a few landmark primary studies (especially up to 2017), which is reflected in a high to very high overlap of primary studies for many outcomes across SRs.^9, 36–40^ Only one recently published SR by Scarpone et al. added new primary studies to the evidence base, i.e. RCTs conducted after 2017.^18^ This lack of extensive research for most outcomes within SRs overemphasised results from single RCTs that were repeatedly included as the only available evidence in multiple SRs. Moreover, the studies were often underpowered and not designed to examine certain rare events such as anaphylaxis. Hence, it is important to recognize that the absence of evidence should not be misconstrued as evidence for absence, especially for safety outcomes and rare allergic diseases. Moreover, the CoE was often downgraded due to large inconsistency between effects which is presumed to be a result of differences between primary studies in terms of the nature and doses of foods, control group designs, populations (high-risk or general), outcome measurement and time periods of CF.

### Limitations and strengths

Most of the current research was conducted in high income countries and populations with high prevalence of specific allergies which limits generalisability to low and middle income countries. The overview relied on the analyses conducted in the SRs and thus might not have captured heterogeneity within SRs. Only a few studies explored heterogeneity, publication bias, or conducted subgroup analyses, which could not be summarised as the evidence was too scarce and of low CoE.

We strictly adhered to the PRIOR statement and focused on results based on RCTs to rely on the best available evidence regarding causal intervention effects.^24^ RoB was re-assessed for all primary studies using the most recent version of the RoB 2.0 Tool to provide comparable results for RoB and CoE. The overview allowed to identify evidence gaps in primary studies that were reflected in the SRs, which may be used to guide future research in this field.

### Integration in previous literature

One overview with a similar research question was recently published, investigating the association between the age of CF in the first year of life of infants and various health outcomes, also including observational evidence.^41^ Findings were in line with our findings on the likely preventive effect of early introduction of eggs and peanuts. Additionally, the authors concluded that there is uncertain evidence on the effects of early CF on food allergy, eczema and impaired growth (ADE) and that there is a lack of extensive evidence regarding asthma and allergic rhinitis.

### Implications for research and practice

Although the findings of this overview underline the potential of early introduction of specific foods for the prevention of specific food allergies, it remains unclear whether the timing of CF in general is effective in preventing food allergy (as a whole) and other allergic diseases. Current recommendations on the timing of CF from different international organisations vary slightly. The World Health Organization recommends the introduction of CFs at six months of age.^42^ Similarly, guidelines published by the American Academy of Pediatrics^43^ and the Australian government state that starting complementary feeding around six months of age is optimal.^44^ According to the European Society for Paediatric Gastroenterology, Hepatology, and Nutrition, CF should not commence before four months but should not be postponed beyond six months.^45^ The report by the EFSA panel found no evidence that introduction before six months of age confers any benefits or harms but that ‘age and development appropriate’ earlier introduction is acceptable.^15^ Evidence on safety outcomes synthesised in this review supports that conclusion, although higher quality research is needed to corroborate these findings. Future research should also examine more extensively any health-related risks in infants that adhere to the dietary regimen.

## Conclusion

Parents and healthcare professionals are faced with dynamic knowledge regarding ECAP with the number of SRs exceeding the number of RCTs for almost all exposure-outcome comparisons. Therefore, clear public health messaging about effective prevention measures is necessary. The high overlap of primary research amongst SRs overemphasises landmark trials and the limited evidence base makes it difficult to adequately investigate sources of heterogeneity or publication bias in SRs. Current evidence does not support early CF for preventing FA in general, but rather for preventing allergy to specific foods. Higher quality research is required to evaluate benefits and potential short-term and long-term harms of early CF in infants.

## Data Availability

The data underlying this article will be shared on reasonable request to the corresponding author.

## List of Abbreviations

ADE: Adverse events
AMSTAR-2: A MeaSurement Tool to Assess systematic Reviews
AS: Allergic sensitisation
CCA: Corrected Cover Area
COE: Certainty of Evidence
CF: Complementary food
ECAP: Early Childhood Allergy Prevention
EFSA: European Food Standards Authority
FA: Food Allergy
GRADE: Grading of Recommendations, Assessment, Development and Evaluation
GROOVE: Graphical Representation of Overlap of OVerviews
ITT: Intention-to-treat
LEAP: Learning Early About Peanut allergy study
n.a.: Not available
NRSI: Non-randomised studies of interventions
PP: Per-protocol
PRIOR: Preferred Reporting Items for Overviews of Reviews
RCT: Randomised Controlled Trials
ROB: Risk of Bias
RoB 2.0 Tool: Cochrane risk-of-bias tool for randomised trials
ROBIS: Risk Of Bias In Systematic reviews tool
SADE: Severe adverse events
SPT: Skin-prick test
SR: Systematic Review
WS: Wheat Sensitisation

## Declarations

### Author’s Contributions

CA and UM conceived the idea for the review. CA obtained funding, provided resources, coordinated and supervised the review. DP and JG supervised the methodology of the review and reviewed the manuscript. UM and JW performed the initial search and screening, data extraction, AMSTAR-2 and ROBIS assessment and reviewed the manuscript. PK and CH carried out the update search, screening, data extraction, RoB, GRADE, AMSTAR-2 and ROBIS assessment and wrote the manuscript. SP was involved in the writing of the study protocol, performed data extraction, and wrote the manuscript. CaH performed RoB assessments and reviewed the manuscript. EG and ML carried out data extraction and reviewed the manuscript. ML performed RoB and GRADE assessment, especially for reviews in Chinese (Mandarin). MP, AS and JK reviewed the manuscript. All authors read and approved the final manuscript.

## Acknowledgements

We would like to thank Dr. Jiancong Wang, Dr. Melissa Theurich, Anna Xu, Marco Strecker, and Aiad Hasoon for supporting the research team during the course of the study.

## Funding

This work was supported by the German Research Foundation (DFG, Deutsche For-schungsgemeinschaft), partially funded by the subprojects of DFG Research Group FOR 2959 (HELICAP), AP 253/3-1, project number 409800133.

HELICAP is an association of leading scientists at four locations in Germany. Besides the University of Magdeburg, the University of Education Freiburg, the University of University of Freiburg, the University of Regensburg and the University of Hannover are involved in the interdisciplinary research group HELICAP. Members of the HELICAP Steering Group are Prof. Dr. Christian Apfelbacher, Prof. Dr. Eva Maria Bitzer, Dr. Susanne Brandstetter, Dr. Janina Curbach, Prof. Dr. Marie-Luise Dierks and Prof. Dr. Markus Antonius Wirtz.

### Data sharing

The data underlying this article will be shared on reasonable request to the corresponding author.

## Supplementary material

eAppendix 1

eAppendix 2

