## Supplementary Material for "Timing of Complementary Feeding for Early Childhood Allergy Prevention: An Overview of Systematic Reviews"

### Supplementary Online Content

**eFigure 1.** Overall Study Overlap of Primary Studies within Systematic Reviews (figure and formula)

**eTable 1.** Adherence to the PRIOR criteria

**eTable 2**. Excluded Systematic Reviews After Full-text Screening and Reasons for Exclusion

**eTable 3.** Characteristics of Included Systematic Reviews

**eTable 4.** Methodological Quality of the Included Systematic Reviews (AMSTAR-2 checklist)

**eTable 5.** Risk of Bias in the Included Systematic Reviews (ROBIS)

**eTable 6.** Risk of Bias of Primary Studies within Systematic Reviews

**eTable 7.** Risk of Bias of Primary Studies Within Systematic Reviews Based on Tools Used in the Systematic Reviews for Secondary Outcomes/ Over-all Studies

**eTable 8.** Summary of Findings of Primary Outcomes

**eTable 9.** Certainty of Evidence Assessments (GRADE approach) for all Primary Outcomes

**eTable 10.** Summary of Findings of Secondary Outcomes

**eTable 11.** Outcome related Study Overlap of Primary Studies within Systematic Reviews

**eAppendix 1 Deviations from the Protocol**

**eAppendix 2 Search Strategies**

**Literature**

### eFigure 1. Study Overlap of Primary Studies within Systematic Reviews

**Formula:** Overlap of RCTs included in the SRs and investigating timing of complementary feeding was assessed using by calculating the corrected cover area (CCA), as recommended by Pieper et al.[1].

$$CCA = \frac{N-r}{r*c-r}$$

N = Number of primary studies included in SR (double counting)
r = Total number of publications from primary studies
c = Number of included primary studies

| **55** | **=** | **Total nodes (pairs of reviews)** |
|---|---|---|
| 8 | = | Slight overlap (<5%) |
| 9 | = | Moderate overlap (5% to <10%) |
| 10 | = | High overlap (10% to <15%) |
| 28 | = | Very High overlap (≥15%) |

| | Al-Saud 2018 | Burgess 2019 | Chmielewksa 2017 | Dai 2019 | De Silva 2020 | EFSA Panel 2019 | Ierodiakonou 2016 | Larson 2017 | Scarpone 2023 | Smith 2016 |
|---|---|---|---|---|---|---|---|---|---|---|
| Burgess 2019 | 66,7% | | | | | | | | | |
| Chmielewksa 2017 | 16,7% | 11,1% | | | | | | | | |
| Dai 2019 | 75,0% | 70,0% | 12,5% | | | | | | | |
| De Silva 2020 | 71,4% | 66,7% | 16,7% | 75,0% | | | | | | |
| EFSA Panel 2019 | 38,5% | 50,0% | 8,3% | 42,9% | 38,5% | | | | | |
| Ierodiakonou 2016 | 7,9% | 12,8% | 2,9% | 13,2% | 10,8% | 9,3% | | | | |
| Larson 2017 | 14,3% | 22,2% | 0,0% | 25,0% | 33,3% | 16,7% | 5,7% | | | |
| Scarpone 2023 | 25,0% | 27,8% | 7,1% | 37,5% | 25,0% | 30,0% | 8,9% | 6,7% | | |
| Smith 2016 | 16,7% | 11,1% | 100,0% | 12,5% | 16,7% | 8,3% | 2,9% | 0,0% | 7,1% | |
| Waidyatillake 2018 | 33,3% | 22,2% | 0,0% | 25,0% | 14,3% | 16,7% | 0,0% | 0,0% | 14,3% | 0,0% |

### eTable 1. PRIOR checklist (preferred reporting items for overviews of reviews)

| Section topic | Item No | Item | Location where item is reported |
|---|---|---|---|
| **Title** | | | |
| Title | 1 | Identify the report as an overview of reviews. | Cover page |
| **Abstract** | | | |
| Abstract | 2 | Provide a comprehensive and accurate summary of the purpose, methods, and results of the overview of reviews. | p.4 |
| **Introduction** | | | |
| Rationale | 3 | Describe the rationale for conducting the overview of reviews in the context of existing knowledge. | p.6 |
| Objectives | 4 | Provide an explicit statement of the objective(s) or question(s) addressed by the overview of reviews. | p.6 |
| **Methods** | | | |
| Eligibility criteria | 5a | Specify the inclusion and exclusion criteria for the overview of reviews. If supplemental primary studies were included, this should be stated, with a rationale. | p.7 |
| | 5b | Specify the definition of “systematic review” as used in the inclusion criteria for the overview of reviews. | p.8 |
| Information sources | 6 | Specify all databases, registers, websites, organisations, reference lists, and other sources searched or consulted to identify systematic reviews and supplemental primary studies (if included). Specify the date when each source was last searched or consulted. | p.8 |
| Search strategy | 7 | Present the full search strategies for all databases, registers and websites, such that they could be reproduced. Describe any search filters and limits applied. | eAppendix 2 |
| Selection process | 8a | Describe the methods used to decide whether a systematic review or supplemental primary study (if included) met the inclusion criteria of the overview of reviews. | p.8 |
| | 8b | Describe how overlap in the populations, interventions, comparators, and/or outcomes of systematic reviews was identified and managed during study selection. | p.8 |
| Data collection process | 9a | Describe the methods used to collect data from reports. | p.8 |
| | 9b | If applicable, describe the methods used to identify and manage primary study overlap at the level of the comparison and outcome during data collection. For each outcome, specify the method used to illustrate and/or quantify the degree of primary study overlap across systematic reviews. | p.8-9 |

| Section topic | Item No | Item | Location where item is reported |
|---|---|---|---|
| | 9c | If applicable, specify the methods used to manage discrepant data across systematic reviews during data collection. | p.8 |
| Data items | 10 | List and define all variables and outcomes for which data were sought. Describe any assumptions made and/or measures taken to identify and clarify missing or unclear information. | p.8 |
| Risk of bias assessment | 11a | Describe the methods used to assess risk of bias or methodological quality of the included systematic reviews. | p.9 |
| | 11b | Describe the methods used to collect data on (from the systematic reviews) and/or assess the risk of bias of the primary studies included in the systematic reviews. Provide a justification for instances where flawed, incomplete, or missing assessments are identified but not reassessed. | p.9 |
| | 11c | Describe the methods used to assess the risk of bias of supplemental primary studies (if included). | Not applicable |
| Synthesis methods | 12a | Describe the methods used to summarise or synthesise results and provide a rationale for the choice(s). | p.10 |
| | 12b | Describe any methods used to explore possible causes of heterogeneity among results. | p.10 |
| | 12c | Describe any sensitivity analyses conducted to assess the robustness of the synthesised results. | p.10 |
| Reporting bias assessment | 13 | Describe the methods used to collect data on (from the systematic reviews) and/or assess the risk of bias due to missing results in a summary or synthesis (arising from reporting biases at the levels of the systematic reviews, primary studies, and supplemental primary studies, if included). | p.8 |
| Certainty assessment | 14 | Describe the methods used to collect data on (from the systematic reviews) and/or assess certainty (or confidence) in the body of evidence for an outcome. | p.9-10 |
| **Results** | | | |
| Systematic review and supplemental primary study selection | 15a | Describe the results of the search and selection process, including the number of records screened, assessed for eligibility, and included in the overview of reviews, ideally with a flow diagram. | p.10-11 |
| | 15b | Provide a list of studies that might appear to meet the inclusion criteria, but were excluded, with the main reason for exclusion. | Supplement (eTable 2) |
| Characteristics of systematic reviews and supplemental primary studies | 16 | Cite each included systematic review and supplemental primary study (if included) and present its characteristics. | p.12-13/ Supplement (eTable 3) |

| Section topic | Item No | Item | Location where item is reported |
|---|---|---|---|
| Primary study overlap | 17 | Describe the extent of primary study overlap across the included systematic reviews. | p.15-20/ Suplement (eFigure 1, eTable 11) |
| Risk of bias in systematic reviews, primary studies, and supplemental primary studies | 18a | Present assessments of risk of bias or methodological quality for each included systematic review. | p.14/ Supplement (eTables 4-5) |
| | 18b | Present assessments (collected from systematic reviews or assessed anew) of the risk of bias of the primary studies included in the systematic reviews. | p.14/ Supplement (eTables 6-7) |
| | 18c | Present assessments of the risk of bias of supplemental primary studies (if included). | Not applicable |
| Summary or synthesis of results | 19a | For all outcomes, summarise the evidence from the systematic reviews and supplemental primary studies (if included). If meta-analyses were done, present for each the summary estimate and its precision and measures of statistical heterogeneity. If comparing groups, describe the direction of the effect. | p.15-20/ Supplement (eTable 8, eTable 10) |
| | 19b | If meta-analyses were done, present results of all investigations of possible causes of heterogeneity. | Not applicable |
| | 19c | If meta-analyses were done, present results of all sensitivity analyses conducted to assess the robustness of synthesised results. | Not applicable |
| Reporting biases | 20 | Present assessments (collected from systematic reviews and/or assessed anew) of the risk of bias due to missing primary studies, analyses, or results in a summary or synthesis (arising from reporting biases at the levels of the systematic reviews, primary studies, and supplemental primary studies, if included) for each summary or synthesis assessed. | Supplement (eTable 8, eTable 10) |
| Certainty of evidence | 21 | Present assessments (collected or assessed anew) of certainty (or confidence) in the body of evidence for each outcome. | p.14-15/ Supplement (eTables 8-10) |
| **Discussion** | | | |
| Discussion | 22a | Summarise the main findings, including any discrepancies in findings across the included systematic reviews and supplemental primary studies (if included). | p.20-21 |
| | 22b | Provide a general interpretation of the results in the context of other evidence. | p.21-22 |

| Section topic | Item No | Item | Location where item is reported |
|---|---|---|---|
| | 22c | Discuss any limitations of the evidence from systematic reviews, their primary studies, and supplemental primary studies (if included) included in the overview of reviews. Discuss any limitations of the overview of reviews methods used. | p.21 |
| | 22d | Discuss implications for practice, policy, and future research (both systematic reviews and primary research). Consider the relevance of the findings to the end users of the overview of reviews, eg, healthcare providers, policymakers, patients, among others. | p.22 |
| **Other information** | | | |
| Registration and protocol | 23a | Provide registration information for the overview of reviews, including register name and registration number, or state that the overview of reviews was not registered. | p.6 |
| | 23b | Indicate where the overview of reviews protocol can be accessed, or state that a protocol was not prepared. | p.6 |
| | 23c | Describe and explain any amendments to information provided at registration or in the protocol. Indicate the stage of the overview of reviews at which amendments were made. | p.6-7/ Supplement (eAppendix 1) |
| Support | 24 | Describe sources of financial or non-financial support for the overview of reviews, and the role of the funders or sponsors in the overview of reviews. | p.3 |
| Competing interests | 25 | Declare any competing interests of the overview of reviews' authors. | p.2 |
| Author information | 26a | Provide contact information for the corresponding author. | Cover page |
| | 26b | Describe the contributions of individual authors and identify the guarantor of the overview of reviews. | p.2 |
| Availability of data and other materials | 27 | Report which of the following are available, where they can be found, and under which conditions they may be accessed: template data collection forms; data collected from included systematic reviews and supplemental primary studies; analytic code; any other materials used in the overview of reviews. | p.3/ Supplemet |

### eTable 2. Excluded SRs After Full-text Screening and Reasons for Exclusion

| Systematic Review | Reason for exclusion | Note |
|---|---|---|
| Anagnostou et al. 2021 | Wrong study design | No systematic review |
| Brough et al. 2021 | Wrong study design | No systematic review |
| Calamelli et al. 2018 | Wrong study design | No systematic review |
| Chiale et al. 2021 | Wrong study design | Outcome of the included RCTs do not match with our prespecified PICOTS criteria (outcomes: Body Mass Index, weight, height) |
| De Silva et al. 2014 | Wrong study design | No RCT on early introduction included in systematic review |
| Dogaru et al. 2014 | Wrong intervention | Role of breastfeeding |
| Garcia-Marcos et al. 2013 | Wrong study design | No RCT included in systematic review |
| Garcia-Larsen et al. 2018 | Wrong intervention | No RCT on early introduction included in systematic review |
| Golpanian et al. 2020 | Wrong study design | No systematic review |
| Güngör et al. 2019 | Wrong intervention | Milk feeding practices |
| Halken et al. 2004 | Wrong study design | No systematic review |
| Host et al. 2008 | Wrong study design | No systematic review |
| Hosseini et al. 2017 | Wrong study design | No RCT included in systematic review |
| Kremmyda et al. 2011 | Wrong intervention | No RCTs on early introduction |
| Lutter et al. 2021 | Wrong study design | No systematic review |
| Lv et al. 2014 | Wrong study design | No RCT included in systematic review |
| McGowan et al. 2014 | Wrong study design | Comment on de Silva et al. 2014 |
| Moustaki et al. 2021 | Wrong study design | No systematic review |
| Nurmatov et al. 2011 | Wrong study design | No RCT included in systematic review |
| Nuzzi et al. 2022 | Wrong intervention | Intervention of the included RCTs do not match with our prespecified PICOTS criteria (intervention: vitamin D or omega-3 fatty acid supplementation, no timing of CF) |
| Obaggy et al. 2019 | Wrong study design | No RCT included in systematic review regarding timing of complementary feeding |
| Oykhman et al. 2022 | Wrong intervention | Intervention of the included RCTs do not match with our prespecified PICOTS criteria (intervention: complete avoidance, only delayed introduction of CF) |
| Robison et al. 2010 | Wrong study design | No RCT included in systematic review |
| Schroer et al. 2021 | Wrong study design | No systematic review |
| Seyedrezazadeh et al. 2014 | Wrong study design | No RCT included in systematic review |

| Systematic Review | Reason for exclusion | Note |
|---|---|---|
| Szajewska et al. 2011 | Wrong study design | No systematic review |
| Thompson et al. 2010 | Wrong intervention | No RCT on early introduction included in systematic review |
| Trivillin et al. 2022 | Wrong intervention | Comparison of the included RCTs do not match with our prespecified PICOTS criteria (no comparison between early vs. late introduction) |
| Ulfman et al. 2022 | Wrong study design | No systematic review |
| West et al. 2017 | Wrong study design | No systematic review |
| Yuan et al. 2020 | Wrong intervention, wrong comparator | Comparison and outcome of the included RCTs do not match with our prespecified PICOTS criteria (intervention: formula feeding only, comparator: not after 6 months of age) |
| Zhang et al. 2020 | Wrong study design | No RCT included in systematic review |
| Zhang et al. 2017 | Wrong intervention | Included RCT on maternal intervention |

**Note.** The table lists the excluded articles with at least one reason for exclusion but may not reflect all possible reasons.

RCT=Randomised controlled trial

### eTable 3. Characteristics of Included Systematic Reviews

| Author, Year | Design | Databases and additional sources searched | Search period, Date of last search update | Included studies (participants) | Search restrictions | Participant characteristics | Interventions addressed | Comparators | Outcomes |
|---|---|---|---|---|---|---|---|---|---|
| **Al-Saud 2018** [2] | Systematic review and meta-analysis | MEDLINE,<br>EMBASE,<br>Cochrane Central Register of Controlled Trials,<br>Meta Register,<br>OpenGREY Repository,<br>Conference abstracts | Initially up to November 2016, updated in March 2017 | 6 RCTs (3032) | Studies on human subjects, no language restrictions | Infants | Early (as early as 3 months of age) introduction of egg to the diet | Exclusive breastfeeding until 6 months of age or placebo | Primary:<br>Egg allergy diagnosed by oral food challenge<br>Secondary:<br>Food sensitisation (presence of allergen specific IgE) confirmed by a positive<br>allergy skin prick test or a positive ImmunoCAP,<br>diagnosis of asthma, allergic rhinitis, eczema, anaphylaxis |
| **Burgess 2019** [3] | Systematic review and meta-analysis | PubMed,<br>EMBASE,<br>reference lists of included studies, trial registries | Up to 20th May 2016,<br>updated on 15th February 2017 | 8 RCTs, 16 cohort studies, 1 case-control study (7 RCTs and 9 cohort studies included in MA)[a] | Humans studies are eligible but not animal studies | Humans from general and high-risk populations (not specified but inferred: children 0-3 years) | Timing of introduction to complementary solid food or specific allergenic foods | Delayed or no introduction (not explicitly stated, protocol: any comparison with infants not introduced at that age) | Food sensitisation determined by skin prick test or food-specific IgE, food<br>allergy determined by food challenge or diagnosed by a physician |
| **Chmielewska 2017** [4] | Systematic review | MEDLINE,<br>EMBASE,<br>Cochrane Library,<br>Web of Science, CINAHL,<br>Contacting researchers in the field, proceedings from scientific meetings | Up to July 2015, additional search of MEDLINE and EMBASE in March 2016 | 2 RCTs, 4 cohort studies, 1 case-control study (8853) | No restrictions by either date or language | Infants at population risk or increased risk of developing a wheat allergy | Consumption of wheat- or gluten-containing products of any type (cereals, flour or any other food items containing gluten) | Placebo or no intervention/ no exposure | Incidence of wheat allergy or sensitisation (increased wheat-specific IgE and/or positive skin-prick test) |
| **Dai 2020** [5] | Systematic review and meta-analysis | PubMed, Cochrane Library, China national knowledge infrastructure (CNKI); Wanfang | Up to 31th December 2019 | 8 RCTs (4112) | No restriction on study types;<br>Languages limited | Infants at population risk and increased risk of developing allergy | Late consumption of any complementary foods (after 6 months of age) | Early consumption of any complementary foods (before 6 months of age) | Allergic diseases (egg allergy, peanut allergy, cow's milk protein allergy) |

| Author, Year | Design | Databases and additional sources searched | Search period, Date of last search update | Included studies (participants) | Search restrictions | Participant characteristics | Interventions addressed | Comparators | Outcomes |
|---|---|---|---|---|---|---|---|---|---|
| **Dai 2020** [5] | | Data database; Reference lists of included trials | | | to Chinese and English | | | | |
| **De Silva 2020** [6] | Systematic review | MEDLINE, EMBASE, Cochrane Library, ISI Web of Science, CINAHL, Science Citation Index and Social Sciences Citation Index, WHOLIS, PAHO, TRIP, WHO ICTRP, US National Institutes of Health Ongoing Trials Register (Clinicaltrials.gov, NIH web), Trial registries, reference lists of identified studies, discussion with experts in the fields | Up to 31th October 2019 | 41 RCTs, 5 cohort studies[a] | No language or geographical restrictions | Infants (up to 1 year old), children (13 months to 17 years), and/or adults (18+ years) with or without an increased risk for developing allergic disease and with or without any sensitisation or atopic manifestations | Any intervention to prevent the development of new cases of immediate-onset food allergy (protocol: includes dietary initiatives and skin barrier initiatives) | Any independent, concurrently sampled group(s) with or without a placebo, intervention, or combination of interventions | New cases of immediate-onset food allergy, defined as a reproducible adverse response to a food protein within hours caused by an immunologic reaction |
| **EFSA Panel 2019** [7] | Systematic review | PubMed, Cochrane Library, Web of Science Core Collection, Reference lists of SRs, grey literature and included primary studies, NTIS, the System for Information on Grey Literature in Europe, CAB Abstracts, Open Access Theses and Dissertations, the US National Guideline Clearinghouse | Up to October 2018 | Prospective studies:<br>13 RCTs,<br>107 prospective cohort studies,<br>9 nested case-control studies,<br>2 pooled analyses of prospective studies[a]<br>Retrospective studies:<br>12 cross-sectional baseline analysis of 9 otherwise prospective studies,<br>29 papers on cross- | No language limits | All population groups, males and females: generally healthy term infants, preterm infants, infants not older than 12 months of age at introduction of CFs | Timing of introduction to CFs | Group alike in terms of the type of initial feeding (breast milk or breast-milk substitutes) and the only important difference being the time at which CF is introduced | Allergy (and overweight and obesity, DM type I and II, risk factors of CVD, coeliac disease, dental health, renal function, gastrointestinal infections, respiratory tract infections etc.) |

| Author, Year | Design | Databases and additional sources searched | Search period, Date of last search update | Included studies (participants) | Search restrictions | Participant characteristics | Interventions addressed | Comparators | Outcomes |
|---|---|---|---|---|---|---|---|---|---|
| **EFSA Panel 2019** [7] | | | | sectional studies, 37 papers on case-control studies, 3 papers on retrospective cohort studies, 1 paper on a prospective cohort study in which the timing of introduction of CFs was assessed after the outcome | | | | | |
| **Ierodiakonou 2016** [8] | Systematic review and meta-analysis | MEDLINE, EMBASE, Web of Science, The Cochrane Library (CENTRAL), LILACS, http://apps.who.int/trialsearch, bibliography of eligible studies, Open Grey | 25th July 2013, updated on 8th March 2016 | Allergic outcomes: 24 intervention trials (13298), 69 observational studies (142103) Autoimmune diseases: 5 intervention trials (5623), 48 observational studies (63576) | No language restrictions | Infants between birth and the end of their 12th post-partum month | Early timing of introduction of allergenic food: cow's milk, egg, fish, crustacean shellfish, tree nuts, wheat, peanuts and soybeans | Delayed or standard introduction of allergenic foods | Allergic, autoimmune disease or allergic sensitisation: Asthma/ wheeze, eczema, allergic rhinitis, food allergy (a reproducible hypersensitivity reaction to a food), allergic sensitisation (the presence of specific IgE to an allergen), type 1 DM, celiac disease, IBD, autoimmune thyroid disease, juvenile rheumatoid arthritis, psoriasis, and vitiligo |
| **Larson 2017** [9] | Systematic review | CINAHL, Medline, PubMed, Science Direct, Web of Science | Not reported | 2 RCTs, 9 prospective cohort studies, 1 case-control study, 2 cross-sectional studies; 14 in total[a] | Primary research articles published in English, with human subjects | Infants | Delaying introduction of potentially allergenic foods until after 12 months of age | Introduction of potentially allergenic foods prior to 12 months | Development of food allergies |
| **Scarpone 2023** [10] | Systematic review and meta-analysis | MEDLINE, Embase, CENTRAL, reference | Initial search on 11th May 2021, updated on 28th | 23 trials (13794); 12 ongoing studies | No language restrictions, RCTs | Infants enrolled from birth to 12 months of age | Earlier allergenic food introduction (milk, egg, fish, shellfish, tree nuts, | Later allergenic food introduction; different doses or types | Primary efficacy outcome: |

| Author, Year | Design | Databases and additional sources searched | Search period, Date of last search update | Included studies (participants) | Search restrictions | Participant characteristics | Interventions addressed | Comparators | Outcomes |
|---|---|---|---|---|---|---|---|---|---|
| **Scarpone 2023** [10] | | lists of relevant included studies | June 2022 and 29th December 2022 | (16765 intended participants) | | | wheat, peanuts, and soya) during the first year of life | of exposures; breastfeeding or breastmilk, amino acid formula, other low-allergen exposures, or standard care | immunoglobulin E (IgE)-mediated food allergy at age 1 to 5 years assessed by double-blind, placebo-controlled food challenge; open food challenge; medical diagnosis; or parental report at the closest reported time point to age 3 years<br><u>primary safety outcome:</u> withdrawal from study intervention<br><u>secondary outcomes:</u> allergenic sensizitation to any food (SPT and/or allergen-specific IgE); allergy and allergenic sensitisation to specific foods |
| **Smith 2016** [11] | Systematic review and meta-analysis | Cochrane Pregnancy and Childbirth Group's Trials Register (contains trials identified from CENTRAL, MEDLINE, Embase, CINAHL, handsearches of 30 journals and conference proceedings, awareness alerts for further 44 journals and monthly BioMed Central email alerts), reference lists of all relevant retrieved papers | Up to 1st March 2016 | 11 randomised or quasi-randomised controlled trials (2542) | No language, geographic or date restrictions | Healthy breastfeeding full-term (37-42 months of gestation; (singleton or multiple births)) infants up to the age of six months, or mothers of these infants | Non-exclusive breastfeeding infants (artificial milk, glucose, water, foods)<br><br>Breastfeeding with any additional food or fluids (once or more) in the first six months | Exclusive breastfeeding infants | <u>Primary</u>: duration of breastfeeding, incidence of infant morbidity (e.g. asthma, eczema, GI infection), infant mortality (at discharge, 28 days, or one year), physiological jaundice;<br><u>Secondary</u>: weight, growth, development, duration of hospital stays, confidence in breastfeeding, maximum serum bilirubin levels, phototherapy |

| Author, Year | Design | Databases and additional sources searched | Search period, Date of last search update | Included studies (participants) | Search restrictions | Participant characteristics | Interventions addressed | Comparators | Outcomes |
|---|---|---|---|---|---|---|---|---|---|
| **Waidyatillake 2018** [12] | Systematic review and meta-analysis | PubMed, EMBASE, trial registries (Australian and New Zealand, European, Japanese), citation alerts, reference lists of included studies | Up to 18th February 2017 | 2 RCTs, 11 cohort studies, 2 case-control studies, 1 cross-sectional studies[a] | English-language, human studies | Infants | Timing of solid food introduction (allergenic or non-allergenic) | Non-exposed group (e.g. infants not introduced to solid food by a certain age), if not available comparison depending on the age at introduction | Diagnosis of eczema |

[a] Number of participants were not reported in the systematic review

### eTable 4. Methodological Quality of the Included Systematic Reviews (AMSTAR-2)

| | AMSTAR-2 item | | | | | | | | | | | | | | | | | |
|---|---|---|---|---|---|---|---|---|---|---|---|---|---|---|---|---|---|---|
| | **1** | **2†** | **3** | **4†** | **5** | **6** | **7†** | **8** | **9a†** | **9b†** | **10** | **11a†** | **11b†** | **12** | **13†** | **14** | **15†** | **16** |
| **Author (Year)** | | | | | | | | | | | | | | | | | | |
| **Al-Saud et al.** (2018) | Yes | Yes | No | PY | Yes | Yes | No | PY | PY | n.a. | No | Yes | n.a. | No | No | Yes | No | Yes |
| **Burgess et al.** (2019) | Yes | PY | Yes | No | Yes | Yes | No | PY | Yes | PY | No | No | Yes | No | No | No | No | Yes |
| **Chmielewska et al.** (2017) | Yes | PY | No | PY | Yes | Yes | Yes | No | Yes | PY | No | n.a. | n.a. | n.a. | Yes | Yes | n.a. | Yes |
| **Dai et al.** (2021) | Yes | No | No | PY | Yes | No | No | No | PY | n.a. | No | No | n.a. | Yes | Yes | Yes | No | Yes |
| **De Silva et al.** (2020) | Yes | Yes | Yes | PY | Yes | Yes | Yes | Yes | Yes | Yes | Yes | n.a. | n.a. | n.a. | Yes | Yes | n.a. | Yes |
| **EFSA Panel et al.** (2019) | Yes | PY | Yes | No | No | Yes | Yes | PY | Yes | Yes | No | Yes | Yes | Yes | Yes | Yes | Yes | No |
| **Ierodiakonou et al.** (2016) | Yes | Yes | Yes | Yes | Yes | Yes | No | PY | PY | PY | No | Yes | Yes | Yes | Yes | Yes | Yes | Yes |
| **Larson et al.** (2017) | Yes | No | No | PY | No | No | No | No | No | No | No | n.a. | n.a. | n.a. | No | No | n.a. | Yes |
| **Scarpone et al.** (2023) | Yes | Yes | No | PY | Yes | Yes | No | PY | Yes | n.a. | Yes | Yes | n.a. | Yes | Yes | Yes | Yes | Yes |
| **Smith et al.** (2016) | Yes | Yes | No | Yes | Yes | Yes | Yes | Yes | Yes | n.a. | Yes | No | n.a. | No | Yes | No | Yes | Yes |
| **Waidyatillake et al.** (2018) | Yes | PY | Yes | No | Yes | Yes | NI | PY | Yes | PY | No | No | No | No | Yes | Yes | No | Yes |

Abbreviations: n.a.=not applicable; PY=partial yes, NI=no information; †indicates critical domain=flaw item

### eTable 5. Risk of Bias in the Included Systematic Reviews (ROBIS)

| | Phase 2 | | | | Phase 3 |
|---|---|---|---|---|---|
| **Review** | **1. STUDY ELIGIBILITY CRITERIA** | **2. IDENTIFICATION AND SELECTION OF STUDIES** | **3. DATA COLLECTION AND STUDY APPRAISAL** | **4. SYNTHESIS AND FIND-INGS** | **RISK OF BIAS IN THE RE-VIEW** |
| **Author(Date)** | | | | | |
| **Al-Saud et al.** (2018) | High risk | High risk | Low risk | High risk | High risk |
| **Burgess et al.** (2019) | High risk | High risk | Low risk | Low risk | High risk |
| **Chmielewska et al.** (2017) | High risk | Low risk | Unclear risk | High risk | High risk |
| **Dai et al.** (2021) | High risk | Low risk | Low risk | High risk | High risk |
| **De Silva et al.** (2020) | Low risk | High risk | Low risk | High risk | High risk |
| **EFSA Panel et al.** (2019) | High risk | High risk | Low risk | High risk | High risk |
| **Ierodiakonou et al.** (2016) | Low risk | Low risk | Low risk | Low risk | Low risk |
| **Larson et al.** (2017) | High risk | High risk | High risk | High risk | High risk |
| **Scarpone et al.** (2023) | Low risk | Low risk | Unclear risk | Unclear risk | Low risk |
| **Smith et al.** (2016) | High risk | Low risk | Low risk | High risk | High risk |
| **Waidyatillake et al.** (2018) | High risk | High risk | Low risk | High risk | High risk |

### eTable 6. Risk of Bias of Primary Studies within Systematic Reviews

| Primary review | Study ID | Outcome | Risk of bias | | | | | |
|---|---|---|---|---|---|---|---|---|
| | | | D1 | D2 | D3 | D4 | D5 | Overall risk of bias |
| **Al Saud et al. 2018** | Bellach 2017 | Risk of hen's egg allergy | Low | Low | Some concerns | Low | Some concerns | Some concerns |
| | Natsume 2017 | | Low | Low | Low | Low | Some concerns | Some concerns |
| | Palmer 2013 | | Low | Low | Low | Low | Some concerns | Some concerns |
| | Palmer 2017 | | Low | Low | Low | Low | Low | Low |
| | Perkin 2016 | | Some concerns | Low | Some concerns | Some concerns | Low | Some concerns |
| | Tan 2017 | | Low | Low | Some concerns | Low | Low | Some concerns |
| | Palmer 2017 | Risk of eczema | Low | Low | Some concerns | Low | Some concerns | Some concerns |
| | Tan 2017 | | Low | Low | Some concerns | Low | Low | Some concerns |
| **Burgess et al. 2019** | Perkin 2016 | Risk of food allergy | Some concerns | Low | Some concerns | High | Low | High |
| | Bellach 2017 | Risk of egg allergy | Low | Low | Some concerns | Low | Some concerns | Some concerns |
| | Halpern 1973 | | High | High | High | Low | Some concerns | High |
| | Palmer 2013 | | Low | Low | Low | Low | Some concerns | Some concerns |
| | Palmer 2017 | | Low | Low | Low | Low | Low | Low |
| | Perkin 2016 | | Some concerns | Low | Some concerns | Some concerns | Low | Some concerns |
| | Natsume 2017 | | Low | Low | Low | Low | Some concerns | Some concerns |
| | Tan 2017 | | Low | Low | Some concerns | Low | Low | Some concerns |
| | Du Toit 2015 | Risk of peanut allergy in the group with initial positive SPT | Some concerns | Some concerns | Low | Some concerns | Low | Some concerns |
| | | Risk of peanut allergy in the group with initial negative SPT | Some concerns | Some concerns | Low | Low | Low | Some concerns |
| | Perkin 2016 | Risk of peanut allergy | Some concerns | Low | Some concerns | High | Low | High |
| **Chmielewska et al. 2017** | Perkin 2016 | Risk of wheat allergy | Some concerns | Some concerns | Low | Some concerns | Low | Low |
| **Dai et al. 2019** | Perkin 2016 | Risk of egg allergy | Some concerns | Low | Some concerns | Some concerns | Low | Some concerns |
| | Natsume 2017 | | Low | Low | Low | Low | Some concerns | Some concerns |
| | Tan 2017 | | Low | Low | Some concerns | Low | Low | Some concerns |
| | Bellach 2017 | | Low | Low | Some concerns | Low | Some concerns | Some concerns |
| | Palmer 2017 | | Low | Low | Low | Low | Low | Low |
| | Palmer 2013 | | Low | Low | Low | Low | Some concerns | Some concerns |
| | Perkin 2016 | Risk of peanut allergy | Some concerns | Low | Some concerns | High | Low | High |

| Primary review | Study ID | Outcome | Risk of bias | | | | | |
|---|---|---|---|---|---|---|---|---|
| | | | D1 | D2 | D3 | D4 | D5 | Overall risk of bias |
| **Dai et al. 2019** | Du Toit 2015 (with initial positive SPT) | | Some concerns | Some concerns | Low | Some concerns | Low | Some concerns |
| | Du Toit 2015 (with initial negative SPT) | | Some concerns | Some concerns | Low | Low | Low | Some concerns |
| | Perkin 2016 | Risk of cow's milk protein allergy | Low | Low | some concerns | Low | some concerns | some concerns |
| | Lowe 2011 | | Low | High | High | some concerns | some concerns | High |
| **De Silva et al. 2020** | Perkin 2016 | Cumulative food allergy prevalence 1-3 years by introduction of multiple foods | Some concerns | Low | Some concerns | High | Low | High |
| | Natsume 2017 | Risk of egg allergy by introduction of cooked egg from 6 months of age | Low | Low | Some concerns | Low | Some concerns | Some concerns |
| | Bellach 2017 | Risk of egg allergy by introduction of raw/pasteurised raw egg powder from 6 month of age | Low | Low | Low | Low | Some concerns | Some concerns |
| | Palmer 2013 | | Low | Low | Low | Low | Some concerns | Some concerns |
| | Palmer 2017 | | Low | Low | Low | Low | Low | Low |
| | Du Toit 2015 | Incidence of peanut allergy at 5 years in the group with initial positive SPT | Some concerns | Some concerns | Low | Some concerns | Low | Some concerns |
| | | Incidence of peanut allergy at 5 years in the group with initial negative SPT | Some concerns | Some concerns | Low | Low | Low | Some concerns |
| **EFSA Panel 2019** | Perkin 2016 | Risk of symptomatic food allergy by introduction of CF in general | Some concerns | Low | Some concerns | High | Low | High |
| | Bellach 2017 | | Low | Low | Some concerns | Low | Some concerns | Some concerns |

| Primary review | Study ID | Outcome | Risk of bias | | | | | |
|---|---|---|---|---|---|---|---|---|
| | | | D1 | D2 | D3 | D4 | D5 | Overall risk of bias |
| **EFSA Panel 2019** | Palmer 2013 | Risk of symptomatic food allergy by introduction of egg | Low | Low | Some concerns | Low | Some concerns | Some concerns |
| | Palmer 2017 | | Low | Low | Low | Low | Low | Low |
| | Perkin 2016 | | Some concerns | Low | Some concerns | Some concerns | Low | Some concerns |
| | Tan 2017 | | Low | Low | Some concerns | Low | Low | Some concerns |
| | Perkin 2016 | Risk of symptomatic food allergy by introduction of cereals | Some concerns | Low | Some concerns | Some concerns | Low | Some concerns |
| | Perkin 2016 | Risk of symptomatic food allergy by introduction of peanut | Some concerns | Low | Some concerns | High | Low | High |
| | Palmer 2017 | Risk of eczema by introduction of egg | Low | Low | Some concerns | Low | Some concerns | Some concerns |
| | Tan 2017 | | Low | Low | Some concerns | Low | Low | Some concerns |
| | Perkin 2016 | Risk of asthma-like symptoms[a] (intervention: timing of CF in general) | Some concerns | Low | Some concerns | High | Low | High |
| | Palmer 2017 | Risk of asthma-like symptoms[a] (intervention: timing of CF in egg) | Low | Low | Low | Low | Low | Low |
| | Perkin 2016 | Odds of developing allergic rhinitis[e] | NA | NA | NA | NA | NA | NA |
| | Perkin 2016 | Atopic diseases (intervention: timing of CF in general) | Some concerns | Low | Some concerns | High | Low | High |
| | Halpern 1973 | Atopic diseases (intervention: timing of egg yolk introduction) | High | High | High | Low | Some concerns | High |
| **Ierodiakonou et al. 2016** | Halmerbauer 2002 and 2003 | Risk of food allergy | Low | Low | Low | Low | Some concerns | Some concerns |
| | Hide 1994 and 1996; Arshad 1992, 2003 and 2007; Scott 2012 | | Low | High | Some concerns | Low | Some concerns | High |
| | Lowe 2011 | | Low | High | High | Some concerns | Some concerns | High |
| | Perkin 2016 | | Some concerns | Low | Some concerns | High | Low | High |

| Primary review | Study ID | Outcome | Risk of bias | | | | | |
|---|---|---|---|---|---|---|---|---|
| | | | D1 | D2 | D3 | D4 | D5 | Overall risk of bias |
| **Ierodiakonou et al. 2016** | Zeiger 1989, 1992 and 1994 | | Some concerns | High | Some concerns | Some concerns | Some concerns | High |
| | Zhou, 2014 | | Some concerns | Some concerns | Low | Low | Some concerns | Some concerns |
| | Bellach 2015[4] | Risk of egg allergy | Some concerns | High | High | Some concerns | Some concerns | High |
| | Natsume 2016[4] | | Some concerns | Low | High | Low | Some concerns | High |
| | Palmer 2013 | | Low | Low | Low | Low | Some concerns | Some concerns |
| | Halpern 1973 | | High | High | High | Low | Some concerns | High |
| | Perkin 2016 | | Some concerns | low | Some concerns | Some concerns | Low | Some concerns |
| | Tan 2016[4] | | Some concerns | High | High | High | Some concerns | High |
| | Du Toit 2015 | Risk of (pea-)nut allergy | Some concerns | Some concerns | Low | Some concerns | Low | Some concerns |
| | Halmerbauer 2002 and 2003 | | Low | Low | Low | Low | Some concerns | Some concerns |
| | Hide 1994 and 1996; Arshad 1992, 2003 and 2007; Scott 2012 | | Low | Low | Some concerns | Low | Some concerns | Some concerns |
| | Zeiger 1989, 1992 and 1994 | | Some concerns | High | Some concerns | Low | Some concerns | High |
| | Perkin, 2016 | Risk of cow's milk allergy | Some concerns | Low | Some concerns | Some concerns | Low | High |
| | Lowe, 2011 | | Low | High | High | Some concerns | Some concerns | High |
| | Becker 2004; Chan-Yeung 2000/5; Carlsten 2013 | Risk of eczema/atopic dermatitis | Some concerns | High | some concerns | Low | Some concerns | High |
| | Brown 1969 | | Some concerns | High | High | Some concerns | Some concerns | High |
| | Burr 1993; Merrett 1988; Miskelly 1988 | | Some concerns | High | High | Low | Some concerns | High |
| | de Jong 2002 | | Some concerns | High | Some concerns | Low | Some concerns | High |
| | Halmerbauer 2002 and 2003 | | Low | Low | low | Low | Some concerns | Some concerns |
| | Hide 1994 and 1996; Arshad 1992, 2003 and 2007; Scott 2012 | | Low | High | Some concerns | Low | Some concerns | High |
| | Johnstone 1966 | | Some concerns | High | High | High | Some concerns | High |
| | Kjellman 1979 | | Some concerns | Some concerns | Low | Some concerns | Some concerns | Some concerns |
| | Lowe 2011 | | Low | Some concerns | High | Some concerns | Some concerns | High |

| Primary review | Study ID | Outcome | Risk of bias | | | | | |
|---|---|---|---|---|---|---|---|---|
| | | | D1 | D2 | D3 | D4 | D5 | Overall risk of bias |
| **Ierodiakonou et al. 2016** | Matthew 1977 | | High | High | High | Some concerns | Some concerns | High |
| | Shao 2006[a] | | Low | Some concerns | Low | Some concerns | Some concerns | Some concerns |
| | Zeiger 1989, 1992 and 1994 | | Some concerns | High | Some concerns | Some concerns | Some concerns | High |
| | Zhou 2014 | | Some concerns | Low | Low | Some concerns | Some concerns | Some concerns |
| | Becker 2004; Chan-Yeung 2000/5; Carlsten 2013 | Risk of asthma[b] | Some concerns | High | Some concerns | Low | Some concerns | High |
| | Brown 1969 | | Some concerns | High | High | Some concerns | Some concerns | High |
| | de Jong 2002 | | Some concerns | High | Some concerns | Low | Some concerns | High |
| | Du Toit 2016 | | High | Some concerns | Low | Low | Low | High |
| | Halmerbauer 2002 and 2003 | | Low | Low | Low | Some concerns | Some concerns | Some concerns |
| | Hide 1994 and 1996; Arshad 1992, 2003 and 2007; Scott 2012 | | Low | High | Some concerns | Low | Some concerns | High |
| | Johnstone 1966 | | Some concerns | High | Some concerns | High | Some concerns | High |
| | Kjellman 1979 | | Some concerns | High | Low | High | Some concerns | High |
| | Lowe 2011 | | Low | Some concerns | High | Some concerns | Some concerns | High |
| | Perkin 2016 | | Some concerns | Low | High | Some concerns | Low | High |
| | Zeiger 1989, 1992 and 1994 | | Some concerns | High | Some concerns | Some concerns | Some concerns | High |
| | Becker 2004; Chan-Yeung 2000/5; Carlsten 2013 | Risk of allergic rhinitis | Some concerns | High | Some concerns | Low | Some concerns | High |
| | Brown 1969 | | Some concerns | High | High | High | Some concerns | High |
| | Burr 1993; Merrett 1988; Miskelly 1988 | | Some concerns | High | High | Low | Some concerns | High |
| | de Jong 2002 | | Some concerns | High | Some concerns | Low | Some concerns | High |
| | Du Toit 2016 | | High | some concerns | Low | Low | Low | High |
| | Hide 1994 and 1996; Arshad 1992, 2003 and 2007; Scott 2012 | | Low | High | Low | Some concerns | Some concerns | High |
| | Johnstone 1966 | | Some concerns | High | High | Some concerns | Some concerns | High |
| | Kjellman 1979 | | Some concerns | Some concerns | Low | Some concerns | Some concerns | Some concerns |

| Primary review | Study ID | Outcome | Risk of bias | | | | | |
|---|---|---|---|---|---|---|---|---|
| | | | D1 | D2 | D3 | D4 | D5 | Overall risk of bias |
| **Ierodiakonou et al. 2016** | Lowe 2011 | | Low | Some concerns | High | Some concerns | Some concerns | High |
| | Zeiger 1989, 1992 and 1994 | | Some concerns | High | Some concerns | Some concerns | Some concerns | High |
| **Larson et al. 2017** | Palmer 2013 | Risk of egg allergy | Low | Low | Low | Low | Some concerns | Some concerns |
| | Du Toit 2015 | Risk of peanut allergy in the group with initial positive SPT | Some concerns | Some concerns | Low | Some concerns | Low | Some concerns |
| | | Risk of peanut allergy in the group with initial negative SPT | Some concerns | Some concerns | Low | Low | Low | Some concerns |
| **Scarpone et al. 2023** | Nishimura 2022 | Risk of any food allergy (intervention: introduction of *multiple allergenic foods*) | Low | Low | Low | Low | Low | Low |
| | Perkin 2016 | | Low | Low | Low | Some concerns | low | Some concerns |
| | Quake 2022 | | Some concerns | Low | Low | Some concerns | Some concerns | high |
| | Skjerven 2022 | | Low | Low | Low | Low | Low | Low |
| | Nishimura 2022 | Risk of any food allergy (intervention: early introduction of *egg*) | Low | Low | Low | Low | Low | Low |
| | Perkin 2016 | | Low | Low | Low | Some concerns | Low | Some concerns |
| | Quake 2022 | | Some concerns | Low | Low | Some concerns | Some concerns | High |
| | Skjerven 2022 | | Low | Low | Low | Low | Low | Low |
| | Du Toit 2018 | Risk of any food allergy (intervention: early introduction of *peanut*) | Low | Low | Low | High | Some concerns | High |
| | Nishimura 2022 | | Low | Low | Low | Low | Low | Low |
| | Perkin 2016 | | Low | Low | Low | Some concerns | low | Some concerns |
| | Quake 2022 | | Some concerns | Low | Low | Some concerns | Some concerns | High |
| | Skjerven 2022 | | Low | Low | Low | Low | Low | Low |
| | Lowe 2011 | Risk of any food allergy (intervention: early introduction of *cow's milk*) | Some concerns | Low | Low | Low | Low | Low |
| | Nishimura 2022 | | Low | Low | Low | Low | Low | Low |
| | Perkin 2016 | | Low | Low | Low | Some concerns | low | Some concerns |
| | Quake 2022 | | Some concerns | Low | Low | Some concerns | Some concerns | High |
| | Skjerven 2022 | | Low | Low | Low | low | Low | Low |
| | Urashima 2019 | | Low | Low | Low | High | Low | High |
| | Perkin 2016 | Risk of any food allergy (intervention: early introduction of *wheat*) | Low | Low | Low | Some concerns | Low | Some concerns |
| | Skjerven 2022 | | Low | Low | Low | Low | Low | Low |
| | Nishimura 2022 | | Low | Low | Low | Low | Low | Low |
| | Quake 2022 | | Some concerns | Low | Low | Some concerns | Some concerns | High |
| | Nishimura 2022 | Risk of any food allergy (intervention: | Low | Low | Low | Low | Low | Low |
| | Quake 2022 | | Some concerns | Low | Low | Some concerns | Some concerns | High |

| Primary review | Study ID | Outcome | Risk of bias | | | | | |
|---|---|---|---|---|---|---|---|---|
| | | | D1 | D2 | D3 | D4 | D5 | Overall risk of bias |
| **Scarpone et al. 2023** | | early introduction of *soya*) | | | | | | |
| | Nishimura 2022 | Risk of any food allergy (intervention: early introduction of *crustaceans or tree nuts*) | Low | Low | Low | Low | Low | Low |
| | Perkin 2016 | Risk of any food allergy (intervention: early introduction of *fish*) | Low | Low | Low | Some concerns | Low | Some concerns |
| | Quake 2022 | | Some concerns | Low | Low | Some concerns | Some concerns | High |
| | Bellach 2017 | Risk of *egg allergy* (intervention: early introduction of egg) | Low | Low | Some concerns | high | Some concerns | High |
| | Iannotti 2017 | | Low | Low | Low | Low | Some concerns | Some concerns |
| | Natsume 2017 | | Some concerns | Some concerns | Some concerns | Low | Low | High |
| | Nishimura 2022 | | Low | Low | Low | Low | Low | Low |
| | Palmer 2013 | | Low | Low | Low | Low | Low | Low |
| | Palmer 2017 | | Low | Low | Low | Low | Low | Low |
| | Perkin 2016 | | Low | Low | Low | Some concerns | Low | Some concerns |
| | Skjerven 2022 | | Low | Low | Low | Low | Low | Low |
| | Tan 2017 | | Low | Low | Some concerns | Low | Low | Some concerns |
| | Du Toit 2015 | Risk of peanut allergy (intervention: early introduction of peanut) | Low | Low | Low | Some concerns | Low | Some concerns |
| | Nishimura 2022 | | Low | Low | Low | Low | Low | Low |
| | Perkin 2016 | | Low | Low | Low | Some concerns | Low | Some concerns |
| | Skjerven 2022 | | Low | Low | Low | Low | Low | Low |
| | Kjellman 1979 | Risk of cow's milk allergy (intervention: Early introduction of cow's milk) | Some concerns | Low | Low | High | Some concerns | High |
| | Lowe 2011 | | Some concerns | Low | Low | Low | Some concerns | High |
| | Nishimura 2022 | | Low | Low | Low | Low | Low | low |
| | Perkin 2016 | | Low | Low | Low | Some concerns | Low | Some concerns |
| | Skjerven 2022 | | Low | Low | Low | Low | Low | low |
| | Urashima 2019 | | Low | Low | Low | High | Low | High |
| | Perkin 2016 | Risk of allergy to wheat (intervention: early introduction of wheat) | Low | Low | Low | Some concerns | Low | Some concerns |
| | Skjerven 2022 | | Low | Low | Low | Low | Low | Low |
| | Nishimura 2022 | | Low | Low | Low | Low | Low | Low |

| Primary review | Study ID | Outcome | Risk of bias | | | | | |
|---|---|---|---|---|---|---|---|---|
| | | | D1 | D2 | D3 | D4 | D5 | Overall risk of bias |
| **Scarpone et al. 2023** | Nishimura 2022 | Risk of allergy to soya (intervention: early introduction of soya) | Low | Low | Low | Low | Low | Low |
| | Perkin 2016 | Risk of allergy to fish (intervention: early introduction fish) | Low | Low | Low | Some concerns | Low | Some concerns |
| **Smith et al. 2016** | Perkin, 2016 | Risk of food allergy | Some concerns | Low | Some concerns | High | Low | High |
| | Perkin 2016 | Risk of eczema | Some concerns | Low | Some concerns | Low | Low | Some concerns |
| **Waidyatillake et al. 2019** | Palmer 2017 | Risk of eczema | Low | Low | Some concerns | Low | Some concerns | Some concerns |
| | Tan 2017 | | Low | Low | Some concerns | Low | Low | Some concerns |

D1=Bias arising from the randomisation process, D2=Bias due to deviations from the intended interventions, D3=Bias due to missing outcome data, D4=Bias in measurement of the outcome, D5=Bias in selection of the reported result;
[a] The article of this primary study was only available in Chinese. Due to language barriers, the RoB 2.0 assessment could only be performed by one reviewer. [b] The overall risk of bias judgement was performed by the overview authors, except for Scarpone et al.[10].

**eTable 7. Risk of Bias of Primary Studies Within Systematic Reviews Based on Tools Used in the Systematic Reviews for Secondary Outcomes/ Overall Studies**

| Risk of Bias (version 1) | | | | | | | | |
|---|---|---|---|---|---|---|---|---|
| Primary review | Study ID | Outcome | Risk of bias[a] | | | | | |
| | | | D1 | D2 | D3 | D4 | D5 | Overall risk of bias |
| **De Silva et al. 2020** | Bellach 2017 | Bias assessment was conducted for the study as a whole by the review authors | Low | Low | Low | Low | Low | Low |
| | Du Toit 2015, 2016 | | Low | Some concerns | Some concerns | Low | Low | Low |
| | Natsume 2017 | | Low | Low | Low | Low | Low | Low |
| | Palmer 2013 (*egg allergy*) | | Low | Low | high | high | Low | High |
| | Palmer 2013 | | Low | Some concerns | Some concerns | Some concerns | Low | Some concerns |
| | Palmer 2017 | | Low | Low | Some concerns | Low | Low | Low |
| | Perkin 2016 | | Low | Some concerns | Some concerns | Low | Some concerns | Some concerns |
| **Scarpone et al. 2023** | Nishimura 2022 | Allergic sensitisation to *any food* (intervention: *multiple allergenic foods or wheat*) | Low | Low | Low | Low | Low | Low |
| | Perkin 2016 | | Low | Low | Low | Low | Low | Low |
| | Skjerven,2022 | | Low | Low | Low | Low | Low | Low |
| | Du Toit 2018 | Allergic sensitisation *to any food* (intervention: early introduction of *peanut*) | Low | Low | Low | Some concerns | Some concerns | high |
| | Nishimura 2022 | | Low | Low | Low | Low | Low | Low |
| | Perkin 2016 | | Low | Low | Low | Low | Low | Low |
| | Skjerven 2022 | | Low | Low | Low | Low | Low | Low |
| | Nishimura 2022 | Allergic sensitisation to any food (intervention: early intro-duction of *soya*) | Low | Low | Low | Low | Low | Low |
| | Perkin 2016 | Allergic sensitisation to any food (intervention: early intro-duction of *fish*) | Low | Low | Low | Low | Low | Low |
| | Bellach 2017 | Allergic sensitisation *to egg* (intervention: early introduction of *egg*) | Low | Low | Some concerns | Low | Some concerns | high |
| | Makrides 2002 | | Low | Some concerns | high | Low | Some concerns | high |
| | Nishimura 2022 | | Low | Low | Low | Low | Low | Low |
| | Palmer 2013 | | Low | Low | Low | Low | Low | Low |
| | Palmer 2017 | | Low | Low | Low | Low | Low | Low |
| | Perkin 2016 | | Low | Low | Low | Low | Low | Low |
| | Skjerven 2022 | | Low | Low | Low | Low | Low | Low |
| | Tan 2017 | | Low | Low | Low | Low | Some concerns | Some concerns |
| | Du Toit 2018 | | Low | Low | Low | Some concerns | Low | Some concerns |

| Risk of Bias (version 1) | | | | | | | | |
|---|---|---|---|---|---|---|---|---|
| Primary review | Study ID | Outcome | Risk of bias[a] | | | | | |
| | | | D1 | D2 | D3 | D4 | D5 | Overall risk of bias |
| **Scarpone et al. 2023** | Nishimura 2022 | Allergic sensitisation *to peanut* (intervention: early introduction of *peanut*) | Low | Low | Low | Low | Low | Low |
| | Perkin 2016 | | Low | Low | Low | Low | Low | Low |
| | Skjerven 2022 | | Low | Low | Low | Low | Low | Low |
| | de Jong 1998 | Allergic sensitisation to cow's milk (intervention: early introduction of cow's milk) | Low | Low | Low | Low | Some concerns | Some concerns |
| | Kjellman 1979 | | Some concerns | Low | Low | Some concerns | Some concerns | High |
| | Lowe 2011 | | Some concerns | High | Some concerns | Low | Some concerns | High |
| | Nishimura 2022 | | Low | Low | Low | Low | Low | Low |
| | Perkin 2016 | | Low | Low | Low | Low | Low | Low |
| | Skjerven 2022 | | Low | Low | Low | Low | Low | Low |
| | Urashima 2019 | | Low | Low | Low | Low | Low | Low |
| | Perkin 2016 | Allergic sensitisation to wheat (intervention: early introduction of wheat) | Low | Low | Low | Low | Low | Low |
| | Skjerven 2022 | | Low | Low | Low | Low | Low | Low |
| | Nishimura 2022 | | Low | Low | Low | Low | Low | Low |
| | Kjellman 1979 | Allergic sensitisation to soya (intervention: early introduction of soya) | Some concerns | Low | Low | Some concerns | Some concerns | High |
| | Nishimura 2022 | | Low | Low | Low | Low | Low | Low |
| | Perkin 2016 | Allergic sensitisation to fish (intervention: early introduction of fish) | Low | Low | Low | Low | Low | Low |
| | Holl 2020 | Risk of withdrawal from study intervention (intervention: *multiple allergenic foods*) | Some concerns | Low | Low | Low | Some concerns | High |
| | Nishimura 2022 | | Low | Low | Low | Low | Low | Low |
| | Perkin 2016 | | Low | Low | Low | High | Low | High |
| | Quake 2022 | | Some concerns | Low | Low | Some concerns | Some concerns | High |
| | Skjerven 2022 | | Low | Low | Low | High | Low | High |
| | Bellach 2017 | Risk of withdrawal from study intervention (intervention: early introduction of *egg*) | Low | Low | Low | Low | Some concerns | Some concerns |
| | Holl 2020 | | Some concerns | Low | Low | Low | Some concerns | High |
| | Iannotti 2017 | | Low | Low | Low | high | Some concerns | High |
| | Makrides 2002 | | Low | Low | Low | high | Some concerns | High |
| | Natsume 2017 | | Some concerns | Low | Low | Low | Low | Some concerns |
| | Nishimura 2022 | | Low | Low | Low | Low | Low | Low |
| | Palmer 2013 | | Low | Low | Low | Low | Low | Low |
| | Palmer 2017 | | Low | Low | Low | Low | Low | Low |

| Risk of Bias (version 1) | | | | | | | | |
|---|---|---|---|---|---|---|---|---|
| Primary review | Study ID | Outcome | Risk of bias[a] | | | | | |
| | | | D1 | D2 | D3 | D4 | D5 | Overall risk of bias |
| **Scarpone et al. 2023** | Perkin 2016 | | Low | Low | Low | high | Low | High |
| | Quake 2022 | | Some concerns | Low | Low | Some concerns | Some concerns | High |
| | Skjerven 2022 | | Low | Low | Low | high | Low | High |
| | Stewart 2019 | | Low | Low | Low | high | Low | High |
| | Tan 2017 | | Low | Low | Low | Low | Low | Low |
| | Du Toit 2015 | Risk of withdrawal from study intervention (intervention: early introduction of *peanut*) | Low | Low | Low | High | Low | High |
| | Holl 2020 | | Some concerns | Low | Low | Low | Some concerns | High |
| | Nishimura 2022 | | Low | Low | Low | Low | Low | low |
| | Perkin 2016 | | Low | Low | Low | High | Low | High |
| | Quake 2022 | | Some concerns | Low | Low | Some concerns | Some concerns | High |
| | Skjerven 2022 | | Low | Low | Low | High | Low | High |
| | Brown 1969 | Risk of withdrawal from study intervention (intervention: early introduction of *cow's milk*) | Some concerns | Low | Low | High | Some concerns | High |
| | de Jong,1998 | | Low | Low | Low | Low | Some concerns | some concerns |
| | Holl 2020 | | Some concerns | Low | Low | Low | Some concerns | High |
| | Kjellman 1979 | | Some concerns | Low | Low | High | Some concerns | High |
| | Lowe 2011 | | Some concerns | Low | Low | Low | Some concerns | High |
| | Nishimura 2022 | | Low | Low | Low | Low | Low | Low |
| | Perkin 2016 | | Low | Low | Low | High | Low | High |
| | Quake 2022 | | Some concerns | Low | Low | Some concerns | Some concerns | High |
| | Sakihara 2021 | | High | Low | Low | High | Some concerns | High |
| | Skjerven 2022 | | Low | Low | Low | High | Low | High |
| | Urashima 2019 | | Low | Low | Low | High | Low | High |
| | Perkin 2016 | Risk of withdrawal from study intervention (intervention: early introduction of wheat) | Low | Low | Low | High | Low | High |
| | Skjerven 2022 | | Low | Low | Low | High | Low | High |
| | Holl 2020 | | Some concerns | Low | Low | Low | Some concerns | High |
| | Nishimura 2022 | | Low | Low | Low | Low | Low | Low |
| | Quake 2022 | | Some concerns | Low | Low | Some concerns | Some concerns | High |
| | Brown 1969 | Risk of withdrawal from study intervention (intervention: early introduction of *soya*) | Some concerns | Low | Low | High | Some concerns | High |
| | Holl 2020 | | Some concerns | Low | Low | Low | Some concerns | High |
| | Kjellman 1979 | | Some concerns | Low | Low | high | Some concerns | High |
| | Lowe,2011 | | Some concerns | Low | Low | Low | Some concerns | High |
| | Nisjimura 2022 | | Low | Low | Low | Low | Low | Low |
| | Sakihara 2021 | | High | Low | Low | High | Some concerns | High |
| | Perkin 2016 | | Low | Low | Low | High | Low | High |
| | Holl 2020 | | Some concerns | Low | Low | Low | Some concerns | High |

| Risk of Bias (version 1) | | | | | | | | |
|---|---|---|---|---|---|---|---|---|
| Primary review | Study ID | Outcome | Risk of bias[a] | | | | | |
| | | | D1 | D2 | D3 | D4 | D5 | Overall risk of bias |
| **Scarpone et al. 2023** | Quake 2022 | Risk of withdrawal from study intervention (intervention: early introduction of fish) | Some concerns | Low | Low | Some concerns | Some concerns | High |
| | Holl 2020 | Risk of withdrawal from study intervention (intervention: early introduction of crustaceans and nuts) | Some concerns | Low | Low | Low | Some concerns | High |
| | Quake 2022 | | Some concerns | Low | Low | Some concerns | Some concerns | High |

[a]D1=Bias arising from the randomisation process, D2=Bias due to deviations from the intended interventions, D3=Bias due to missing outcome data, D4=Bias in measurement of the outcome, D5=Bias in selection of the reported result

| Risk of Bias (version 1), not result-specific[a] | | | | | | | | | |
|---|---|---|---|---|---|---|---|---|---|
| Primary review | Study ID | Selection Bias | | Risk of bias[a] | | | | | |
| | | Randomisation | Concealment | Performance Bias | Detection Bias | Attrition Bias | Reporting Bias | Other sources of bias | Overall risk of bias |
| **Al-Saud et al. 2016** | Bellach 2015 | Low | Low | Low | High | Low | NA | NA | NA |
| | Palmer 2013 | Low | Unclear | Low | Low | Low | NA | NA | NA |
| | Palmer 2017 | Low | Unclear | Low | Unclear | Low | NA | NA | NA |
| | Perkin 2016 | Low | Low | High | High | Low | NA | NA | NA |
| | Tan 2017 | Low | low | Low | Low | Low | NA | NA | NA |
| **Chmielewska et al. 2017** | Wong 2016 | Yes | Yes | No | No | Yes<br>(60 % at age 15y) | No | No | NA |
| | Perkin 2016 | Yes | Unclear | No | No | Yes (91.3 %) | No | No | NA |
| **EFSA Panel 2016** | Perkin 2016<br>(*Asthma-like symptoms, food sensitisation, WL(H)Z, WAZ, L(H)AZ, HCZ, attained head circumference*) | Definitely low | Definitely low | Probably low | Probably low<br>(except for food sensitisation: definitely low) | Definitely low | Definitely low | Definitely low | Low |
| | Bellach 2017<br>(*Food sensitisation*) | Definitely low | Definitely low | Probably low | Definitely low | Probably low | Definitely low | Definitely low | Low |
| | Palmer 2013<br>(*Food sensitisation*) | Definitely low | Definitely low | Definitely low | Definitely low | Probably low | Definitely low | Definitely low | Low |
| | Palmer 2017<br>(*Asthma-like symptoms, food sensitisation*) | Definitely low | Definitely low | Definitely low | Asthma-like symptoms: Probably high<br><br>Food sensitisation: Definitely low | Definitely low | Definitely low | Probably high | Asthma-like symptoms: Intermediate<br><br>Food sensitisation: Low |
| | Tan 2017<br>(*Food sensitisation*) | Definitely low | Probably high | Probably low | Definitely low | Definitely low | Probably low | Probably low | Low |
| | Cohen 1995 (*WAZ, absolute body gain, L(H)AZ, absolute length gain*) | Definitely high | Probably high | Probably high | Definitely low | Probably low | Definitely low | Probably low | Intermediate |

| Risk of Bias (version 1), not result-specific[a] | | | | | | | | | |
|---|---|---|---|---|---|---|---|---|---|
| Primary review | Study ID | Selection Bias | | Risk of bias[a] | | | | | |
| | | Randomisation | Concealment | Performance Bias | Detection Bias | Attrition Bias | Reporting Bias | Other sources of bias | Overall risk of bias |
| **EFSA Panel 2016** | Mehta 1998 (*WAZ L(H)AZ , attained head circumference*) | Probably high | Probably high | Probably high | Definitely low | Probably low | Probably low | probably high | Intermediate |
| | Dewey 1999 (WAZ, L(H)AZ ) | Definitely high | Probably high | Probably high | Definitely low | Probably low | Definitely low | Probably low | Intermediate |
| | Jonsdottir 2014 (*WAZ, L(H)AZ, HCZ, attained head circumference*) | Definitely low | Definitely low | probably low | Definitely low | Definitely low | Definitely low | Definitely low | Low |

WAZ=Weight-for-age z-score, WL(H)Z=Weight-for-length(height)-z-scores, L(H)AZ=Length(height)-for-age z-scores, HCZ=Head circumference-for-age z-scores,

| Primary review | Risk of Bias assessment individual notation of the authors of the primary reviews | | | | | | | | |
|---|---|---|---|---|---|---|---|---|---|
| | Study ID | Selection bias | | Not assessed | Assessment bias | Attrition | Not assessd | Conflict of interest | Overall risk of bias |
| **Ierodiakonou et al. 2016** | Bellach 2015 | Low | | NA | Low | Unclear | NA | Unclear | Unclear |
| | Becker 2004; Chan-Yeung 2000/5; Carlsten 2013 | Low | | NA | Low | Low | NA | Low | Low |
| | Brown 1969 | Unclear | | NA | Unclear | Low | NA | Unclear | Unclear |
| | Burr 1993; Merrett 1988; Miskelly 1988 | Low | | NA | Low | Low | NA | Low | Unclear |
| | de Jong 2002 | Low | | Na | High | Low | NA | Low | Low |
| | Du Toit 2016 | Unclear | | NA | Unclear | Low | NA | Low | Unclear |
| | Halmerbauer 2002, 2003 | Unclear | | NA | Unclear | Low | NA | Low | Unclear |
| | Hide 1994, 1996; Arshad 1992, 2003, 2007; Scott 2012 | Unclear | | NA | Low | Low | NA | Low | Unclear |
| | Johnstone 1966 | Unclear | | NA | High | Low | NA | Unclear | Unclear |
| | Kjellman 1979 | Unclear | | NA | Unclear | Low | NA | High | Unclear |
| | Lowe 2011 | Unclear | | NA | Low | Low | NA | Low | Unclear |
| | Matthew 1977 | Unclear | | NA | Unclear | High | NA | Unclear | High |
| | Natsume 2016 | Unclear | | NA | Low | Unclear | NA | Unclear | Unclear |
| | Palmer 2013 | Low | | NA | Low | Low | NA | Unclear | Low |
| | Perkin 2016 | Low | | NA | Unclear | Low | NA | Low | Unclear |
| | Shao 2006 | Unclear | | NA | Unclear | Low | NA | Unclear | Unclear |
| | Tan 2016 | Low | | NA | Low | Low | NA | Low | Low |
| | Zeiger 1989, 1992 and 1994 | Unclear | | NA | Low | High | NA | Unclear | High |
| | Zhou 2014 | Low | | NA | Low | Low | NA | High | Unclear |
| **Smith et al. 2016** | Perkin 2016 | Low | Unclear | Unclear | Low | Low | Low | Unclear | NA |
| | Dewey 1999 | High | High | Unclear | Unclear | Unclear | Unclear | Low | NA |
| | Cohen 1994 | High | High | Unclear | Unclear | Unclear | Unclear | High | NA |
| | Jonsdottir 2012 | Low | Low | Unclear | Low | Low | high | Low | NA |
| **Waidyatillake et al. 2018** | Tan 2016 | Low | Low | Low | Unclear | Low | Low | Low | NA |
| | Palmer 2016 | Low | Low | Low | Unclear | Low | Unclear | Unclear | NA |

[a]Except of the SR by EFSA Panel et al. 2019

| Primary review | Study ID | Risk of Bias (version 1) |
|---|---|---|
| | | Narrative assessment, not result-specific |
| **Burgess et al. 2019** | Bellach 2017 | Bellach et al.carried out a study in a sample from the general population but once more the planned sample size was not attained. With that limitation, the study did not find evidence that early introduction of egg reduced the risk of later egg allergy or sensitisation. |
| | Du Toit 2015 | This was a strong study with almost no loss to follow-up and excellent adherence to the assigned interventions. Obvious limitations were absence of a placebo control group and lack of generalizability given that the study population was high-risk. |
| | Halpern 1973 (not included in meta-analysis) | Randomization and allocation were not done correctly and the results must be regarded as unreliable. In addition, the presentation of the results did not allow inclusion in a meta-analysis. |
| | Palmer 2013, 2017 | Palmer et al. enrolled infants with eczema, limiting any findings in terms of generalizability to the wider infant population. Each of these studies was further limited by failure to attain the planned sample size. While the earlier study provided some evidence of a protective effect from early egg exposure, the later and larger study did not confirm these findings. |
| | Perkin 2016 | This was a strong study that enrolled exclusively breast-fed infants from an unselected general population in the UK, had little loss to follow-up, was adequately powered and food allergy was confirmed by double-blind, placebo-controlled food challenge. However, it should be noted that this was an open study and participation bias may have been introduced. Low per-protocol adherence in the intervention group was a limitation. |
| | Natsume 2017 | Natsume et al. enrolled a relatively small number of high-risk infants with eczema, limiting generalizability of the findings. |
| | Tan 2016 | The generalizability of this study was also limited as it was carried out in a high-risk population. |

| Primary review | Study ID | Strength of Recommendation Taxonomy (SORT) |
|---|---|---|
| | | Level of evidence (Not result-specific) |
| **Larson et al. 2017** | Palmer 2013 | 1 - High quality |
| | Du Toit 2015 | 1- High quality |

1- High quality: Studies that merit a "1" are systematic reviews of high quality studies with consistent findings, high quality randomized control trials (RCT), high quality prospective cohort studies with > 80% follow-up, and high quality diagnostic cohort studies. Level 1 studies also include key patient-oriented outcomes such as improvement in morbidity or mortality, symptoms, and quality of life or decreased costs (Larson et al. 2017).

### eTable 8. Summary of Findings of Primary Outcomes

| Interventions for preventing allergic outcomes in infants and young children | | | | | | | | |
|---|---|---|---|---|---|---|---|---|
| **Systematic Review** | **Intervention and Comparator** | **Age exposure** | **Outcome details** | **Age outcome** | **Effect estimates and 95% Confidence intervals** | **Number of studies (participants)** | **Quality of the evidence (GRADE)[c]** | **Comments** |
| **Atopic Diseases (Outcome-Cluster)** | | | | | | | | |
| **EFSA Panel 2019** | Early introduction of CF in general | 3-4 m ***vs.*** 6 m | Odds of developing an atopic disease | Up to 3 y | OR 1.10 (0.86-1.40) | 1 RCT [13] (1139) | ⊕◯◯◯ Very low[a] | Study population: general population |
| | Introduction of specific foods (egg yolk consumption) | <0.7 m ***vs.*** >6 m | Odds of developing an atopic disease | 5-7 y | OR 2.27 (0.70-7.40) | 1 RCT [14] (978) | Not available | Study population: general and high-risk population<br>Outcome was diagnosed by a physician;<br>The authors deemed the evidence basis for this intervention to sparse to draw conclusions and therefore did not assess the quality of evidence and did not consider this study further. |
| **Food Allergy in General** | | | | | | | | |
| **Burgess et al. 2019** | Early introduction of a group of six allergenic foods ***vs.*** standard introduction | >3 m ***vs***. >6 m | Risk of developing food allergy | 12-36m | RR 0.80 (0.51-1.25) | 1 RCT [13] (1303) | ⊕◯◯◯ Very low[b] | Study population: general population<br>ITT analysis was used, different results in a PP analysis |
| **De Silva et al. 2020** | Early introduction of CF (*6 allergenic foods*) ***vs.*** exclusive breastfeeding to approximately 6 m | from 3 m ***vs.*** later introduction | Risk of developing food allergy (cumulative prevalence) | 12-36 m | RR 0.8 (0.51-1.25) | 1 RCT [13] (1303) | ⊕◯◯◯ Very low[a] | Study population: general population<br>Overall conclusion: Probably no reduction of food allergy |
| **EFSA Panel 2019** | Introduction of CF in general | 3-4 m ***vs.*** 6 m | Risk of developing symptomatic food allergy | 12-36 m | RR 0.80 (0.51-1.25)<br>PP Analysis:<br>RR 0.33 (0.13-0.83) | 1 RCT [13] (1162) | ⊕◯◯◯ Very low[a] | Study population: general population |

| Interventions for preventing allergic outcomes in infants and young children | | | | | | | | |
|---|---|---|---|---|---|---|---|---|
| **Systematic Review** | **Intervention and Comparator** | **Age exposure** | **Outcome details** | **Age outcome** | **Effect estimates and 95% Confidence intervals** | **Number of studies (participants)** | **Quality of the evidence (GRADE)[c]** | **Comments** |
| **Larson et al. 2017** | Delayed introduction of potentially allergenic foods until after 12 m of age ***vs.*** introduction of allergenic food prior to 12 m | 4-11 m ***vs.*** > 12 m | Risk of developing food allergy | 60 m | **Low risk infants:** Absolute RD 11.8 % (3.4-20.3), p<0.001 (86.1 % relative reduction in the prevalence of peanut allergy) **High risk infants:** Absolute RD 24.7 % (4.9-43.3),p = 0.004 (70 % relative reduction in the prevalence of peanut allergy) | 1 RCT [15] (640) | ⊕◯◯◯ Very low[b] | Study population: high-risk population |
| **Scarpone et al. 2023** | Earlier ***vs.*** later introduction of multiple allergenic foods | 2-12 m ***vs.*** later | Risk of developing allergy to any food | 1-3 y | RR 0.49 (0.33-0.74) $I^2$=49 %, $\tau^2$=0.08 Absolute risk difference for a population with 5 % incidence of food allergy was −26 cases per 1000 population (95% CI −34 to −13 cases per 1000 population) Sensitivity analysis for low RoB data: RR 0.39 (0.24-0.65), $I^2$=0% | 4 RCTs [13,16-18] (3295) Studies with low RoB: 2 RCTs [16,18] (2000) | ⊕⊕⊕◯ Moderate | Study population: general and high-risk population Scarpone et al.[19] reported that the statistical heterogeneity was explained by a less pronounced effect in one large study (p. E3); They indicated that the trial sequential analysis showed that the heterogeneity-adjusted optimal information size for detection of a 30 % risk reduction had not been reached (p. E3, eFigure 2 in Supplement 1) |
| | Earlier ***vs.*** later introduction of egg | 2-12 m ***vs.*** 6-24 m | Risk of developing allergy to any food | 18-48 m | RR 0.50 (0.34-0.75) $I^2$=45 %, $\tau^2$=0.07 Sensitivity analysis for low RoB data: RR 0.39 (0.24-0.65) $I^2$=0 % | 4 RCTs [13,16-18] (3295) Studies with low RoB: 2 RCTs [16,18] (2000) | ⊕◯◯◯ Very low | Study population: general and high-risk population |

| Interventions for preventing allergic outcomes in infants and young children | | | | | | | | |
|---|---|---|---|---|---|---|---|---|
| **Systematic Review** | **Intervention and Comparator** | **Age exposure** | **Outcome details** | **Age outcome** | **Effect estimates and 95% Confidence intervals** | **Number of studies (participants)** | **Quality of the evidence (GRADE)[c]** | **Comments** |
| **Scarpone et al. 2023** | Earlier ***vs.*** later introduction of peanut | 3-10 m ***vs***. ≥ 6-60 m | Risk of developing allergy to any food | 18-60 m | RR 0.60 (0.38-0.94)<br>Sensitivity analysis for low RoB data:<br>RR 0.39 (0.24-0.65)<br>$I^2$=0 % | 5 RCTs [13,15-18] (3927)<br>Studies with low RoB: 2 RCTs [16,18] (2000) | ⊕◯◯◯<br>Very low | Study population: high-risk and general population |
| | Earlier ***vs.*** later introduction of cow's milk | 0-5 m ***vs.*** 5-12 m | Risk of developing allergy to any food | 18-48 m | RR 0.67 (0.39-1.13)<br>Sensitivity analysis for low RoB data:<br>RR 0.39 (0.24-0.65)<br>$I^2$=0 % | 6 RCTs [13,16-18,20,21] (3981)<br>Studies with low RoB: 2 RCTs [16,18] (2000) | ⊕◯◯◯<br>Very low | Study population: high-risk and general population |
| | Early introduction of wheat | 2-12 m ***vs.*** 6-12m | Risk of developing allergy to any food | 11-60 m | RR 0.40 (0.20-0.79)<br>$I^2$=72 % | 4 RCTs [13,16-18] (3250) | ⊕⊕◯◯<br>Low[b] | Study population: high-risk and general population |
| | Early introduction of soya | 0-12 m ***vs.*** 6-12m | Risk of developing allergy to any food | 6-36 m | RR 0.74 (0.19-2.93)<br>$I^2$= 88 % | 2 RCTs [16,17] (545) | | Study population: high-risk and general population |
| | Early introduction of fish | 2-12 m ***vs.*** 6-24 m | Risk of developing allergy to any food | 12-48 m | RR 0.31 (0.04-2.44)<br>$I^2$=88 % | 2 RCTs [13,17] (1250) | ⊕◯◯◯<br>Very low[b] | Study population: high-risk and general population |
| | Earlier introduction of crustaceans or tree nuts | 2-12 m ***vs.*** 28 d-12 m | Risk of developing allergy to any food | After 24- 48 m | Allergy to any food developed in 2/44 ***vs.*** 21/44 participants in the earlier vs. later introduction group[19] (eTable 12) | 1 RCT [16] (88) | ⊕⊕◯◯<br>Low[b] | Study population: high-risk population<br>Only one study of introduction of crustaceans or tree nuts (with other foods), reporting this outcome, was identified (88 participants). |

| Interventions for preventing allergic outcomes in infants and young children | | | | | | | | |
|---|---|---|---|---|---|---|---|---|
| **Systematic Review** | **Intervention and Comparator** | **Age exposure** | **Outcome details** | **Age outcome** | **Effect estimates and 95% Confidence intervals** | **Number of studies (participants)** | **Quality of the evidence (GRADE)[c]** | **Comments** |
| **Smith et al. 2016** | Early introduction of potentially allergenic food ***vs.*** exclusively breast-feeding | 3 m ***vs***. >6 m | Risk of developing food allergy to one or more foods | 12-36 m | RR 0.80 (0.51-1.25)<br>Risk with exclusive breast-feeding infants: 71 per 1000<br>Risk with non-exclusive breastfeeding infants (foods): 56 per 1000 | 1 RCT [13] (1162) | ⊕◯◯◯ Very low[a] | Study population: general population<br>Overall conclusion: Early introduction of potentially allergenic foods was not found to reduce the risk of "food allergy" to one or more of these foods between one to three years of age compared to the group exclusively breastfeeding to about six months. |
| Risk of egg allergy | | | | | | | | |
| **Al-Saud et al. 2018** | Early introduction of egg powder ***vs.*** no early egg introduction | 3-9 m ***vs.*** exclusive breastfeeding until 6 m or placebo | Risk of developing egg allergy | Median 12 m | RR 0.60 (0.44-0.82)<br>$I^2$=23 %, p=0.002<br>Absolute risk reduction for a population with an incidence of egg allergy of 9.3 % was 37 fewer cases (95 % CI 17-52 cases) per 1,000 people | 6 RCTs[13,22-26] (2663) | ⊕⊕⊕⊕ High[a] | Study population: high-risk and general population<br>Heterogeneity due to dose of egg protein introduced to infants;<br>Subgroup analysis which excludes the studies using heated eggs[13,23]:<br>Early introduction of egg using a lower dose of egg protein (< 4,000 mg/week) may be associated with a more successful prevention of egg allergy (RR 0.65, 95% CI 0.43–0.99, p=0.04, $I^2$=0 %, ***vs.*** RR 0.69, 95% CI 0.41–1.16, p=0.16, $I^2$=0 %) |
| **Burgess et al. 2019** | Early exposure to egg or food containing egg over similar age ranges ***vs.*** varying later rages | 4-6 m ***vs.*** varying later ages | Odds of developing egg allergy | 12 m | OR 0.63 (0.44-0.90)<br>$I^2$=40.6 %, p=0.134 | 6 RCTs [13,22,23,25-27] (3058) | ⊕⊕⊕◯ Moderate[b] | Study population: high-risk and general population |
| **Dai et al. 2019** | Introduction of egg | <6 m ***vs.*** >6 m | Risk of developing egg allergy | 12-36 m | RR 0.60 (0.46-0.79)<br>$I^2$=33 %, p< 0.05 | 6 RCTs [13,22-26] (2663) | ⊕⊕⊕◯ High[b] | Study population: high-risk and general population<br>Subgroup analyses for:<br>Family history:<br>RR 0.55 (95%CI 0.40-0.75) (p= 0.18) |

| Interventions for preventing allergic outcomes in infants and young children | | | | | | | | |
|---|---|---|---|---|---|---|---|---|
| **Systematic Review** | **Intervention and Comparator** | **Age exposure** | **Outcome details** | **Age outcome** | **Effect estimates and 95% Confidence intervals** | **Number of studies (participants)** | **Quality of the evidence (GRADE)[c]** | **Comments** |
| **Dai et al. 2019** | | | | | | | | Raw eggs: RR 0.67 (95% CI 0.49-0.93) (p=0.56)<br>Doses (0-4g): RR 0.55 (95%CI 0.36-0.85) (p=0.16)<br>Time (4-6m ***vs.*** <4m): RR 0.58 (95%CI 0.43-0.78) (p=0.12) |
| **De Silva et al. 2020** | Introducing CF (*cooked egg*) | >6 m ***vs.*** placebo | Risk of developing egg allergy | 12 m | RR 0.22 (0.09-0.54) | 1 RCT [23] (147) | ⊕⊕⊕◯ Moderate[a] | Study population: high-risk population |
| | Introducing CF (*raw egg/ pasteur-ized egg powder*) | >4 m ***vs.*** usual diet with placebo | Risk of developing egg allergy | 12 m | Range of RRs: 0.65 (0.38-1.11) - 3.3 (0.35- 31.32) | 3 RCTs [22,24,25] (1289) | ⊕⊕◯◯ Low[a] | Study population: high-risk and general population |
| **EFSA Panel 2019** | Introduction of egg | General population: 3-4 m/4-6 m ***vs.*** 6m/ egg avoidance<br><br>At-risk population: 4-6 m ***vs.*** 8-10 m | Risk of developing symp-tomatic egg allergy | General population: 12 or 36 m<br><br>At-risk population: 12 m | General population: RR 0.69 (0.40-1.18)[13] (FA) RR 3.3 (0.35-31.32)[22] (egg allergy)<br><br>At-risk population: RR 0.69 (0.51- 0.93); $I^2$ =0 % (egg allergy) | 5 RCTs [13,22,25-27] General population: 2 RCTs [13,22]: (1568)[c]<br><br>At-risk populations: 3 RCTs (1087)[c] | ⊕⊕◯◯ Low[b] | EFSA Panel et al.[7] concluded from four RCTs (Tiers 1 and 2) that introduction of egg at 3-4 months of age compared with 6 months of age may reduce the risk of developing egg allergy (p. 83);<br><br>The RCT by Bellach et al.[22] was not considered in the conclusions as it was underpowered and it was not comparable to the other available evidence and the inconsistent findings might be explained by these factors according to EFSA Panel et al. (p.81). |
| **Ierodiakonou et al. 2016** | Introduction of egg | 4-6 m ***vs.*** ≥6 m or placebo | Risk of developing egg allergy | Up to 36 m (median 12 m) | RR 0.56 (0.36–0.87), p= 0.009<br>$I^2$=35.8 %, p=0.1829<br>Absolute risk reduction for a population with 5.4 % incidence of egg allergy was 24 cases | 6 RCTs (3665) of which 5 RCTs [13,23,26-28] (1915) were in-cluded in the meta-analysis<br><br>Sensitivity analyses: | ⊕◯◯◯ Very low[a] | Study population: high-risk and general population<br>According to Ierdiakonou et al.[8] the heterogeneity was due to the abstract publication by Natsume et al.[23] — the authors declined to share further infor-mation about their study (p.1183).<br>Sensitivity analysis (1): excluding studies at high or unclear risk of bias (by using RoB tool 1.0) |

| Interventions for preventing allergic outcomes in infants and young children | | | | | | | | |
|---|---|---|---|---|---|---|---|---|
| **Systematic Review** | **Intervention and Comparator** | **Age exposure** | **Outcome details** | **Age outcome** | **Effect estimates and 95% Confidence intervals** | **Number of studies (participants)** | **Quality of the evidence (GRADE)[c]** | **Comments** |
| **Ierodiakonou et al. 2016** | | | | | (95% CI 7-35 cases) per 1000 population<br>Sensitivity analysis (1): RR 0.65 (0.46-0.92) $I^2$ =0 %, p=0.9527<br>Sensitivity analysis (2): RR 0.63 (0.40-0.99) $I^2$=0%, p=0-842 | (1) 3 RCTs [13,26,27] (1496)<br>(2) 2 RCTs [26,27] (331) | | Sensitivity analysis (2): excluding studies at high risk of bias (by using RoB tool 1.0) |
| **Larson et al. 2017** | Delayed introduction of potentially allergenic foods until after 12 m of age ***vs.*** introduction of allergenic food prior to 12 m | <4 m ***vs.*** >8 m | Risk of developing egg allergy | 12 m | No significant differences in presence of egg allergy between infants introduced to egg at <4 m and those introduced at >8 m | 1 RCT [27] (86) | ⊕⊕◯◯ Low[b] | Study population: high-risk population<br>Overall conclusion: There was no evidence that delaying allergenic foods beyond 4 to 8 months of age is beneficial in reducing allergies in infants. |
| **Scarpone et al. 2023** | Earlier ***vs.*** later introduction of egg | 3-9 m ***vs.*** ≥ 6 m-15 m | Risk of developing egg allergy | 6-36 m | RR 0.60 (0.46-0.77) $I^2$=0 %; $\tau^2$=0.00<br>Absolute risk difference for a population with 4 % incidence of egg allergy was −16 cases per 1000 population (95% CI -22 to -9 cases per 1000 population)<br>Sensitivity analysis for low RoB data: RR 0.39 (0.24-0.65) $I^2$=0 % | 9 RCTs [13,16,18,22,23,25-27,29] (4811)<br>Studies with low RoB: 2 RCTs [16,18] (2000) | ⊕⊕⊕⊕ High | Study population: high-risk and general population<br>Scarpone et al.[19] reported that the trial sequential analysis has shown that the heterogeneity-adjusted optimal information size for detection of a 30 % risk reduction had been reached (p. E4);<br>They indicated that the subgroup analysis has found a significant interaction related to dose of egg, with evidence for a greater reduction in risk of egg allergy in the low-dose group (p=0.02 for interaction) (p, E4, eTable 8 in Supplement 1). |

| Interventions for preventing allergic outcomes in infants and young children | | | | | | | | |
|---|---|---|---|---|---|---|---|---|
| **Systematic Review** | **Intervention and Comparator** | **Age exposure** | **Outcome details** | **Age outcome** | **Effect estimates and 95% Confidence intervals** | **Number of studies (participants)** | **Quality of the evidence (GRADE)[c]** | **Comments** |
| **Risk of peanut allergy** | | | | | | | | |
| **Burgess et al.** **2019** | Early exposure to peanut or allergenic food containing peanut ***vs.*** late introduction/ avoidance | >3 m ***vs.*** >6 m or 4-11 m ***vs.*** avoidance | Odds of developing peanut allergy | 12-36 m or 60 m | OR 0.28 (0.14-0.57) $I^2$= 43.5 %, p=0.170 | 2 RCTs [13,15] (1943) | ⊕⊕◯◯ Low[b] | Study population: high-risk and general population |
| **Dai et al.** **2019** | Introduction of peanut | 3/4-11 m ***vs.*** >6 m/ peanut avoidance up to 5y | Risk of developing peanut allergy | Up to 5 y | Not available | 2 RCTs [13,15] (1793) | Not applicable | Study population: high-risk and general population<br><br>No meta-analysis conducted;<br>The authors did not draw conclusions from the evidence of both studies because of conflicting evidence that is assumed to be due to heterogeneity of the study populations and the interventions. |
| **De Silva et al. 2020** | Early introduction of CF (*peanut*) ***vs.*** completely abstaining from peanut in the first 5 y | from 4-11 m (median 7.8 m) ***vs.*** peanut avoidance | Peanut allergy incidence | 60 m | Range of IRRs: 0.14 (0.05-0.34) - 0.35 (0.14-0.85) | 2 RCTs [15,30] (640) | ⊕◯◯◯ Very low[a] | Study population: high-risk population |
| **EFSA Panel** **2019** | Introduction of peanut | 3-4 m ***vs.*** 6 m | Risk of developing symtomatic peanut allergy | Up to 36 m | RR 0.49 (0.20-1.19)<br><br>PP Analysis:<br>0 % ***vs.*** 2.5 %, p=0.003; RR not calculable due to zero events in the early introduction group | 1 RCT [13] (1168) | ⊕◯◯◯ Very low[a] | Study population: general population |

| Interventions for preventing allergic outcomes in infants and young children | | | | | | | | |
|---|---|---|---|---|---|---|---|---|
| **Systematic Review** | **Intervention and Comparator** | **Age exposure** | **Outcome details** | **Age outcome** | **Effect estimates and 95% Confidence intervals** | **Number of studies (participants)** | **Quality of the evidence (GRADE)[c]** | **Comments** |
| **Ierodiakonou et al. 2016** | Introduction of peanut | From 3/ 4-11m ***vs.*** peanut avoidance | Risk of developing peanut allergy | Up to 72 m | RR 0.29 (0.11–0.74)<br>$I^2$ =66.1 %,(p=0.0857<br>Absolute risk reduction for a population with 2.5 % incidence of peanut allergy was 18 cases (95% CI 6-22 cases) per 1000 population. | 2 RCTs [13,15] (1793) | ⊕◯◯◯<br>Very low[a] | Study population: high-risk and general population<br>Ierodiakonou et al.[8] reported that the high heterogeneity was attributed to the high treatment adherence in the study by Du Toit et al.[15] compared with more variable treatment adherence in the study by Perkin et al.[13] (p. 1183). |
| **Scarpone et al. 2023** | Earlier ***vs.*** later introduction of peanut | 3-10 m ***vs***. ≥ 6-60 m | Risk of developing peanut allergy | 6-60 m | RR 0.31 (0.19-0.51)<br>Sensitivity analysis for low RoB data:<br>RR 0.40 (0.19-0.82)<br>$I^2$=0 % | 4 RCTs [13,15,16,18] (3796)<br>Studies with low RoB: 2 RCTs [16,18] (2000) | ⊕⊕⊕⊕<br>High | Study population: high-risk and general population<br>Low heterogeneity:<br>$\tau^2$=0.06; $\chi^2$=3.80; df=3 (p=0.28); $I^2$=21 % |
| **Risk of grain allergy** | | | | | | | | |
| **Chmieleweska et al. 2017** | Early introduction of six allergenic foods ***vs.*** standard introduction | 3 m v***s.*** >6 m | Risk of developing wheat allergy | 36 m | 1/572 ***vs.*** 0/597, p=0.49 | 1 RCT [13] (1303) | ⊕⊕◯◯<br>Low[b] | Study population: general population<br>According to Chmielweska et al.[4] there was only one case of wheat allergy (early introduction group), so drawing firm conclusions was not possible (p.893);<br>The authors reported that delaying the introduction of solid foods after 6 months of age was only one of the components of the multifaceted intervention in this study (p. 894). |
| **EFSA Panel 2019** | Introduction of cereals (*wheat*) | 3-4 m ***vs.*** 6 m | Risk of developing wheat allergy | 12 m or 36 m | Not available | 1 RCT [13] (1169) | ⊕⊕◯◯<br>Low[a] | Study population: general population |
| **Scarpone et al. 2023** | Early introduction of wheat | 3-4 m ***vs.*** 6-7 m | Risk of developing wheat allergy | 6-60 m | RR 0.66 (0.10-4.47)<br>$I^2$=2 % | 3 RCTs [13,16,18] (3169) | ⊕◯◯◯<br>Very low[b] | Study population: high-risk and general population |

| Interventions for preventing allergic outcomes in infants and young children | | | | | | | | |
|---|---|---|---|---|---|---|---|---|
| **Systematic Review** | **Intervention and Comparator** | **Age exposure** | **Outcome details** | **Age outcome** | **Effect estimates and 95% Confidence intervals** | **Number of studies (participants)** | **Quality of the evidence (GRADE)[c]** | **Comments** |
| **Risk of Cow's milk allergy** | | | | | | | | |
| **Dai et al. 2019** | Introduction of cow's milk protein | 3-6 m ***vs.*** ≥ 6 m | Risk of developing milk protein allergy | 12-36 m | No meta-analysis conducted | 2 RCTs [13,20] (1550) | Not applicable | Study population: high-risk and general population<br>Both studies demonstrated no significant difference between the intervention groups;<br>The studies differed in terms of interventions and study characteristics |
| **Ierodiakonou et al. 2016** | Introduction of cow milk | From birth (median 4 m) ***vs.*** placebo or 3 m ***vs.*** ≥6 m | Risk of developing milk allergy | Up to 84 m | RR 0.76 (0.32-1.78)<br>$I^2$ =0 %, p=0.9498 | 2 RCTs [13,20] (1550) | ⊕◯◯◯<br>Very low[a] | Study population: high-risk and general population |
| **Scarpone et al. 2023** | Earlier ***vs.*** later introduction of cow's milk | 0-5 m ***vs.*** 5-12 m | Risk of developing cow's milk allergy | 6-48 m | RR 0.84 (0.38-1.87)<br>Sensitivity analysis for low RoB data:<br>RR 0.32 (0.09-1.18)<br>$I^2$=0 % | 6 RCTs [13,16,18,20,21,31] (3900)<br>Studies with low RoB: 2 RCTs [16,18] (2000) | ⊕◯◯◯<br>Very low | Study population: high-risk and general population<br>Heterogeneity:<br>$\tau^2$=0.35; $\chi^2$=7.81; df= 5 (p=0.17); $I^2$=36% |
| **Other allergenic foods** | | | | | | | | |
| **Scarpone et al. 2023** | Early introduction of soya | 3-4 m **vs.** 12 weeks later | Risk of developing allergy to soya | 6-18 m | Soya allergy developed in 1/82 ***vs.*** 0/79 participants in the earlier vs later introduction group[19] (eTable 12) | 1 RCT [16] (161) | ⊕◯◯◯<br>Very low[b] | Study population: high-risk population |
| | Early introduction of fish | 3-6 m ***vs.*** 6 m | Risk of developing allergy to fish | 12-48 m | Fish allergy developed in 1/573 ***vs.*** 1/601 participants in the earlier vs later introduction group[19] (eTable 12) | 1 RCT [13] (1174) | ⊕⊕◯◯<br>Low[b] | Study population: general population |

| Interventions for preventing allergic outcomes in infants and young children | | | | | | | | |
|---|---|---|---|---|---|---|---|---|
| **Systematic Review** | **Intervention and Comparator** | **Age exposure** | **Outcome details** | **Age outcome** | **Effect estimates and 95% Confidence intervals** | **Number of studies (participants)** | **Quality of the evidence (GRADE)[c]** | **Comments** |
| **Eczema** | | | | | | | | |
| **Al-Saud et al. 2018** | Early introduction of egg powder ***vs.*** no early egg introduction | 3-9 m ***vs.*** exclusive breastfeeding until 6m/ placebo | Risk of developing eczema | Median 12 m | RR 0.90 (0.70-1.16)<br>$I^2$=0 %, p=0.42<br>19 cases fewer per 1,000 | 2 RCTs [25,26] (1001) | ⊕⊕⊕◯<br>Moderate[a] | Study population: high-risk population<br><br>No significant difference in the development of eczema by the timing of egg introduction. |
| **EFSA Panel 2019** | Early **vs.** late Introduction of egg | 4-6.5 m ***vs.*** >8 - >10m | Odds of developing eczema (endpoints 'symptomatic eczema' and 'atopic dermatitis' as investigated in the individual studies) | 12 m | OR 0.82 (0.01-130.12)<br>$I^2$=69 %, p=0.07 | 2 RCTs [25,26] (1001) | ⊕◯◯◯<br>Very low[a] | Study population: high-risk population |
| **Ierodiakonou et al. 2016** | Early introduction of cow's milk | Multifaceted interventions:<br>(a) Standard care ***vs.*** dietary avoidance or delayed food introduction;<br>(b) Early introduction from birth to 6 m/ until 9 m **vs.** placebo/ avoidance or delayed introduction after 6 m | Risk of developing eczema | ≤4 y and 5-14 years | **≤4 years**<br>RR 1.14 (0.87-1.49)<br>$I^2$=54.1 %, p=0.0328<br><br>**5-14 years**<br>RR 1.05 (0.90-1.23)<br>$I^2$=0 %, p=0.4743 | **≤4 years**<br>8 RCTs [20,31-37] (2333)<br>**5-14 years**<br>6 RCTs [20,38-42] (1665) | **≤4 years**<br>⊕◯◯◯<br>Very low[b]<br><br>**5-14 years**<br>⊕◯◯◯<br>Very low[b] | Study population: high-risk and general population<br>Subgroup analyses for:<br>**Risk of eczema/atopic dermatitis at age ≤4 years:**<br>High risk of disease (6 RCTs):<br>RR 1.07 (95 % CI 0.80-1.44), $I^2$=60.2 % (p=0.21)<br>Low/Normal risk of disease (2 RCTs):<br>RR 1.59 (95% CI 0.92-2.76), $I^2$=0 % (p=0.12)<br>Intervention multifaceted (3 RCTs):<br>RR 1.96 (95 % CI 0.89-4.30), $I^2$=60.9 % (p=0.1210)<br>Intervention not multifaceted (5 RCTs):<br>RR 0.98 (95 % CI 0.77-1.24), $I^2$=36.1 % (p=0.1210)<br>Overall risk of bias high/unclear (7 RCTs):<br>RR 1.13 (95% CI 0.85-1.50), $I^2$=59.4 % (p=0.63)<br>Overall risk of bias low (1 RCT):<br>RR 1.49 (95% CI 0.50-4.48), $I^2$=n.a. (p=0.63)<br><br>**Risk of eczema/atopic dermatitis at age 5-14 years:** |

| Interventions for preventing allergic outcomes in infants and young children | | | | | | | | |
|---|---|---|---|---|---|---|---|---|
| **Systematic Review** | **Intervention and Comparator** | **Age exposure** | **Outcome details** | **Age outcome** | **Effect estimates and 95% Confidence intervals** | **Number of studies (participants)** | **Quality of the evidence (GRADE)[c]** | **Comments** |
| | | | | | | | | High risk of disease (6 RCTs):<br>RR 1.05 (95 % CI 0.90-1.23), $I^2$=0 %<br>Intervention multifaceted (3 RCTs):<br>RR 1.22 (95 % CI 0.94-1.59), $I^2$=0 % (p=0.18)<br>Intervention not multifaceted (3 RCTs):<br>RR 0.97 (95 % CI 0.80-1.19), $I^2$=2.8 % (p=0.18)<br>Overall risk of bias high/unclear (6 RCTs):<br>RR 1.05 (95% CI 0.90-1.23), $I^2$=0 % |
| **Smith et al. 2016** | Early introduction of potentially allergenic food ***vs.*** exclusively breastfeeding | 3 m | Risk of developing visible eczema at 12 m visit (stratified by visible eczema at enrolment) | 12 m | RR 0.86 (0.51-1.44)<br>Risk with exclusive breastfeeding infants: 182 per 1000<br>Risk with non-exclusive breastfeeding infants (foods): 156 per 1000 (93 to 262) | 1 RCT [13] (284) | ⊕⊕⊕◯<br>Moderate[a] | Study population: general population<br><br>Overall conclusion:<br>The early introduction of potentially allergenic foods did not reduce the risk of visible eczema at 12 months stratified by visible eczema at enrolment (at three months of age). |
| **Waidyatillake et al. 2019** | Introduction of egg | 4-10 m | Odds of developing eczema | 12 m | OR 0.87 (0.68-1.12)<br>$I^2$=0.0 %, p=0.788 | 2 RCTs [13,43] (1139) | ⊕⊕◯◯<br>Low[a] | Study population: high-risk and general population<br><br>When the children with eczema were stratified by atopic status, there was some evidence for a reduction of atopic eczema in the intervention group (p=0.09) (see Waidyatillake et al. 2019, p. 1003). |
| **Asthma** | | | | | | | | |
| **EFSA Panel 2019** | Timing of introduction of CF in general | 3-4 m ***vs.*** 6 m | Odds of developing asthma-like symptoms (endpoints 'wheeze', 'asthma' and associated endpoints | Up to 36 m | OR 0.98 (0.47-2.05) | 1 RCT [13] (1127) | ⊕⊕◯◯<br>Low[a] | Study population: general population |

| Interventions for preventing allergic outcomes in infants and young children | | | | | | | | |
|---|---|---|---|---|---|---|---|---|
| **Systematic Review** | **Intervention and Comparator** | **Age exposure** | **Outcome details** | **Age outcome** | **Effect estimates and 95% Confidence intervals** | **Number of studies (participants)** | **Quality of the evidence (GRADE)[c]** | **Comments** |
| | | | as investigated in the individual studies) | | | | | |
| | Timing of introduction of egg | 4-6.5 m ***vs***. ≥ 10 m | Odds of developing asthma-like symptoms (endpoints 'wheeze', 'asthma' and associated endpoints as investigated in the individual studies) | 12 m | RR 1.12 (0.87-1.45) | 1 RCT [25] (820) | ⊕⊕◯◯ Low[a] | Study population: high-risk population |
| **Allergic Rhinitis** | | | | | | | | |
| **EFSA Panel 2019** | Introduction of CF in general | 3-4 m ***vs.*** 6 m | Odds of developing allergic rhinitis | Up to 36 m | OR 1.01 (0.57-1.79) | 1 RCT [13] (1127) | 3/ 5 domains not applicable | Study population: general population |
| **Ierodiakonou et al. 2016** | Early introduction of cow's milk | Multifaceted interventions: (a) Standard care ***vs.*** dietary avoidance or delayed food introduction; (b) Early introduction from birth to 6 m/ until 9 m ***vs.*** placebo/ avoidance or delayed introduction after 6 m for further information | Risk of developing allergic rhinitis | ≤4 years or 5-14 years | **≤4 years** RR 1.52 (0.97-2.37) $I^2$=69.9 %, p=0.0053 Sensitivity analysis: RR 1.31 (0.95-1.82) $I^2$=50 % **5-14 years** RR 1.21 (0.85-1.73) $I^2$=78.7 %, p=0.0003 Sensitivity analysis: RR 0.98 (0.82-1.18) $I^2$=21 % | **≤4 years** 6 RCTs [31,37,42,44-46] (1735) **5-14 years** 5 RCTs [20,39,40,42,47] (1673) | **≤4 years** ⊕⊕◯◯ Low[b] **5-14 years** ⊕⊕◯◯ Low[b] | Study population: high-risk population **≤4 years and 5-14 years:** The statistical heterogeneity was partly explained by the positive findings in the study of Johnstone et al. 1966 (see Ierodiakonou et al. 2016, archived Supplement AR, p. 12); The sensitivity analyses (Johnstone et al. 1966 was excluded) found reduced statistical heterogeneity, and no significant effect (see Ierodiakonou et al. 2016, archived Supplement, p. 12); Subgroup analysis of early cow's milk introduction and risk of atopic dermatitis at ages ≤ 4 and 5-14 years did not show any significant subgroup differences, although opportunities for meaningful subgroup analysis were limited; |

| Interventions for preventing allergic outcomes in infants and young children | | | | | | | | |
|---|---|---|---|---|---|---|---|---|
| Systematic Review | Intervention and Comparator | Age exposure | Outcome details | Age outcome | Effect estimates and 95% Confidence intervals | Number of studies (participants) | Quality of the evidence (GRADE)[c] | Comments |
| | | see Ierodiakonou et al. (2016) | | | | | | |

[a] GRADE assessment for single domains was extracted except of dimension 1 (ROB), which was re-assessed by the authors based on the self-assessed ROB 2.0 tool results. The integration of the ratings of the single domains was also redone.
[b] All GRADE dimensions were re-assessed by the overview authors because there was no GRADE assessment available in the original systematic review.
[c] Number of participants (at baseline) was extracted from the primary studies by the overview authors due to missing reported data in the systematic review.
FA=Food allergy, RCT=Randomised controlled trial, SR=Systematic review, RR=Risk ratio, OR=Odds ratio, RoB=Risk of bias

### eTable 9. Certainty of Evidence Assessments (GRADE approach) for all Primary Outcomes

| | Certainty assessment | | | | | | | Comments |
|---|---|---|---|---|---|---|---|---|
| Systematic Review | № of studies | Study design | Risk of bias | Inconsistency | Indirectness | Imprecision | Other considerations | |
| Food allergy in general | | | | | | | | |
| **Burgess et al. 2019** | **1** | RCT | Serious; the RoB dimension “Measurement of the outcome” resulted in "high" | Not applicable (only one underlying primary study) | Serious; study populations consisted of exclusively breastfed infants in Australia only, which is a country with an unexplained higher allergy prevalence as others; in the intervention multiple foods are introduced, which may not be generalisable to the general population | Serious; the effect estimate comes from only one trial; the 95% CI is wide and the number of events is low | None | The certainty of evidence was downgraded three levels due to serious concerns in RoB, indirectness and imprecison;<br>the evidence is based on a single study, assessing the domain „inconsistency“ is therefore not applicable (4/5 domains considered) |
| **EFSA Panel 2019** | **1** | RCT | Serious; the RoB dimension “Measurement of the outcome” resulted in "high" | Not applicable (only one underlying primary study) | Serious; the study population consisted of breastfed infants only; the authors of the primary SR considered that the study results cannot be generalised to formula fed infants and thus to the whole population of infants living in Europe (EFSA Panel et al. 2019, p. 79ff) | Serious; the effect estimate comes from one trial only; wide CI including no effect | None | Intervention: Introduction of CF in general; the certainty of evidence was downgraded three levels due to serious concerns in RoB, and serious concerns regarding indirectness and imprecision;<br>the evidence is based on a single study, assessing the domain „inconsistency“ is therefore not applicable (4/5 domains considered) |

| | Certainty assessment | | | | | | | Comments |
|---|---|---|---|---|---|---|---|---|
| **Systematic Review** | **№ of studies** | **Study design** | **Risk of bias** | **Inconsistency** | **Indirectness** | **Imprecision** | **Other considerations** | |
| **EFSA Panel 2019** | **5** | RCT | Not serious | Not serious; $I^2$ =0 % | Serious; three of four trials were conducted in at-risk study populations (Palmer et al. 2013, Palmer et al. 2017, Tan et al. 2017) in Australia, a country that has an unexplained higher prevalence of allergy than Europe; the trials used pasteurised raw egg powder as an intervention product, which is not the form that would be used when egg is introduced to infants according to EFSA Panel et al. 2019 (p. 67) | Serious; one trial reported no effect with a wide confidence interval including potential for benefits and harms, while evidence of three trials suggests a small risk reduction in the intervention group | None | Intervention: Introduction of egg; The certainty of evidence was downgraded three levels due to serious concerns in RoB, and serious concerns regarding indirecntess and imprecision |
| **De Silva et al. 2020** | **1** | RCT | Serious; the RoB dimension “Measurement of the outcome” resulted in "high" | Serious; inconsistency in results based on per protocol or intention to treat analysis in reference to table S3 of the primary SR | Very serious; multiple foods introduced, therefore it is more difficult to assign causality; conducted in a country with high prevalence of allergy; issues with adherence (De Silva et al 2020, S3 Table) | Serious; issues related to loss to follow up and associated impact on imprecision (De Silva et al 2020, S3 Table) | None | The certainty of evidence was downgraded three levels due to serious concerns in RoB, inconsistency and imprecision and very serious concerns regarding indirectness |
| **Smith et al. 2016** | **1** | RCT | Serious; the RoB dimension “Measurement of the outcome” resulted in "high" | Not applicable (only one underlying primary study) | Not serious | Serious; wide CI crossing the line of no effect and small sample size (p. 8) | None | The certainty of evidence was downgraded three levels due to very serious concerns regarding RoB, and serious concerns regarding imprecision |

| | Certainty assessment | | | | | | | Comments |
|---|---|---|---|---|---|---|---|---|
| **Systematic Review** | **№ of studies** | **Study design** | **Risk of bias** | **Inconsistency** | **Indirectness** | **Imprecision** | **Other considerations** | |
| **Scarpone et al. 2023** | **4** | RCT | Not serious; Most information is from studies at low risk of bias or some concerns for one domain only. | Serious; $I^2$=49 %; relatively small difference in RR in one large study (Perkin et al. 2016) appears to explain heterogeneity, but the reason for the different findings in this study is not clear. | Not serious; Populations and interventions are similar to those which might be targeted outside of trial settings. Some studies used food powder, and some used normal foods. One study included infants with eczema. | Not serious; The 95% CI excludes a RR of no effect or effect sizes that are not clinically meaningful. | Insufficient studies to undertake formal testing of publication bias | Introduction of multiple allergenic foods |
| **Scarpone et al. 2023** | **4** | RCT | Not serious; Most information is from studies at low risk of bias or some concerns for one domain only. | Serious; $I^2$=45 %; relatively small difference in RR in one large study (Perkin et al. 2016) appears to explain heterogeneity, but the reason for the different findings in this study is not clear. | Very serious; Almost all the information for this analysis is derived from trials of multiple food interventions. The limited information for egg only shows 6/15 children developed food allergy, compared with 21/44 in a control group. There is therefore little meaningful information about the effect of earlier egg introduction on risk of allergy to any food. | Not serious; The 95% CI excludes a RR of no effect or effect sizes that are not clinically meaningful. | Insufficient studies to undertake formal testing of publication bias | Introduction of egg |

| | Certainty assessment | | | | | | | Comments |
|---|---|---|---|---|---|---|---|---|
| **Systematic Review** | **№ of studies** | **Study design** | **Risk of bias** | **Inconsistency** | **Indirectness** | **Imprecision** | **Other considerations** | |
| **Scarpone et al. 2023** | **5** | RCT | Not serious; Most or all information is from studies at low risk of bias or some concerns for one domain only. | Serious; $I^2$=81 %; heterogeneity appears to be partly (but not wholly) explained by a lack of effect in the single peanut-only intervention trial but reduced allergy in the multiple food intervention trials. This interaction was statistically significant (p=0.02). | Serious; Some data contributing to this effect estimate are from trials of multiple allergenic food introduction. The single trial of peanut only shows no effect. Populations varied in risk factors such as eczema at baseline, in whether and how peanut allergy was excluded prior to the intervention and in the nature of the intervention. | Serious; The 95% CI includes both a meaningful reduction and a trivial reduction or small increase in RR. | Insufficient studies to undertake formal testing of publication bias | Introduction of peanut |
| **Scarpone et al. 2023** | **6** | RCT | Not serious; Most information is from studies at high risk of bias, but findings appear to be similar to studies at low risk of bias or some concerns for one domain only. | Serious; $I^2$=83 %; heterogeneity appears to be partly, but not completely, explained by one high risk of bias study. | Serious; Interventions were not all representative of typical ways to introduce cow's milk products to an infant's diet. Some studies used food powder, others used formula milk compared with a control group using soya formula, but often using very early and specific exposure regimes. | Serious; The 95% CI includes both a meaningful reduction and increase in risk or no effect | Insufficient studies to undertake formal testing of publication bias | Introduction of cow's milk;<br>In one further high risk of bias study which could not be included in meta-analysis, none of 120 children who were given cow's milk (formula) and none of 115 children who were given a soya formula for the first 9 months of life developed allergy to foods such as soya, fish, orange, and nuts, and one child became clinically sensitive to chocolate after a 10-year follow-up. |

| | Certainty assessment | | | | | | | Comments |
|---|---|---|---|---|---|---|---|---|
| **Systematic Review** | **№ of studies** | **Study design** | **Risk of bias** | **Inconsistency** | **Indirectness** | **Imprecision** | **Other considerations** | |
| **Scarpone et al. 2023** | **4** | RCT | Serious; Information is from two studies at low risk of bias and one study with some concerns, but one study was at high risk due to bias of the randomisation process, measurement of the outcome and selection of the reported results. | Serious; $I^2$=72 %; no further anylses conducted to explore heterogeneity. | Not serious; Populations are similar to the target population and trials were conducted in the general population (three trials) and in a high-risk population (one trial). However, multiple foods (both food powder and normal foods) were introduced so it is diffucult to draw causal conclusions about the risk of developing wheat allergy by the introduction of wheat. | Not serious; The 95% CI excludes a RR of no effect | None | Introduction of wheat |
| **Scarpone et al. 2023** | **2** | RCT | Serious; Information is from one study at low risk of bias and one study at hish risk of bias due to bias of the randomisation process, measurement of the outcome and selection of the reported results. | Serious; $I^2$=88 %; no further anylses conducted to explore heterogeneity. | Serious; The studies varied in baseline risks with one study that was conducted in the general population and one study in infants at high-risk (who have atopic dermatitis). | | None | Introduction of soy |
| **Scarpone et al. 2023** | **2** | RCT | Serious; Information is from one study with some concers for the risk of bias and one study at hish risk of bias due to bias of the randomisation process, measurement of the outcome and selection of the reported results. | Serious; $I^2$=88 %; no further anylses conducted to explore heterogeneity. | Not serious; Both studies were conducted in the general population which is thus similar to the target population, the intervention consisted of introducing multiple foods | Serious; The 95% CI includes no effect, clinical meaningful harm and benefit | None | Introduction of fish |

| | Certainty assessment | | | | | | | Comments |
|---|---|---|---|---|---|---|---|---|
| **Systematic Review** | **№ of studies** | **Study design** | **Risk of bias** | **Inconsistency** | **Indirectness** | **Imprecision** | **Other considerations** | |
| **Scarpone et al. 2023** | **1** | RCT | Not serious; low risk of bias for all domains | Not applicable (only one underlying primary study) | Not serious; The study was conducted in the general population with an intervention consisting of introducing multiple foods | Very serious; The number of participants and events is low | None | Introduction of crustaceans or tree nuts |
| **Risk of egg allergy** | | | | | | | | |
| **Al-Saud et al. 2018** | **6** | RCT | Not serious | Not serious | Not serious | Not serious | None | |
| **Burgess et al. 2019** | **6** | RCT | Not serious | Not serious; $I^2$= 40.6% (p=0.134) | Serious; two trials used placebo control groups, which allows no direct conclusions about effects of the timing of introducing CFs | Not serious | None | The certainty of the evidence was downgraded two levels due to serious concerns in RoB and indirectness |
| **Dai et al. 2019** | **6** | RCT | Not serious | Not serious | Serious; two trials used placebo control groups, which allows no direct conclusions about effects of the timing of introducing CFs | Not serious; one study with a very low number of events introduces heterogeneity. However, the confidence interval is narrow and the number of participants is high | None | |
| **De Silva et al. 2020** | **1** | RCT | Not serious | Not serious | Serious; very small amounts of egg; rather comparable to immunotherapy approach in reference to S3 Table of the primary SR | Not serious | None | Introduction of cooked egg; the certainty of evidence was downgraded two levels due to serious concerns in indirectness and imprecision |
| **De Silva et al. 2020** | **1** | RCT | Not serious | Serious; variations depending in quantity/ frequency; variations in risk profil in reference to S3 Table of the primary SR | Serious; drop out due to adverse events in reference to S3 Table of the primary SR | Not serious | None | Introduction of raw pateurized egg; the certainty of evidence was downgraded three levels due to serious concerns in RoB, inconsistency and indirectness |

| | Certainty assessment | | | | | | | Comments |
|---|---|---|---|---|---|---|---|---|
| **Systematic Review** | **№ of studies** | **Study design** | **Risk of bias** | **Inconsistency** | **Indirectness** | **Imprecision** | **Other considerations** | |
| **EFSA Panel** | | RCT | Not serious | Not serious | Very serious; some trials used placebo control groups, which allows no direct conclusions about effects of the timing of introducing CFs | Not serious | | |
| **Ierodiakonou et al. 2016** | **6** | RCT | Very serious; the majority of RoB assessments resulted in "high" | Not serious; $I^2$ = 35.8% (p=0.1829) | Serious; according to the primary SR three trials recruited only infants without egg sensitisation; one trial only infants with eczema; one trial used multiple allergenic foods (p. 1187) | Not serious | None | The certainty of evidence was downrated three levels because of very serious concerns regarding risk of bias and serious concerns regarding indirectness |
| **Larson et al. 2017** | **1** | RCT | Not serious | Not applicable (only one underlying primary study) | Serious; the trial was conducted in a high-risk study population only and was performed in Australia, a country with an unexplained higher prevalence of allergic diseases | Serious; the effect estimate comes from only one small study, the 95% CI includes no effect; the number of events is low | None | Ther certainty of evidence was downgraded two levels due to serious concerns regarding indirectness, and imprecision;<br>the evidence is based on a single study, assessing the domain „inconsistency“ is therefore not applicable (4/5 domains considered) |

| | Certainty assessment | | | | | | | Comments |
|---|---|---|---|---|---|---|---|---|
| Systematic Review | № of studies | Study design | Risk of bias | Inconsistency | Indirectness | Imprecision | Other considerations | |
| **Scarpone et al. 2023** | **9** | RCT | Not serious; Most information is from studies at low risk of bias or some concerns for one domain only. | Not serious; $I^2$=0%; mostly similar RRs and overlapping CIs. No obvious difference in outcome between single and multiple allergenic food introduction. Borderline interaction for high versus low dose egg introduction (P=0.02), with increased effect in low dose group. | Not serious; Populations and interventions are similar to those which might be targeted outside of trial settings. Some studies used food powder, but findings were similar to studies using normal foods. Populations varied in risk factors such as eczema at baseline, in whether and how egg allergy was excluded prior to the intervention and in the nature of the intervention. | Not serious; The 95% CI excludes a RR of no effect or effect sizes that are not clinically meaningful. | Insufficient studies to undertake formal testing of publication bias | |
| **Risk of peanut allergy** | | | | | | | | |
| **Burgess et al. 2019** | **2** | RCT | Serious; all of the RoB assesments resulted in "some concerns" | Not serious; $I^2$= 43.5% (p= 0.170) | Serious; two trials used placebo or avoidance control groups, which allows no direct conclusions about effects of the timing of introducing CF and the study population of one study was at high risk | Not serious | None | The certianty of evidence was downgraded two levels due to serious concerns in RoB and indirectness |
| **Dai et al. 2019** | **2** | RCT | Serious; one study was at high risk of bias due to high risk regarding outcome measurement, the other study's assessment resulted in "some concerns" | | Serious; one study population consisted of infants at risk for allergy; one study introduced multiple allergenic foods as intervention | | None | |

| | Certainty assessment | | | | | | | Comments |
|---|---|---|---|---|---|---|---|---|
| Systematic Review | № of studies | Study design | Risk of bias | Inconsistency | Indirectness | Imprecision | Other considerations | |
| **De Silva et al. 2020** | **2** | RCT | Serious; the majority of RoB assessements resulted in "some concerns" | Not serious | Very serious; very high risk infants in high prevalence country; comparison group had total abstinence from peanut for five years rather than usual diet according to the S3 Table in the primary SR | Not serious | None | The certainty of evidence was downgraded three levels due to serious concerns in RoB, and very serious concerns regarding indirectness |
| **EFSA Panel 2019** | **1** | RCT | Serious; the RoB dimension "Measurement of the outcome" resulted in "high" | Not applicable (only one underlying primary study) | Serious; the study population consisted of breastfed infants only; the authors of the primary SR considered that the study results cannot be generalised to formula fed infants and thus to the whole population of infants living in Europe (EFSA Panel et al 2019, p. 79ff) | Serious; the effect estimate comes from one single study; wide CI including no effect | None | The certainty of evidence was downgraded three levels due to serious concerns regarding risk of bias, indirectness, and imprecision;<br>the evidence is based on a single study, assessing the domain „inconsistency" is therefore not applicable (4/5 domains considered) |
| **Ierodiakonou et al. 2016** | **2** | RCT | Serious; the majority of RoB assessements resulted in "some concerns" | Not serious; $I^2$ = 66.1 % (p =0.0857) | Serious; one study recruited only infants with egg allergy or eczema and without high-level peanut sensitisation; one study used multiple allergenic foods (Ierodiakonou et al. 2016, p. 1187) | Serious; wide 95% CI for RR | None | The certainty of evidence was downrated three levels due to serious concers regarding risk of bias, indirectness and imprecision |

| | Certainty assessment | | | | | | | Comments |
|---|---|---|---|---|---|---|---|---|
| **Systematic Review** | **№ of studies** | **Study design** | **Risk of bias** | **Inconsistency** | **Indirectness** | **Imprecision** | **Other considerations** | |
| **Larson et al. 2017** | **1** | RCT | Serious; the RoB assessment resulted in "some concerns" in three out of five dimensions | Not applicable (only one underlying primary study) | Very serious; the study population consisted of high risk infants and comes from only one trial, the trial was conducted in a country with a high prevalence of atopic diseases and the control group had complete avoidance of CF instead of delayed introduction of CF | Not serious | None | The certainty of evidence was downgraded three levels due to serious concerns regarding RoB, and very serious concerns regarding indirectness;<br>the evidence is based on a single study, assessing the domain „inconsistency“ is therefore not applicable (4/5 domains considered) |
| **Scarpone et al. 2013** | **4** | RCT | Not serious; Most or all information is from studies at low risk of bias or some concerns for one domain only. | Not serious; $I^2$=21 %; mostly similar RRs and overlapping CIs. | Not serious; Most of the events contributing to this effect estimate are from a trial of single allergenic food introduction. Populations varied in risk factors such as eczema at baseline, in whether and how peanut allergy was excluded prior to the intervention and in the nature of the intervention. | Not serious; The 95% CI excludes a RR of no effect or effect sizes that are not clinically meaningful. | Insufficient studies to undertake formal testing of publication bias | |

| | Certainty assessment | | | | | | | Comments |
|---|---|---|---|---|---|---|---|---|
| Systematic Review | № of studies | Study design | Risk of bias | Inconsistency | Indirectness | Imprecision | Other considerations | |
| **Risk of grain allergy** | | | | | | | | |
| **Chmielewska et al. 2017** | **1** | RCT | Not serious | Not applicable (only one underlying primary study) | Serious; trial was performed in a country with study population at high risk but in the general population; the intervention condition consisted of introducing six allergenic foods. Therefore, the intervention effect cannot be attributed to one specific underlying mechanism and may not be representative for the general population. | Serious; CI is not reported in the primary SR; however, the number of participants is high (N>1000) but the event rate is very low | None | The certainty of evidence was downgraded two levels due to serious concerns in indirectness and imprecision;<br>the evidence is based on a single study, assessing the domain „inconsistency“ is therefore not applicable (4/5 domains considered) |

| | Certainty assessment | | | | | | | Comments |
|---|---|---|---|---|---|---|---|---|
| Systematic Review | № of studies | Study design | Risk of bias | Inconsistency | Indirectness | Imprecision | Other considerations | |
| **EFSA Panel 2019** | **1** | RCT | Serious; the ROB assessment resulted in "some concerns" in three out of five dimensions | Not applicable (only one underlying primary study) | Serious; the study population consisted of breastfed infants only; the authors of the primary SR considered that the study results cannot be generalised to formula fed infants and thus to the whole population of infants living in Europe (EFSA Panel et al. 2019, p.86) | Not serious | None | Intervention: Introduction of cereals (wheat); the certainty of evidence was downgraded two levels due to serious concerns in risk of bias, and serious concerns regarding indirectness;<br>the evidence is based on a single study, assessing the domain „inconsistency“ is therefore not applicable (4/5 domains considered) |
| **Scarpone et al. 2023** | **3** | RCT | Not serious; Two studies were at low risk of bias, one study was rated with some concerns due to bias in measurement of the outcome | Not serious; $I^2$=2 %. | Serious; All studies were conducted in the general population; but the interventions consisted of introducing multiple foods | Very serious; the 95% CI includes no effect and clinically meaningful harm | None | Intervention: Introduction of wheat |

| | Certainty assessment | | | | | | | Comments |
|---|---|---|---|---|---|---|---|---|
| **Systematic Review** | **№ of studies** | **Study design** | **Risk of bias** | **Inconsistency** | **Indirectness** | **Imprecision** | **Other considerations** | |
| **Risk of cow's milk allergy** | | | | | | | | |
| **Dai et al. 2019** | **2** | RCT | Serious; one study was at high risk of bias due to high risk regarding outcome measurement, the other study's assessment resulted in "some concerns" | | Serious; one study population consisted of infants at high risk only and was conducted in Austria, a country with an unexplained higher prevalence of allergic diseases; one trial consisted of a study population of exclusively breastfed infants | | | |
| **Ierodiakonou et al. 2016** | **2** | RCT | Serious; the majority of the RoB assessements resulted in "some concerns". | Not serious; $I^2 = 0\%$ (p=0.998) | Serious; one trial was conducted in a high-risk study population only; and was performed in Australia, a country with an unexplained higher prevalence of allergic diseases; one trial consisted of a study population of breastfed infants only | Serious; the 95% CI for the pooled risk ratio is wide, indicating that future interventions could include potential benefits and harms | None | The certainty of evidence was downrated three levels due to serious concerns regarding risk of bias, indirectness and imprecision |
| **Scarpone et al. 2023** | **6** | RCT | Serious; Most information is from studies at high risk of bias for at least one domain and/or some concerns for multiple domains, with high heterogeneity in the high risk of bias information. The main reason for risk of bias is that outcome assessors were not blinded. | Not serious; $I^2$=36%; heterogeneity appears to be explained by risk of bias, with low risk of bias studies showing no heterogeneity and RR 0.32 (95% CI, 0.09-1.18). | Serious; Interventions were not all representative of typical ways to introduce cow's milk products to an infant's diet. Some studies used food powder, others used formula milk compared with a control group using soya formula, but often using very early and specific exposure regimes. | Serious; The 95% CI includes both a meaningful reduction and increase in risk or no effect. | Insufficient studies to undertake formal testing of publication bias | One further high risk of bias trial[48] reported milk allergy in 2/242 in the earlier and 17/249 in the later introduction group, assessed at 6 months, so could not be included in meta-analysis. |

| | Certainty assessment | | | | | | | Comments |
|---|---|---|---|---|---|---|---|---|
| Systematic Review | № of studies | Study design | Risk of bias | Inconsistency | Indirectness | Imprecision | Other considerations | |
| **Other common allergenic foods** | | | | | | | | |
| **Scarpone et al. 2023** | **1** | RCT | Not serious; All domains were rated at low risk of bias | Not applicable (only one underlying primary study) | Serious; The study was conducted in a population with high-risk of allergies (diagnosed atopic dermatitis), the intervention consisted of introducing multiple allergenic foods | Very serious; The number of participants was low and the number of events was very low | None | Outcome: risk of developing soya allergy |
| **Scarpone et al. 2023** | **1** | RCT | Not serious; all but one domain were rated at low risk of bias (measurement of the outcome with some concerns) | Not applicable (only one underlying primary study) | Serious; The study was conducted in the general population, the intervention consisted of introducing multiple allergenic foods | Serious; the number of events was very low | None | Outcome: risk of developing fish allergy |
| **Eczema** | | | | | | | | |
| **Al-Saud et al. 2018** | **2** | RCT | Not serious | Not serious; low heterogeneity, $I^2$ = 0% (p=0.99) | Not serious | Serious;the 95% CI for the pooled RR is wide | None | The certainty of evidence was downgraded for ROB (high concerns) and imprecision (95% CI for the pooled RR is wide) |

| | Certainty assessment | | | | | | | Comments |
|---|---|---|---|---|---|---|---|---|
| **Systematic Review** | **№ of studies** | **Study design** | **Risk of bias** | **Inconsistency** | **Indirectness** | **Imprecision** | **Other considerations** | |
| **EFSA Panel 2019** | **2** | RCT | Not serious | Not serious; $I^2$ = 69% (p=0.07) | Very serious; both trials were performed in at-risk study populations in Australia, a country that has an unexplained higher prevalence of allergy than Europe; one the study used pasteurised raw egg powder as an intervention product, which is not the form that would be used when egg is introduced to infants (p. 73) | Serious; the effect estimate comes from only two trials, very wide CI; low event rate; the sample size of each trial is moderate to high | None | Intervention: introduction of egg; the certainty of evidence was downgraded three levels due to serious concerns regarding RoB and imprecision, and very serious concerns regarding indirectness |
| **Ierodiakonou et al. 2016** | **8** | RCT | Very serious; the majority of RoB assessment resulted in "high" | Serious; the amount of heterogeneity quantified via $I^2$ is moderate (ca. 54%) and significant; the point estimates range from 0.54 - 3.63 with partly non-overlapping CIs | Serious; multiple studies examined the effects of multifaceted interventions; control group comparisons consisted of placebo or avoidance in some cases instead of delaying the food introduction | Not serious | None | Outcome measured at age ≤ 4 years; the certainty of evidence was downgraded three levels due to very serious concerns regarding RoB and serious concerns regarding inconsistency and indirectness |
| **Ierodiakonou et al. 2016** | **6** | RCT | Very serious; the majority of RoB assessment resulted in "high" | Not serious | Serious; multiple studies examined the effects of multifaceted interventions; control group comparisons consisted of placebo or avoidance in some cases instead of delaying the food introduction | Not serious | None | Outcome measured at age 5-15 years; the certainty of evidence was downgraded three levels due to very serious concerns regarding RoB and serious concerns regarding indirectness |

| | Certainty assessment | | | | | | | Comments |
|---|---|---|---|---|---|---|---|---|
| **Systematic Review** | **№ of studies** | **Study design** | **Risk of bias** | **Inconsistency** | **Indirectness** | **Imprecision** | **Other considerations** | |
| **Smith et al. 2016** | **1** | RCT | Not serious | Not applicable (only one underlying primary study) | Not serious | Serious; wide CI crossing the line of no effect and small sample size (p.8) | None | The certainty of evidence was downgraded one levels due to serious concerns regarding imprecision;<br>the evidence is based on a single study, assessing the domain „inconsistency“ is therefore not applicable (4/5 domains considered for assessing the certainty level) |
| **Waidyatillake et al. 2019** | **2** | RCT | Not serious | Not serious | Serious; both trials are conducted in Australia, which is a country with an unexplainable increased risk of atopic diseases; in both interventions egg powder was introduced, which is a form that can not necessarily be compared to formula introduction. | Serious; CI included potential for harm and benefit; the overall sample size is high with N>1000 but the number of events is low | None | The certainty of evidence was downgraded three levels due to serious concerns in RoB, indirectness and imprecision |
| **Allergic Rhinitis** | | | | | | | | |
| **EFSA Panel 2019** | **1** | RCT | Not applicable (information beyond the effect estimate provided in the EFSA Panel report is not publicly available to the overview authors, whereas the review authors contacted Perkin et al. 2016 to obtain more information) | Not applicable (only one underlying primary study) | Not applicable (information beyond the effect estimate provided in the EFSA Panel report is not publicly available to the overview authors, whereas the review authors contacted Perkin et al. 2016 to obtain more information) | Serious; the effect estimate comes from only one trial; wide CI; low event rate; the sample size of each study is moderate to high | None | Three of five domains not assessible (“inconsistency” not applicable for one study |

| | Certainty assessment | | | | | | | Comments |
|---|---|---|---|---|---|---|---|---|
| Systematic Review | № of studies | Study design | Risk of bias | Inconsistency | Indirectness | Imprecision | Other considerations | |
| **Ierodiakonou et al. 2016** | **6** | RCT | Very serious; the majority of RoB assessment resulted in "high" | Serious; the amount of heterogeneity quantified via the $I^2$ value is moderate to high (ca. 70%) and significant; point estimates range from 0.23 - 9.58 with a high variance of point estimates | Serious; multiple studies examined the effects of multifaceted interventions; control group comparisons consisted of placebo or avoidance in some cases instead of delaying the food introduction | Not serious | None | Outcome measured at ≤4 years of age; the certainty of evidence was downgraded three levels due to very serious concerns regarding RoB and serious concerns regarding inconsistency and indirectness |
| **Ierodiakonou et al. 2016** | **5** | RCT | Very serious; all of RoB assesments resulted in "high" | Serious; the amount of heterogeneity quantified via the $I^2$ value is moderate to high (ca. 79%) and significant; point estimates range from 0.83-4.79 with a high variance of point estimates | Serious; multiple studies examined the effects of multifaceted interventions; control group comparisons consisted of placebo or avoidance in some cases instead of delaying the food introduction | Not serious | None | Outcome measured between 5-14 years of age; the certainty of evidence was downgraded three levels due to very serious concerns regarding RoB and serious concerns regarding inconsistency and indirectness |
| **Allergic asthma, wheeze or asthma-like symptoms** | | | | | | | | |
| **EFSA Panel 2019** | **1** | RCT | Serious; the RoB dimension "Measurement of the outcome" resulted in "high" | Not applicable | Serious; the study population consisted only of breastfed infants; the authors of the primary SR considered that the results of this study cannot be generalised to formula fed infants and thus to the whole population of infants living in Europe (p. 67) | Serious; the effect estimate comes from only one trial; wide 95% CI; low number of events, but the sample size is high (N >1000) | None | Introduction of CF in general;<br>evidence is based on a single study, assessing the domain „inconsistency" is therefore not applicable (4/5 domains considered);<br>the certainty of evidence was downgraded two levels due to serious concerns in RoB, indirectness and imprecision |

| | Certainty assessment | | | | | | | Comments |
|---|---|---|---|---|---|---|---|---|
| Systematic Review | № of studies | Study design | Risk of bias | Inconsistency | Indirectness | Imprecision | Other considerations | |
| **EFSA Panel 2019** | **1** | RCT | Not serious | Not applicable | Serious; the study population consisted of breastfed infants only; the authors of the primary SR considered that the study results cannot be generalised to formula fed infants and thus to the whole population of infants living in Europe (EFSA Panel et al. 2019, p. 79ff) | Serious; the effect estimate comes from one single study; wide 95% CI including no effect | None | Introduction of egg;<br>evidence is based on a single study, assessing the domain „inconsistency“ is therefore not applicable (4/5 domains considered);<br>the certainty of evidence was downgraded two levels due to very serious concerns in indirectness and serious concerns in imprecision; |
| **Atopic diseases (Outcome-Cluster)** | | | | | | | | |
| **EFSA Panel 2019** | **2** | RCT | Very serious; Both RoB assessments for each trial resulted in "high" | Not applicable | Serious; the study population of the RCT available in the general population consisted only of breastfed infants; the authors of the primary SR considered that the results of this study cannot be generalised to formula fed infants and thus to the whole population of infants living in Europe (p. 65) | Not serious | None | Introduction of CF in general;<br>evidence is based on a single study, assessing the domain „inconsistency“ is therefore not applicable (4/5 domains considered);<br>the certainty of evidence was downgraded two levels due to serious concerns in RoB, indirectness and imprecision<br>For the effects of introducing egg in specific, the certainty of the evidence was not further considered in EFSA Panel et al. 2019 as the authors deemed the evidence basis was too sparse to draw valid conclusions |

| | Certainty assessment | | | | | | | Comments |
|---|---|---|---|---|---|---|---|---|
| **Systematic Review** | **№ of studies** | **Study design** | **Risk of bias** | **Inconsistency** | **Indirectness** | **Imprecision** | **Other considerations** | |
| **Incidence of adverse events (ADE), severe adverse events (SADE), and withdrawals , due to ADEs or SADEs** | | | | | | | | |
| **Scarpone et al. 2023** | **4** | RCT | Serious; Most information is from studies at high risk of bias for one domain or some concerns for multiple domains. The main reason for risk of bias is that withdrawal could have been influenced by knowledge of intervention received | Not serious; $I^2$=89%; heterogeneity appears to be explained by lower compliance in two largest studies (both high allergen intake for multiple allergenic foods, using normal foods rather than powders). | Not serious; Populations and interventions are similar to those which might be targeted outside of trial settings. Some studies used food powder, and some used normal foods. One study included infants with eczema. | Not serious; The 95% CI excludes a RR of no effect or effect sizes that are not clinically meaningful. | Insufficient information to undertake formal testing of publication bias | Introduction of multiple allergic foods on withdrawals of study interventions |
| **Scarpone et al. 2023** | **13** | RCT | Not serious; Most information is from studies at high risk of bias, but findings appear to be similar to studies at low risk of bias or some concerns for one domain only. | Serious; $I^2$=90%; heterogeneity appears to be explained by lower compliance in two large, pragmatic studies of high-dose multiple allergenic food introduction using normal foods, together with reports of allergic reactions to egg intervention in some smaller, single-food intervention trials. | Not serious; Populations and interventions are similar to those which might be targeted outside of trial settings. Some studies used food powder, but findings were similar to studies using normal foods. Populations varied in risk factors such as eczema at baseline, in whether and how egg allergy was excluded prior to the intervention and in the nature of the intervention. | Serious; The 95% CI excludes a RR of no effect but includes effect sizes that are both trivial or uncertain and clinically meaningful. | Not likely; Funnel plot asymmetrical and Egger's test P=0.004; however, likely due to lower compliance in larger studies of multiple, high-dose allergenic food introduction, compared with smaller studies of single allergen (egg) introduction. | Introduction of egg on withdrawals of study interventions |

| | Certainty assessment | | | | | | | Comments |
|---|---|---|---|---|---|---|---|---|
| Systematic Review | № of studies | Study design | Risk of bias | Inconsistency | Indirectness | Imprecision | Other considerations | |
| **Scarpone et al. 2023** | **6** | RCT | Serious; Most information is from studies at high risk of bias for one domain or some concerns for multiple domains. The main reason for risk of bias is that withdrawal could have been influenced by knowledge of intervention received | Serious; $I^2$=91%; heterogeneity appears to be partly explained by lower compliance in two large, pragmatic studies of high-dose multiple allergenic food introduction using normal foods. | Serious; Some data contributing to this effect estimate are from trials of multiple allergenic food introduction. The single trial of peanut only shows no effect. Populations varied in risk factors such as eczema at baseline, in whether and how peanut allergy was excluded prior to the intervention and in the nature of the intervention. | Not serious; The 95% CI excludes a RR of no effect or effect sizes that are not clinically meaningful. | Insufficient information to undertake formal testing of publication bias | Introduction of peanut on withdrawals of study interventions |
| **Scarpone et al. 2023** | **11** | RCT | Not serious; Most information is from studies at high risk of bias, but findings appear to be similar to studies at low risk of bias or some concerns for one domain only. | Not serious; $I^2$=94%; heterogeneity is extreme and is fully explained by high rates of withdrawal in two pragmatic trials of multiple, high-dose, real food intervention trials, and a trial where many participants withdrew from the soya milk intervention due to a preference for cow's milk. Without these three trials included, the RR is 0.86 (95% CI, 0.70-1.06; $I^2$=0%). | Serious; Interventions were not all representative of typical ways to introduce cow's milk products to an infant's diet. Some studies used food powder, others used formula milk compared with a control group using soya formula, but often using very early and specific exposure regimes. | Serious; The 95% CI includes both a meaningful reduction and increase in risk or no effect. | Not likely; Funnel plot asymmetrical and Egger's test P=0.01; however, likely due to lower compliance in larger studies of multiple, high-dose allergenic food introduction, compared with smaller studies of single allergen (milk) introduction. | Introduction of cow's milk on withdrawals of study interventions;<br>In one further high risk of bias trial which could not be included in meta-analysis, 32/249 participants in the cow's milk group were given soya milk and 41/238 participants in the cow's milk avoidance group were given cow's milk. However, total number of participants randomized in each group was not clear as these numbers excluded some post-randomization exclusions and the number of events in the cow's milk avoidance group was inconsistent between study publications. |
| **Smith et al. 2016** | **1** | RCT | Serious; Quasi-random sequence generation (by week of birth) | Not serious | Not serious | Serious; Wide confidence interval crossing the line of no effect and small sample size | Not reported | Outcome: Fever (% of days) |

| | Certainty assessment | | | | | | | Comments |
|---|---|---|---|---|---|---|---|---|
| Systematic Review | № of studies | Study design | Risk of bias | Inconsistency | Indirectness | Imprecision | Other considerations | |
| **Smith et al. 2016** | **2** | RCT | Serious; Quasi-random sequence generation (by week of birth) | Not serious | Not serious | Serious; Wide confidence interval crossing the line of no effect and small sample size | Not reported | Outcome: weight change (gain) (g) |
| | **1** | RCT | Not serious | Not serious | Not serious | Serious; Estimate based on small sample size (-1) | Not reported | Outcome: weight change (z score) |
| | **1** | RCT | Not serious | Not serious | Not serious | Serious; wide confidence interval crossing the line of no effect and small sample | Not reported | Outcome: gastrointestinal events (diarrhoea, vomiting, constipation), colic, enterocolitis syndrome reactions |
| **Allergic sensitisation** | | | | | | | | |
| **Al-Saud et al. 2018** | **5** | RCT | Serious; Allocation sequence concealment was unclear in 2 studies and 1 study was not blinded. | Not serious; low heterogeneity, $I^2$= 17% (*p*= 0.31) | Not serious; allergy sensitisation is a surrogate marker for food allergy | Not serious; the 95% CI for the pooled RR is 0.58-0.95 | Not reported | |
| **Scarpone et al. 2023** | **3** | RCT | Not serious; All information is from studies at low risk of bias. | Serious; $I^2$=73%; one study (Skjerven et al, 2022) which used skin prick test (SPT) ≥3mm showed reduced sensitisation and two studies which used SPT >0mm and specific immunoglobulin E test showed no effect. | Not serious; Populations and interventions are similar to those which might be targeted outside of trial settings. Some studies used food powder, and some used normal foods. One study included infants with eczema. | Serious; Although the estimate indicates reduced allergic sensitisation, the 95% CI includes the possibility of no effect. | Insufficient studies to undertake formal testing of publication bias | Intervention: introducing *multiple allergenic foods* (intervention foods were egg, milk, wheat, soya, buckwheat, and peanut (1 trial; 150 participants); milk, peanut, egg, sesame, white fish, and wheat (1 trial; 1173 participants); peanut, milk, wheat, and egg (1 trial; 1504 participants))<br>Outcome: allergic sensitisation *to any food* |

| | Certainty assessment | | | | | | | Comments |
|---|---|---|---|---|---|---|---|---|
| Systematic Review | № of studies | Study design | Risk of bias | Inconsistency | Indirectness | Imprecision | Other considerations | |
| **Scarpone et al. 2023** | **3** | RCT | Not serious; All information is from studies at low risk of bias. | Serious; $I^2$=73%; one study (Skjerven et al, 2022) which used skin prick test (SPT) ≥3mm showed reduced sensitisation and two studies which used SPT >0mm and specific immunoglobulin E test showed no effect. | Not serious; Populations and interventions are similar to those which might be targeted outside of trial settings. Some studies used food powder, and some used normal foods. One study included infants with eczema. | Serious; Although the estimate indicates reduced allergic sensitisation, the 95% CI includes the possibility of no effect. | Insufficient studies to undertake formal testing of publication bias | Intervention: introducing egg<br>Outcome: risk of allergic sensitisation *to any food*<br>Results as for "earlier introduction of multiple allergenic foods"; no study of egg introduction without other foods, reporting this outcome, was identified. |
| | **8** | RCT | Not serious; Most information is from studies at low risk of bias or some concerns for one domain only. | Not serious; $I^2$=18%; mostly similar RRs and overlapping CIs. Post hoc subgroup analysis showed a possible interaction for method of assessment (P=0.06): skin prick test (5 studies; 3740 participants) RR 0.72 (95% CI 0.58-0.89; $I^2$=0%); specific immunoglobulin E test (3 studies; 585 participants) RR 0.92 (95% CI, 0.80-1.04; $I^2$=43%). | Not serious; Populations and interventions are similar to those which might be targeted outside of trial settings. Some studies used food powder, but findings were similar to studies using normal foods. Populations varied in risk factors such as eczema at baseline, in whether and how egg allergy was excluded prior to the intervention and in the nature of the intervention. | Serious; The 95% CI excludes a RR of no effect but includes effect sizes that are both trivial or uncertain and clinically meaningful. | Insufficient studies to undertake formal testing of publication bias | Intervention: introducing egg (Intervention foods were egg (5 trials; 1504 participants); egg, milk, wheat, soya, buckwheat, and peanut (1 trial; 150 participants); milk, peanut, egg, sesame, white fish, and wheat (1 trial; 1167 participants); peanut,milk, wheat, and egg (1 trial; 1504 participants))<br>Outcome: risk of allergic sensitisation *to egg* |

| | Certainty assessment | | | | | | | Comments |
|---|---|---|---|---|---|---|---|---|
| **Systematic Review** | **№ of studies** | **Study design** | **Risk of bias** | **Inconsistency** | **Indirectness** | **Imprecision** | **Other considerations** | |
| **Scarpone et al. 2023** | **4** | RCT | Not serious; Most or all information is from studies at low risk of bias or some concerns for one domain only. | Serious; $I^2$=61%; one study (Skjerven et al, 2022) of multiple allergenic food introduction which used skin prick test (SPT) ≥3mm showed reduced sensitisation and appears to explain the heterogeneity, but the reason for different findings in this trial is unclear. | Not serious; Most of the events contributing to this effect estimate are from a trial of single allergenic food introduction. Populations varied in risk factors such as eczema at baseline, in whether and how peanut allergy was excluded prior to the intervention and in the nature of the intervention. | Serious; The 95% CI includes both a meaningful reduction and a trivial reduction or small increase in RR. | Insufficient studies to undertake formal testing of publication bias | Intervention: introducing peanut (Intervention foods were peanut (1 trial; 629 participants); egg, milk, wheat, soya, buckwheat, and peanut (1 trial; 150 participants); milk, peanut, egg, sesame, white fish, and wheat (1 trial; 1173 participants); peanut, milk, wheat, and egg (1 trial; 1504 participants))<br><br>Outcome: sensitisation *to any food* |
| | **4** | RCT | Not serious; Most or all information is from studies at low risk of bias or some concerns for one domain only. | Serious; $I^2$=77 %; one study (Skjerven et al. 2022) of multiple allergenic food introduction which used SPT ≥3mm showed reduced sensitisation and appears to explain the heterogeneity, but the reason for different findings in this trial is unclear. Post hoc subgroup analysis showed a possible interaction for method of assessment (p=0.06): SPT (2 studies; 2672 participants) RR 0.46 (95% CI, 0.20-1.05; $I^2$=72 %); specific immunoglobulin E test (2 studies; 762 participants) RR 1.07 (95% CI, 0.85-1.34; $I^2$=0 %). | Not serious; Most of the events contributing to this effect estimate are from a trial of single allergenic food introduction. Populations varied in risk factors such as eczema at baseline, in whether and how peanut allergy was excluded prior to the intervention and in the nature of the intervention. | Serious; The 95% CI includes both a meaningful reduction and a trivial reduction or small increase in RR. | Insufficient studies to undertake formal testing of publication bias | Intervention: introducing peanut (Intervention foods were peanut (1 trial; 617 participants); egg, milk, wheat, soya, buckwheat, and peanut (1 trial; 145 participants); milk, peanut, egg, sesame, white fish, and wheat (1 trial; 1168 participants); peanut, milk, wheat, and egg (1 trial; 1504 participants))<br><br>Outcome: sensitisation *to peanut* |

| | Certainty assessment | | | | | | | Comments |
|---|---|---|---|---|---|---|---|---|
| Systematic Review | № of studies | Study design | Risk of bias | Inconsistency | Indirectness | Imprecision | Other considerations | |
| **Scarpone et al. 2023** | **3** | RCT | Not serious; All information is from studies at low risk of bias. | Serious; $I^2$=73%; one study (Skjerven et al, 2022) which used skin prick test (SPT) ≥3mm showed reduced sensitisation and two studies which used SPT >0mm and specific immunoglobulin E test showed no effect. | Not serious; Populations and interventions are similar to those which might be targeted outside of trial settings. Some studies used food powder, and some used normal foods. One study included infants with eczema. | Serious; Although the estimate indicates reduced allergic sensitisation, the 95% CI includes the possibility of no effect. | Insufficient studies to undertake formal testing of publication bias | Intervention: introducing cow's milk<br>Outcome: sensitisation *to any food*<br>Results as for "earlier introduction of multiple allergenic foods"; no study of cow's milk introduction without other foods, reporting this outcome, was identified. |
| | **7** | RCT | Not serious; Most information is from studies at low risk of bias or some concerns for one domain only. | Serious; $I^2$=45 %; heterogeneity may be partly explained by method of outcome measurement. Post hoc subgroup analysis showed a possible interaction for method of assessment (P=0.08): skin prick test (3 studies; 2974 participants) RR 0.64 (95% CI, 0.32-1.29; $I^2$=0 %); specific immunoglobulin E test (4 studies; 1913 participants) RR 1.29 (95% CI, 0.89-1.89; $I^2$=62 %). | Serious; Interventions were not all representative of typical ways to introduce cow's milk products to an infant's diet. Some studies used food powder, others used formula milk compared with a control group using soya formula, but often using very early and specific exposure regimes. | Serious; The 95% CI includes both a meaningful reduction and increase in risk or no effect. | Insufficient studies to undertake formal testing of publication bias | Intervention: introducing cow's milk (Intervention foods were milk (4 trials; 2071 participants); egg, milk, wheat, soya, buckwheat, and peanut (1 trial; 145 participants); milk, peanut, egg, sesame, white fish, and wheat (1 trial; 1167 participants); peanut, milk, wheat, and egg (1 trial; 1504 participants))<br>Outcome: sensitisation *to cow's milk* |

CI=Confidence interval, CF=Complementary food, RoB=Risk ob Bias, RCT=Randomised controlled trial, SR=Systematic review

### eTable 10. Summary of Findings of Secondary Outcomes

| Systematic Review | Intervention and Comparator | Age exposure | Outcome details | Age outcome | Effect estimaes and 95% Confidence intervals | Number of studies (participants) | Quality of the evidence (GRADE) | Comments |
|---|---|---|---|---|---|---|---|---|
| Incidence of adverse events (ADE), severe adverse events (SADE) | | | | | | | | |
| **Al-Saud et al. 2018** | Early introduction of egg powder ***vs.*** no early egg introduction | 3-9 m ***vs.*** exclusive breastfeeding until 6 m or placebo | Occurrence of anaphylaxis | Median 12 m | Anaphylaxis occurred in two participants in the intervention group and one participant in the control group, which was not statically significant (p=0.62) | 1 RCT [25] (820) | Not available | Study population: high-risk population |
| **EFSA Panel 2019** | Introduction of egg | 4 m ***vs.*** >8 m | Occurrence of anaphylaxis to egg powder (adverse event) | 12 m | 31 % (n=15/49) of participants had a reaction to egg powder, one case of anaphylaxis | 1 RCT [24] (86)[b] | Not available | Study population: high-risk population |
| **Smith et al. 2016** | Early introduction of potentially allergenic food ***vs.*** exclusively breastfeeding | 16 weeks ***vs.*** 6 m | Mean difference in percentage of days with fever | 4-6 m | MD -0.70 days (-3.40-2.00)<br><br>Infants with additional foods had a fever on average 0.7% of days lower (95 % CI 3.4% fewer to 2 % more days) | 1 RCT [49] (119) | ⊕⊕◯◯ Low | Study population: general population |
| | Early introduction of potentially allergenic food ***vs.*** exclusively breastfeeding | 3-6 m | Risk of gastrointestinal events (diarrhoea, vomiting, constipation), colic, enterocolitis syndrome reactions | Up to 3 years of age | RR 2.00 (0.18-22.04)<br><br>Risk with exclusive breastfeeding infants: 2 per 1000<br>Risk with non-exclusive breastfeeding infants (foods): 3 per 1000 | 1 RCT [13] (1303) | ⊕⊕⊕◯ Moderate | Study population: Exclusively breastfed infants |

| Systematic Review | Intervention and Comparator | Age exposure | Outcome details | Age outcome | Effect estimaes and 95% Confidence intervals | Number of studies (participants) | Quality of the evidence (GRADE) | Comments |
|---|---|---|---|---|---|---|---|---|
| **Incidence of withdrawals from the study intervention** | | | | | | | | |
| **Scarpone et al. 2023** | Earlier introduction of multiple allergenic foods | 2-12 m[a] ***vs.*** 6-24m | Risk of withdrawal from study inter-vention | 12-60 m | RR 2.29 (1.45-3.63)<br><br>Sensitivity analysis for low RoB data:<br>RR 0.96 (0.06-15.15) | 5 RCTs [13,16-18,50] (4703) | ⊕⊕⊕◯ Moderate | Study population: high-risk population<br>High heterogeneity: $I^2$=89 %; $\tau^2$=0.16<br>Scarpone et al.[10] showed that the statistical heterogeneity was extatistical by high rates of withdrawal from the intervention in the two largest studies. Both studies used high allergen intake for multiple allergenic foods and normal foods rather than powders (see Scarpone et al 2023, p. E4). |
| **Scarpone et al. 2023** | Earlier egg introduction | 3-12 m ***vs.*** 6-24 m | Risk of withdrawal from study intervention | 3-60 m | RR 1.58 (1.12- 2.22)<br><br>Sensitivity analysis for low RoB data:<br>RR 1.62 (1.09-2-40) | 13 RCTs [13,16-18,22-26,29,50-52] (7442) | ⊕⊕◯◯ Low | Study population: high-risk and general population<br>High heterogeneity: $\tau^2$=0.27, p=0.00, $I^2$=90 %<br>There was low-certainty evidence that earlier introduction of egg was associated with increased risk of withdrawal.<br>There was an asymmetrical funnel plot (eFigure 5 in Supplement 1), with increased withdrawal in larger studies. (see Scarpone et al 2023, p. E4) |
| | Earlier peanut introduction | 3-12 m ***vs.*** 6-24 m | Risk of withdrawal from study intervention | 3-60 m | RR 1.91 (1.19-3.05)<br><br>Sensitivity analysis for low RoB data:<br>RR 0.75 (0.49-1.15) | 6 RCTs [13,15-18,50] (5343) | ⊕◯◯◯ Very low | Study population: high-risk and general population<br>High heterogeneity: $\tau^2$=0.22, p=0.00, $I^2$=91 % |
| | Earlier cow's milk introduction | 0-12 m **vs.** 3d-24 m | Risk of withdrawal from study intervention | 5-60 m | RR 1.05 (0.61-1.82)<br><br>Sensitivity analysis for low RoB data:<br>RR 0.96 (0.06-15.15) | 11 RCTs [13,16-18,20,21,31,37,48,50,53] (7895) | ⊕⊕◯◯ Low | Study population: high-risk and general population<br>High heterogeneity: $\tau^2$=0.51, p=0.00, $I^2$=94 % |
| | Earlier introduction of wheat | 2-12 m ***vs.*** 6-24 m | Risk of withdrawal from study intervention | 5-60 m | RR 2.34 (1.49-3.68) | 5 RCTs [13,16-18,50] (4658) | Not available | Study population: high-risk and general population<br>High heterogeneity ($I^2$=89 %) |

| Systematic Review | Intervention and Comparator | Age exposure | Outcome details | Age outcome | Effect estimaes and 95% Confidence intervals | Number of studies (participants) | Quality of the evidence (GRADE) | Comments |
|---|---|---|---|---|---|---|---|---|
| **Scarpone et al. 2023** | Earlier introduction of soya | 0-11 m ***vs.*** 3-12 m | Risk of withdrawal from study intervention | 5-48 m | RR 1.43 (0.80-2.54) | 6 RCTs [16,20,31,37,48,50] (2215) | Not available | Study population: high-risk and general population<br>Moderate heterogeneity ($I^2$=63 %) |
| | Earlier introduction of fish | 2-12 m **vs.** 6-24 m | Risk of withdrawal from study intervention | 5-60 m | RR 1.80 (0.56-5.74) | 3 RCTs [13,17,50] (2098) | Not available | Study population: high-risk and general population<br>High heterogeneity ($I^2$=94 %) |
| | Earlier introduction of crustaceans and tree nuts | 2-12 m ***vs.*** 6-24 m | Risk of withdrawal from study intervention | 5-60 m | RR 1.02 (0.69-1.50) | 2 RCTs [17,50] (795) | Not available | Study population: high-risk population<br>Low heterogeneity ($I^2$=0 %) |
| **Incidence of AEs, SADEs: impaired growth (height, weight, head circumference)** | | | | | | | | |
| **EFSA Panel 2019** | Early ***vs.*** late introduction of CF | 3-4 m ***vs.*** 6 m | Mean difference in weight-for-age z-score (WAZ) or attained body weight | Up to 3 y | WAZ:<br>MD (z-score) -0.01 (-0.29-0.28)<br><br>Attained body weight:<br>MD (g) -40 (-199-118)<br>Prediction interval:<br>95% PI [-509;-428] | 5 RCTs [13,49,54-56] (1598) | High level of confidence | Study population: high-risk and general population<br><br>Low heterogeneity ($I^2$=0 %) for both WAZ and attained body weight;<br>The assessment of the level of confidence followed an approach that was inspired by the approach proposed by the National Toxicology Program (NTP) Office of Health Assessment and Translation (OHAT) conducts literature-based evaluations to assess. Evidence derived from RCTs was initially attributed a high confidence level (i.e. ++++)[7]. |

| Systematic Review | Intervention and Comparator | Age exposure | Outcome details | Age outcome | Effect estimaes and 95% Confidence intervals | Number of studies (participants) | Quality of the evidence (GRADE) | Comments |
|---|---|---|---|---|---|---|---|---|
| **EFSA Panel 2019** | Early ***vs.*** late introduction of CF | 3-4 m ***vs.*** 6 m | Mean difference in weight-for-length(height)-z-scores (WL(H)Z) | Up to 3 y | **12 m:** MD (z-score) 0.11 (0-0.22) **36 m:** MD (z-score) -0.01 (-0.11-0.09) | 1 RCT [13] (1162) | High level of confidence | Study population: general population |
| | Early ***vs.*** late introduction of CF | 4 m ***vs.*** 6 m | Absolute body weight gain | 6-12 m | Not significant, no effect estimate reported in EFSA Panel et al. 2019 | 1 RCT [54] (141) | High level of confidence | |
| | Early ***vs.*** late introduction of CF | 3-4 m ***vs.*** 6 m | Mean difference in length(height)-for-age z-scores (L(H)AZ) or attained body length/height | Up to around 3 y of age | L(H)AZ MD (z-score) 0.04 (-0.23- 0.30)[13,49] Attained body length/ height: MD (mm) 0.80 (-0.5-2.2); Prediction interval: 95% PI [-3.1; 4.8] | 5 RCTs [13,49,54-56] (1598) | High level of confidence | Study population: high-risk and general population Low heterogeneity (*I*²=0% for both L(H)AZ and attained body length). |
| | Early ***vs.*** late introduction of CF | 4 m ***vs.*** 6 m | Absolute length gain | 6-12 m | Not significant, no effect estimate reported in EFSA Panel et al. 2019 | 1 RCT [54] (141) | High level of confidence | |
| | Early ***vs.*** late introduction of CF | 3-4 m ***vs.*** ≥6 | Difference in head circumference-for-age z-scores (HCZ) | Up to 3 years of age | **12m:** (z-score) 0.06 (-0.05-0.17)[13] **36m:** (z-score) 0.05 (-0.08- 0.18)[56] | 2 RCTs [13,56] (1162) | High level of confidence | Study population: general population Not significant, no effect estimate reported in EFSA Panel et al. 2019 |
| | Early ***vs.*** late introduction of CF | 3-4m ***vs.*** ≥6 | Mean difference in attained head circumference | Up to 3 years of age | MD (mm) 0.50 (-0.3- 1.2) Prediction interval: 95 % PI [-1.7;- 2.7] | 3 RCTs [13,55,56] (1338) | High level of confidence | Study population: general population Low heterogeneity (*I*²=0 %) |
| **Smith et al. 2016** | Early introduction of potentially allergenic | 4-6 m | Mean difference in weight change (gain) (g) | Up to 26 weeks | MD -39.48 g (-128.43- 49.48) | 2 RCTs [49,54] (260) | ⊕⊕◯◯ Low | |

| Systematic Review | Intervention and Comparator | Age exposure | Outcome details | Age outcome | Effect estimaes and 95% Confidence intervals | Number of studies (participants) | Quality of the evidence (GRADE) | Comments |
|---|---|---|---|---|---|---|---|---|
| **Smith et al. 2016** | food ***vs.*** exclusively breastfeeding | | | | Risk with Exclusive breast-feeding infants: The mean weight change (gain) (g) at 4 to 6 months was 1054 g | | | |
| **Incidence of ADEs, SADEs: early cessation of breastfeeding** | | | | | | | | |
| **Smith et al. 2016** | Early introduction of CF | 3 m **vs.** >6 m | Median duration of any breastfeeding | 1-3 y | 50.2 weeks in standard intervention group<br>49 weeks in early introduction group | 1 RCT [13] (1162) | Not available | Study population: general population<br>The median duration was similar in both groups[11]. |
| **Incidence of symptoms of sneeze, wheeze, cough, itch or flexural eczema** | | | | | | | | |
| **Al-Saud et al. 2018** | Early introduction of egg powder ***vs.*** no early egg introduction | 3-9 m ***vs.*** exclusive breastfeeding until 6 m/ placebo | Risk of asthma (wheezing) | Median 12 m | RR 1.12 (0.87–1.45), p=0.39 | 1 RCT [25] (820) | Not available | Study population: high-risk population<br>No significant difference in the development of asthma (wheeze) between the intervention and control groups. |
| **Smith et al. 2016** | Early introduction of CF and breastfeeding ***vs.*** exclusively breastfeeding | 4 m ***vs.*** 6 m | Mean difference in upper respiratory illness: prevalence/ percentage of days of cough, congestion, nasal discharge, hoarseness | Up to 26 weeks | Cough: MD 3.10 (-4.52 - 10.72)<br>Congestion: MD 3.60 (-3.41-10.61)<br>Nasal discharge: MD 4.20 (-1.13-9.53)<br>Hoarseness: MD 0.10 (-1.84-2.04) | 1 RCT [49] (119) | Not available | No difference between groups |

| Systematic Review | Intervention and Comparator | Age exposure | Outcome details | Age outcome | Effect estimaes and 95% Confidence intervals | Number of studies (participants) | Quality of the evidence (GRADE) | Comments |
|---|---|---|---|---|---|---|---|---|
| **Incidence of allergic sensitisation** | | | | | | | | |
| **Smith et al. 2016** | Early introduction of egg powder ***vs.*** no early egg introduction | 3-9 m ***vs.*** exclusive breastfeeding until 6 m/ placebo | Risk of egg sensitisation either by a positive skin test or a positive Immuno-CAP test | Median 12 m | RR 0.74 (0.58-0.95) p=0.02 | 5 RCTs [13,22,24-26] (2534) | ⊕⊕⊕◯ Moderate | Study population: high-risk and general population<br><br>Low heterogeneity ($I^2$=17 %)<br>All five studies showed a decrease in sensitisation in the early intervention group compared to the no early intervention group, except for the study by Bellach et al.[22], which showed the opposite association. |
| **Burgess et al. 2019** | Early introduction (continued to age 6 m-12 m) ***vs.*** varying later ages | 4-6 m **vs.** varying later ages | Odds of egg sensitisation | 12 m | OR 0.76 (0.61-0.95) | 5 RCTs [13,22,24-26] (2793)[c] | The quality assessment of the individual RCTs according to the "Cochrane review Quality assessment Scale" can be found in the supplemental material of Burgess et al. (2019) | Study population: high-risk and general population<br><br>Low heterogeneity ($I^2$=15.6 %, p=0.315) |

| Systematic Review | Intervention and Comparator | Age exposure | Outcome details | Age outcome | Effect estimaes and 95% Confidence intervals | Number of studies (participants) | Quality of the evidence (GRADE) | Comments |
|---|---|---|---|---|---|---|---|---|
| **Chmielewska et al. 2017** | Modified diet, life-style and environment with encouragement of breastfeeding (at least 4 m) and supplementation with a partially hydrolysed formula and delayed introduction of solids, avoidance of house dust mites, pet allergens, and tobacco smoke | >6 m ***vs.*** no intervention | Odds of wheat sensitisation | 15 y | OR 2.57 (0.26-24.94) | 1 RCT [57] (545) | Not available | Infants at risk of allergy breastfeeding compared to formula feeding did not affect risk of sensitisation at 15 years. |
| | Early ***vs.*** standard introduction of of six potentially allergenic foods | >3 m ***vs.*** 6 m | Odds of wheat sensitisation | 12 m and 36 m | **12 months:** 7/550 ***vs.*** 19/601 (p=0.03) RR 0.40 (0.17-0.95) **36 months:** 8/569 ***vs.*** 19/599 (p=0.04) RR 0.44 (0.2-1.00) | 1 RCT [13] (1303) | Not available | Study population: Exclusively breastfed infants at population risk of allergy |
| **EFSA Panel 2019** | Timing of introduction of CFs in general | 3-4 m ***vs.*** 6 m | Risk of sensitisation to food | 12 m and 36 m | **12m:** RR 0.78 (0.60- 1.02) **36m:** RR 0.88 (0.62- 1.25) | 1 RCT [13] (1151/1173) | Not available | Study population: general population |
| **EFSA Panel 2019** | Timing of introduction of egg | 4-6m ***vs.*** no egg/ 3-4 m ***vs.*** 6 m | Odds of sensitisation to egg protein | 12 m/ 36 m | OR 2.20 (0.68-7.13)/ OR 0.71 (0.45-1.12) | 2 RCTs [13,22] (1568)[c] | Not available | Study population: general population; EFSA Panel et al.[7] reported that the PP analysis by Perkin et al. showed a statistically significantly lower risk of developing sensitisation in the inter-vention group compared with controls, while this |

| Systematic Review | Intervention and Comparator | Age exposure | Outcome details | Age outcome | Effect estimaes and 95% Confidence intervals | Number of studies (participants) | Quality of the evidence (GRADE) | Comments |
|---|---|---|---|---|---|---|---|---|
| **EFSA Panel 2019** | | | | | | | | was not the case for the full analysis set (see EFSA Panel et al., p.82). |
| | | 4-6.5 m ***vs.*** ≥10 m | Risk of sensitisation to egg protein | 12 m | RR 0.71 (0.43-1.16) | 3 RCTs [24-26] (1225)[c] | Not available | Study population: high-risk population<br>Low heterogeneity ($I^2$=0 %, p=0.46)<br>EFSA Panel et al.[7] reported that one RCT[26] was powered to detect an effect on sensitisation, showed statistically significantly reduced odds of sensitisation.<br>The EFSA Panel et al. also mentioned that two studies[24,25], which were included in the meta-analysis, were individually underpowered to detect significant findings (see EFSA Panel et al. 2019, p.83). |
| | Timing of introduction of wheat | 3-4 m ***vs.*** 6 m | Risk of sensitisation to wheat protein | 12 m and 36 m | **12m:**<br>RR 0.40 (0.17-0.95)<br>**36m:**<br>RR 0.44 (0.20-1.00) | **12m:**<br>1 RCT [13] (1151)<br>**36m:**<br>1 RCT [13] (1168) | Not available | Study population: general population<br><br>The EFSA Panel et al.[7] reported that the results of the RCT with respect to sensitisation are inconsistent with those on symptomatic wheat allergy (see EFSA Panel et al. 2019, p.84). |
| | Timing of introduction of peanut | 3-4 m ***vs.*** 6 m | Risk of sensitisation to peanut | 12 m and 36 m | **12m:**<br>RR 0.68 (0.41-1.13)<br>**36m:**<br>RR 0.68 (0.40-1.15) | **12m:**<br>1 RCT [13] (1151)<br>**36m:**<br>1 RCT [13] (1168) | Not available | Study population: general population<br><br>The result for sensitisation in the study by Perkin et al. 2016[13] is consistent with the findings in relation to symptomatic peanut allergy (see EFSA Panel et al. 2019, p. 87). |
| **Ierodiakonou et al. 2016** | Introduction of egg | 3-6 m ***vs.*** ≥ 6 or placebo | Risk of sensitisation to egg | Up to 36 m (median at 12 m of age) | RR 0.77 (0.53-1.11)<br>Sensitivity analysis (1):<br>RR 0.71 (0.53-0.94)<br>$I^2$=0 %, p=0.5051 | 4 RCTs [13,24,28,43] (1786)<br>Sensitivity analysis:<br>(1) 3 RCTs [13,24,43] | Not available | Study population: high-risk and general population<br>Moderate heterogeneity ($I^2$=37 %, p=0.19)<br>Heterogeneity was due to the abstract publication by Bellach et al.[28], which used specific IgE rather |

| Systematic Review | Intervention and Comparator | Age exposure | Outcome details | Age outcome | Effect estimaes and 95% Confidence intervals | Number of studies (participants) | Quality of the evidence (GRADE) | Comments |
|---|---|---|---|---|---|---|---|---|
| **Ierodiakonou et al. 2016** | | | | | Sensitivity analysis (2):<br>RR 0.65 (0.46-0.92)<br>$I^2$=0 %, p=0.3695 | (1488) (studies with abstract publications)<br>(2): 2 RCTs [24,43] (321)<br>(studies at unclear risk of bias) | | than skin prick testing to determine egg sensitisation |
| | Introduction of peanut | 3 m ***vs.*** ≥ 6 m | Risk of peanut sensitisation | Up to 36 m | RR 0.68 (0.40-1.15) | 1 RCTs [13] (1168) | Not available | Study population: general population<br>Evidence based on a single study. |
| | Introduction of cow's milk | Up to 6m ***vs.*** placebo | Risk of cow's milk sensitisation | Up to 84 m | RR 0.72 (0.40-1.27) | 3 RCTs [13,20,31] (1571) | Not available | Study population: high-risk and general population<br>Low heterogeneity ($I^2$=0 %, p=0.8382) |
| **Larson et al. 2017** | Early vs. delayed introduction of egg | <4m ***vs.*** >8m | Prevalence of egg sensitisation | 12 m | Early exposure to egg did not result in significantly less egg sensitisation at 12 months of age.<br>36 % of the infants were sensitised to eggs prior to 4 months of age and before introduction of CFs. | 1 RCTs [24] (86)[c] | Evidence Level 1 (SORT) | Study population: high-risk population<br>Evidence Level 1 Strength of Recommendation Taxonomy (SORT) criteria defines RCTs at high quality. These studies include key patient-oriented outcomes such as improvement in morbidity or mortality, symptoms, and quality of life or decreased costs. |
| **Scarpone et al. 2023** | Earlier introduction of multiple allergic foods | 3-4 m ***vs.*** 6-7 m | Risk of any food sensitisation | 11-36 m | RR 0.77 (0.54-1.10)<br>Sensitivity analysis for low RoB data:<br>RR 0.75 (0.49-1.15) | 3 RCTs [13,16,18] (2827) | ⊕⊕○○<br>Low | Study population: high-risk and general population<br>High heterogeneity:<br>$I^2$=73 %, $\tau^2$=0.07, $X^2$=7.40, df=2, p=0.02 |
| | Earlier introduction of egg | 3-4 m ***vs.*** 6-7 m | Risk of any food-sensitisation | 11-36 m | RR 0.86 (0.71-1.05)<br>Sensitivity analysis for low RoB data:<br>RR 0.75 (0.75-1.15) | 3 RCTs [13,16,18] (2827) | ⊕⊕○○<br>Low | Study population: high-risk and general population<br>Results as for "earlier introduction of multiple allergenic foods"; No study of egg introduction without other food, reporting this outcome;<br>Hihg heterogeneity ($I^2$=73 %) |

| Systematic Review | Intervention and Comparator | Age exposure | Outcome details | Age outcome | Effect estimaes and 95% Confidence intervals | Number of studies (participants) | Quality of the evidence (GRADE) | Comments |
|---|---|---|---|---|---|---|---|---|
| **Scarpone et al. 2023** | | 6 m ***vs.*** 6-12 m | Risk of allergic sensitisation to egg | 11-36 m | RR 0.81 (0.69-0.96)<br>Sensitivity analysis for low RoB data:<br>RR 0.85 (0.74-0.99) | 8 RCTs [13,16,18,22,24-26,51] (4325) | ⊕⊕⊕◯<br>Moderate | Study population: high-risk and general population<br>Low heterogeneity:<br>$\tau^2$=0.01, $X^2$=8.55, df=7, p=0.29, $I^2$=18 %;<br>Earlier introduction of egg was associated with decreased risk of egg sensitisation. |
| | Earlier introduction of peanut | 3-10 m ***vs.*** 6-60 m | Risk of sensitisation to any food | 11-60 m | RR 0.86 (0.71-1.05)<br>Sensitivity analysis for low RoB data:<br>RR 0.75 (0.49-1-15) | 4 RCTs [13,16,18,58] (3456) | ⊕⊕◯◯<br>Low | Study population: high-risk and general population |
| | Earlier introduction of peanut | 3-10 m ***vs.*** 6-60 m | Risk of allergic sensitisation to peanut | 11-60 m | RR 0.74 (0.46-1.20)<br>Sensitivity analysis for low RoB data:<br>RR 0.62 (0.30-1.25) | 4 RCTs [13,16,18,58] (3434) | ⊕⊕◯◯<br>Low | Study population: high-risk and general population<br>High heterogeneity:<br>$\tau^2$=0.18, $X^2$=13.24, df=3, p=0.00, $I^2$=77 % |
| | Earlier introduction of cow's milk | 3-4 m ***vs.*** 6-7 m | Risk of sensitisation to any food | 11-36 m | RR 0.77 (0.54-1.10)<br>Sensitivity analysis for low RoB data:<br>RR 0.75 (0.49-1.15) | 3 RCTs [13,16,18] (2827) | ⊕⊕◯◯<br>Low | Study population: high-risk and general population<br>High heterogeneity: $I^2$=73 %<br>Results as for "earlier introduction of multiple aller-genic foods"; no study of cow's milk introduction without other foods, reporting this outcome, was identified. |
| | | 0-4 m ***vs.*** 3 d-9 m | Risk of cow's milk sensitisation | 11-60 m | RR 1.14 (0.82-1.59)<br>Sensitivity analysis for low RoB data:<br>RR 1.11 (0.62-1.98) | 7 RCTs [13,16,18,20,21,31,53] (4887) | ⊕⊕◯◯<br>Very low | Study population: high-risk and general population<br>Moderate heterogeneity:<br>$\tau^2$=0.07, $X^2$=10.91, df=6, p=0.09, $I^2$=45 % |
| | Earlier introduction of wheat | 3-4 m ***vs.*** 6-7 m | Risk of allergic sensitisation to any food | 11-36 m | RR 0.77 (0.54-1.10); | 3 RCTs [13,16,18] (2827) | ⊕⊕◯◯<br>Low | Study population: high-risk and general population<br>Hig heterogeneity ($I^2$=73 %; $\tau^2$=0.07) |
| | | 3-4 m ***vs.*** 6-7 m | Risk of allergic sensitisation to wheat | 11-36 m | RR 0.62 (0.29-1.34) | 3 RCTs [13,16,18] (2818) | | Study population: high-risk and general population<br>Moderate heterogeneity ($I^2$=59 %); |

| Systematic Review | Intervention and Comparator | Age exposure | Outcome details | Age outcome | Effect estimaes and 95% Confidence intervals | Number of studies (participants) | Quality of the evidence (GRADE) | Comments |
|---|---|---|---|---|---|---|---|---|
| | Earlier introduction of soy | 3-4 m | Risk of allergic sensitisation to any food | 11-13 m | 65/76 ***vs.*** 68/74 participants in the earlier vs. later introduction group developed allergic sensitisation to any food | 1 RCT [16] (150) | | Study population: high-risk population<br>Only one study of introduction of soya (with other foods), reporting this outcome, was identified (150 participants). |
| | | 0-4 m ***vs.*** 6-9 m | Risk of allergic sensitisation to soya | 11-36 m | RR 1.14 (0.79-1.65) | 2 RCTs [16,31] (192) | | Study population: high-risk population<br>Low heterogeneity ($I^2$=0 %); |
| | Earlier introduction of fish | 3 ***vs.*** ≥ 6m | Risk of allergic sensitisation to any food | 3 y | 51/572 ***vs.*** 31/601 participants in the earlier vs. later introduction group developed allergic sensitisation to any food | 1 RCT [13] (1173) | | Study population: general population<br>Only one study of introduction of soya (with other foods), reporting this outcome, was identified (1173 participants). |
| **Scarpone et al. 2023** | | 3 ***vs***. ≥ 6m | Risk of allergic sensitisation to fish | 3 y | 4/567 ***vs.*** 5/599 participants in the earlier vs. later introduction group developed allergic sensitisation to fish | 1 RCT [13] (1166) | | Study population: general population<br>Only one study of introduction of soya (with other foods), reporting this outcome, was identified (1166 participants). |

[a] The results are extracted from the primary study as the reporting in the systematic review by Larson was inconsistent between relative and absolute risk reduction.

[b] Number of participants (at baseline) was extracted from the primary studies by the overview authors due to missing reported data in the systematic review.

[c] high quality - where we are very confident that the true effect lies close to that of the estimate of the effect; moderate quality - where we are moderately confident in the effect estimate, such that the true effect is likely to be close to the estimate of the effect, but there is a possibility that it is substantially different; low quality - where our confidence in the effect estimate is limited and the true effect may besubstantially different from the estimate of the effect; very low quality - where we have very little confidence in the effect estimate and the true effect is likely to be substantially different from the estimate of the effect.[59]

RCT=Randomised controlled trial, SR=Systematic review, RR=Risk ratio, OR=Odds ratio, RD=Risk difference, CF=Complementary food/feeding, WAZ=Weight-for-age z-score, WL(H)Z=Weight-for-length(height)-z-scores, L(H)AZ=Length(height)-for-age z-scores, HCZ=Head circumference-for-age z-scores

### eTable 11. Outcome related Study Overlap of Primary Studies within Systematic Reviews

| Primary Outcomes | Study Overlap | Overall results |
|---|---|---|

**Food Allergy**
n=5 SRs
(8 RCTs)

| | Burgess et al. 2019 | De Silva et al. 2020 | EFSA Panel 2019 | Scarpone et al. 2023 |
|---|---|---|---|---|
| De Silva et al. 2020 | 100,0% | | | |
| EFSA Panel 2019 | 100,0% | 100,0% | | |
| Scarpone et al. 2023 | 14,3% | 14,3% | 14,3% | |
| Smith et al. 2016 | 100,0% | 100,0% | 100,0% | 14,3% |

Overall results: Moderate Overlap

**Eggy Allergy**
n=8 SRs
(9 RCTs)

| | Al-Saud et al. 2018 | Burgess et al. 2019 | Dai et al. 2019 | De Silva et al. 2020 | EFSA Panel 2019 | Ierodiakonou et al. 2016 | Larson et al. 2017 |
|---|---|---|---|---|---|---|---|
| Burgess et al. 2019 | 100,0% | | | | | | |
| Dai et al. 2019 | 100,0% | 100,0% | | | | | |
| De Silva et al. 2020 | 66,7% | 66,7% | 66,7% | | | | |
| EFSA Panel 2019 | 83,3% | 83,3% | 83,3% | 50,0% | | | |
| Ierodiakonou et al. 2016 | 83,3% | 83,3% | 83,3% | 50,0% | 66,7% | | |
| Larson et al. 2017 | 16,7% | 16,7% | 16,7% | 25,0% | 20,0% | 20,0% | |
| Scarpone et al. 2023 | 66,7% | 66,7% | 66,7% | 44,4% | 55,6% | 55,6% | 11,1% |

Overall results: Very High Overlap

**Peanut Allergy**
n=7 SRs
(5 RCTs)

| | Burgess et al. 2019 | Dai et al. 2019 | De Silva et al. 2020 | EFSA Panel 2019 | Ierodiakonou et al. 2016 | Larson et al. 2017 |
|---|---|---|---|---|---|---|
| Dai et al. 2019 | 100,0% | | | | | |
| De Silva et al. 2020 | 33,3% | 33,3% | | | | |
| EFSA Panel 2019 | 50,0% | 50,0% | 0,0% | | | |
| Ierodiakonou et al. 2016 | 100,0% | 100,0% | 33,3% | 50,0% | | |
| Larson et al. 2017 | 50,0% | 50,0% | 50,0% | 0,0% | 50,0% | |
| Scarpone et al. 2023 | 50,0% | 50,0% | 20,0% | 25,0% | 50,0% | 25,0% |

Overall results: Very High Overlap

| Primary Outcomes | Study Overlap | | | | | Overall results |
|---|---|---|---|---|---|---|
| **Wheat Allergy**<br>n=3 SRs<br>(3 RCTs) | | Chmieleweska et al. 2017 | EFSA Panel 2019 | | | Very High Overlap |
| | EFSA Panel 2019 | 100,0% | | | | |
| | Scarpone et al. 2023 | 33,3% | 33,3% | | | |
| **Cow's Milk Allergy**<br>n=3 SRs<br>(6 RCTs) | | Dai et al. 2019 | Ierodiakonou et al. 2016 | | | Very High Overlap |
| | Ierodiakonou et al. 2016 | 100,0% | | | | |
| | Scarpone et al. 2023 | 33,3% | 33,3% | | | |
| **Eczema**<br>n=5 SRs<br>(17 RCTs) | | Al-Saud et al. 2018 | EFSA Panel 2019 | Ierodiakonou et al. 2016 | Smith et al. 2016 | Slight Overlap |
| | EFSA Panel 2019 | 100,0% | | | | |
| | Ierodiakonou et al. 2016 | 0,0% | 0,0% | | | |
| | Smith et al. 2016 | 0,0% | 0,0% | 0,0% | | |
| | Waidyatillake et al. 2019 | 0,0% | 0,0% | 0,0% | 50,0% | |
| **Allergic Rhinitis**<br>n=2 SRs<br>(14 RCTs) | | EFSA Panel 2019 | | | | Slight Overlap |
| | Ierodiakonou et al. 2016 | 0,0% | | | | |

| Secondary Outcomes | Study Overlap | | | Overall results |
|---|---|---|---|---|
| **Adverse events**<br>n=3 SRs<br>(3 RCTs) | | Al-Saud et al. 2018 | EFSA Panel 2019 | Slight Overlap |
| | EFSA Panel 2019 | 0,0% | | |
| | Smith et al 2016 | 0,0% | 0,0% | |

<table>
<tr><th>Secondary Outcomes</th><th>Study Overlap</th><th>Overall results</th></tr>
<tr><td>Impaired growth<br>n=2 SRs<br>(5 RCTs)</td><td><table><tr><th></th><th>EFSA Panel 2019</th></tr><tr><td>Smith et al. 2016</td><td>40,0%</td></tr></table></td><td>Very High Overlap</td></tr>
<tr><td>Allergy sensitisation to egg<br>n=6 SRs<br>(10 RCTs)</td><td><table><tr><th></th><th>Al-Saud et al. 2018</th><th>Burgess et al. 2019</th><th>EFSA Panel 2019</th><th>Ierodiakonou et al. 2016</th><th>Larson et al. 2017</th></tr><tr><td>Burgess et al. 2019</td><td>100,0%</td><td></td><td></td><td></td><td></td></tr><tr><td>EFSA Panel 2019</td><td>100,0%</td><td>100,0%</td><td></td><td></td><td></td></tr><tr><td>Ierodiakonou et al. 2016</td><td>28,6%</td><td>28,6%</td><td>28,6%</td><td></td><td></td></tr><tr><td>Larson et al. 2017</td><td>20,0%</td><td>20,0%</td><td>20,0%</td><td>25,0%</td><td></td></tr><tr><td>Scarpone et al. 2023</td><td>62,5%</td><td>62,5%</td><td>62,5%</td><td>20,0%</td><td>12,5%</td></tr></table></td><td>Very High Overlap</td></tr>
<tr><td>Allergy sensitisation to any food<br>n=2 SRs<br>(4 RCTs)</td><td><table><tr><th></th><th>EFSA Panel 2019</th></tr><tr><td>Scarpone et al. 2023</td><td>25,0%</td></tr></table></td><td>Very High Overlap</td></tr>
<tr><td>Allergy sensitisation to peanut<br>n=3 SRs<br>(4 RCTs)</td><td><table><tr><th></th><th>EFSA Panel 2019</th><th>Ierodiakonou et al. 2016</th></tr><tr><td>Ierodiakonou et al. 2016</td><td>100,0%</td><td></td></tr><tr><td>Scarpone et al. 2023</td><td>25,0%</td><td>25,0%</td></tr></table></td><td>Very High Overlap</td></tr>
<tr><td>Allergy sensitisation to wheat<br>n=3 SRs<br>(4 RCTs)</td><td><table><tr><th></th><th>Chmielewska et al. 2017</th><th>EFSA Panel 2019</th></tr><tr><td>EFSA Panel 2019</td><td>50,0%</td><td></td></tr><tr><td>Scarpone et al. 2023</td><td>25,0%</td><td>33,3%</td></tr></table></td><td>Very High Overlap</td></tr>
<tr><td>Allergy sensitisation to cow's milk<br>n=2 SRs<br>(7 RCTs)</td><td><table><tr><th></th><th>Ierodiakonou et al. 2016</th></tr><tr><td>Scarpone et al. 2023</td><td>42,9%</td></tr></table></td><td>Very High Overlap</td></tr>
</table>

### eAppendix 1 Deviations from the protocol

We did not search all preregistered databases (National Institute for Health and Clinical Excellence search engine/website and the National Institute for Health Care Reserahc Health Technology Assessment search engine website), Evidence for Policy and Practice Information Centre search engine/website/list). We did not use piloted data extraction forms, but relied on adapted forms that were provided within the PRIOR guideline document and the Cochrane Collaboration. More scientists were involved in data extraction, data analysis, data synthesis, and assessment of the quality of SRs than originally planned as more researchers joined and supported the author team. In addition, RoB assessment of primary studies within included reviews was added. This was necessary, as the reporting of RoB across SRs was very inconsistent and not up-to-date which also impacted GRADE assessments.The participant characteristics sex and ethnicity were not included in the data extraction.

### eAppendix 2 Search Strategy

### Medline (Ovid)

(exp infant/ or Child, Preschool/ or (child or children).ti,ab,kf. or (pre-school$ or preschool$).ti,ab,kf. or Nurseries/ or (nursery or nurseries).ti,ab,kf. or exp Parents/ or (parent or parents or mother or mothers).ti,ab,kf. or (infant or infants).ti,ab,kf. or infancy.ti,ab,kf. or toddler?.ti,ab,kf. or (baby or babies).ti,ab,kf. or newborn$.ti,ab,kf. or neonat$.ti,ab,kf. or Pediatrics/ or (pediatric$ or paediatric$).ti,ab,kf. or early childhood.ti,ab,kf. or (Pregnant Women/ or Pregnancy/ or Prenatal Nutritional Physiological Phenomena/) or pregnan$.ti,ab,kf. or Prenatal Exposure Delayed Effects/ or Maternal Exposure/ or ((maternal or prenatal) adj1 exposure$).ti,ab,kf. or (fetus or fetuses or fetal or foetus or foetuses or foetal).ti,ab,kf. or Fetus/ )

AND

(exp Preventive Health Services/ or Preventive Medicine/ or "prevention control".fs. or prevent$.ti,ab,kf. or prophyla$.ti,ab,kf. or Infant Formula/ or (formula or supplement$).ti,ab,kf. or ((risk or protect$ or development or avoidance or exposure or introduction) adj6 (allerg$ or hypersensitivit$ or atopy or atopic or dermatitis or neurodermatitis or asthma)).ti,ab,kf. )

AND

(exp Hypersensitivity/ or Allergens/ or allerg$.ti,ab,kf. or hypersensitivit$.ti,ab,kf. or prick test$.ti,ab,kf. or exp asthma/ or Dyspnea/ or (asthma$ or dyspnea or wheezing).ti,ab,kf. or (difficult$ adj1 breathing).ti,ab,kf. or rhinoconjunctivitis.ti,ab,kf. or (atopic adj1 (dermatit$ or neurodermatit$ or eczema or disease)).ti,ab,kf. or Diaper Rash/ or ((infant or infantile or diaper) adj1 (rash or rashes or eczema or dermatit$)).ti,ab,kf. or Disseminated Neurodermat$.ti,ab,kf. )

AND

((systematic review) or (meta-analysis) NOT (exp animals/ not humans.sh.))

And

(2010:2021.(sa_year))

### Pubmed

("allergie"[All Fields] OR "hypersensitivity"[MeSH Terms] OR "hypersensitivity"[All Fields] OR "allergies"[All Fields] OR "allergy"[All Fields] OR "allergy and immunology"[MeSH Terms] OR ("allergy"[All Fields] AND "immunology"[All Fields]) OR "allergy and immunology"[All Fields] OR ("eczema"[MeSH Terms] OR "eczema"[All Fields] OR "eczemas"[All Fields]))

AND

("prevent"[All Fields] OR "preventability"[All Fields] OR "preventable"[All Fields] OR "preventative"[All Fields] OR "preventatively"[All Fields] OR "preventatives"[All Fields] OR "prevented"[All Fields] OR "preventing"[All Fields] OR "prevention and control"[MeSH Subheading] OR ("prevention"[All Fields] AND "control"[All Fields]) OR "prevention and control"[All Fields] OR "prevention"[All Fields] OR "prevention s"[All Fields] OR "preventions"[All Fields] OR "preventive"[All Fields] OR "preventively"[All Fields] OR "preventives"[All Fields] OR "prevents"[All Fields] OR

"dietary supplements"[MeSH Terms] OR ("dietary"[All Fields] AND "supplements"[All Fields]) OR "dietary supplements"[All Fields] OR "supplement"[All Fields] OR "supplement s"[All Fields] OR "supplemented"[All Fields] OR "supplementing"[All Fields] OR "supplements"[All Fields] OR "probiotic s"[All Fields] OR "probiotical"[All Fields] OR "probiotics"[MeSH Terms] OR "probiotics"[All Fields] OR "probiotic"[All Fields]) OR ("prebiotically"[All Fields] OR "prebiotics"[MeSH Terms] OR "prebiotics"[All Fields] OR "prebiotic"[All Fields])

AND

("systematic review"[Publication Type] OR "systematic reviews as topic"[MeSH Terms] OR "systematic review"[All Fields] OR "meta analysis"[Publication Type] OR "meta analysis as topic"[MeSH Terms] OR "meta analysis"[All Fields])

AND

(2000/01/01:3000/12/31[Date - Publication])

### Web of Science (core collection)

(TS=(child*) OR TS=(infant*) OR TS=(pediatric) OR TS=(paediatric) OR TS=(babies) OR TS=(toddler*))

AND

(TS=(prevent*) OR TS=(protect*))

AND

(TS=(allerg*) OR TS=(eczema) OR TS=(dermatitis) OR TS=(atop*) OR TS=(asthma) OR TS=(sensiti*))

AND

(TS=(systematic review) OR TS=(review) OR TS=(meta-analysis))

AND

(PY=(2000-2021))

59. Schünemann H, Brożek J, Guyatt G, Oxman A, editors. The GRADE Working Group: GRADE handbook for grading quality of evidence and strength of recommendations. https://gdt.gradepro.org/app/handbook/handbook.html.
